# Large-Scale Mendelian Randomization Study Reveals Circulating Blood-based Proteomic Biomarkers for Psychopathology and Cognitive Task Performance

**DOI:** 10.1101/2024.01.18.24301455

**Authors:** Upasana Bhattacharyya, Jibin John, Max Lam, Jonah Fisher, Benjamin Sun, Denis Baird, Chia-Yen Chen, Todd Lencz

## Abstract

Research on peripheral (blood-based) biomarkers for psychiatric illness has typically been low-throughput, and traditional case-control studies are subject to potential confounds of treatment and other exposures. Here, we leverage large-scale, high-throughput proteomics data and Mendelian Randomization (MR) to examine the causal impact of circulating proteins on neuropsychiatric phenotypes. We utilized plasma proteomics data from the UK Biobank (3,072 proteins / 34,557 individuals) and deCODE Genetics (4,719 proteins / 35,559 individuals). Significant proteomic quantitative trait loci served as MR instruments, with the most recent GWAS for schizophrenia, bipolar disorder, major depressive disorder, and cognitive task performance as phenotypic outcomes. MR revealed 109 Bonferroni-corrected causal associations (44 novel) involving 88 proteins across the four phenotypes. Several immune-related proteins, including interleukins and complement factors, stood out as pleiotropic across multiple phenotypes. Identification of causal effects for these circulating proteins suggests potential biomarkers for these conditions and offers insights for developing innovative therapeutic strategies.

## Main

Psychiatric conditions present unique challenges in both research and treatment, in part due to the absence of clear and definitive biomarkers for these diseases^1^. Unlike many physical health conditions where diagnosis and monitoring can be supported by objective laboratory tests or imaging results, psychiatric disorders largely rely on subjective clinical assessments and patient self-reports^2,3^. This reliance on subjective measures not only hampers precision in diagnosing psychiatric conditions but also limits the development of targeted therapies^4^.

The lack of biomarkers in psychiatric conditions can be attributed to the lack of direct access to central nervous system tissue; brain-based biomarkers such as neuroimaging, while in some cases promising^5^, can be expensive and relatively low-resolution compared to the molecular processes underlying disease pathophysiology. By contrast, peripheral (e.g., blood-based) biomarkers have the potential advantage of ease of collection and clinical feasibility. However, research in this area has typically been low-throughput in terms of both the number of subjects and the range of assays performed^6^. Additionally, traditional case-control studies examining blood-based biomarkers are subject to potential confounds of treatment, hospitalization, and other environmental exposures common to patients with psychiatric illness, complicating the search for causal, disease-relevant pathophysiological mechanisms.

Genomewide association studies (GWAS) represent a high-throughput, comprehensive, and unbiased tool in providing insights into psychopathology; however, in-depth mechanistic insights that contribute to the disorders’ pathogenesis might be further elucidated by leveraging additional functional data. The growing utilization of high-throughput proteomics platforms in large-scale genotyped biobanks provides new opportunities to obtain biological insights from GWAS data^7–10^. Notably, the Mendelian Randomization (MR) approach^11^ permits the identification of causal mechanisms, avoiding the potentially confounding effects of environmental exposures while allowing for sample sizes that are orders of magnitude larger than conventional case-control reports.

Here, we provide the largest study utilizing MR to identify potentially causal effects of circulating proteins on psychiatric phenotypes. We employ blood-based proteomics data generated from two independent, large-scale cohorts (UK Biobank and deCODE Genetics) and well-powered GWAS for three psychiatric disorders: schizophrenia (SCZ), bipolar disorder (BIP), and major depressive disorder (MDD).

These disorders are strongly inter-correlated at the molecular genetic level^12^, and all three are marked by deficits in cognitive task performance (CTP)^13^. Moreover, we have demonstrated in our recent work that the genetic architecture of cognitive traits is intricately associated with psychopathology^14–16^. Consequently, we included CTP as an additional outcome phenotype in the MR analysis. In addition to identifying potential peripheral biomarkers for these outcome phenotypes, our downstream analyses aimed to yield pathophysiological insights and novel potential drug targets for psychiatric and nootropic interventions.

## Results

We have performed Mendelian randomization to investigate the causal impact of circulating proteins on SCZ, BIP, MDD and CTP (Figure 1, Supplementary Figure 1) by leveraging large-scale, high-throughput protein quantitative trait loci (pQTL) and genomic data obtained from individuals of European ancestry (see Methods). We performed MR analyses for cis-pQTLs and trans-pQTLs separately, using data obtained from two proteomic reference cohorts (UK Biobank and deCODE; Supplementary Tables 1a-1d).

**Figure 1.**
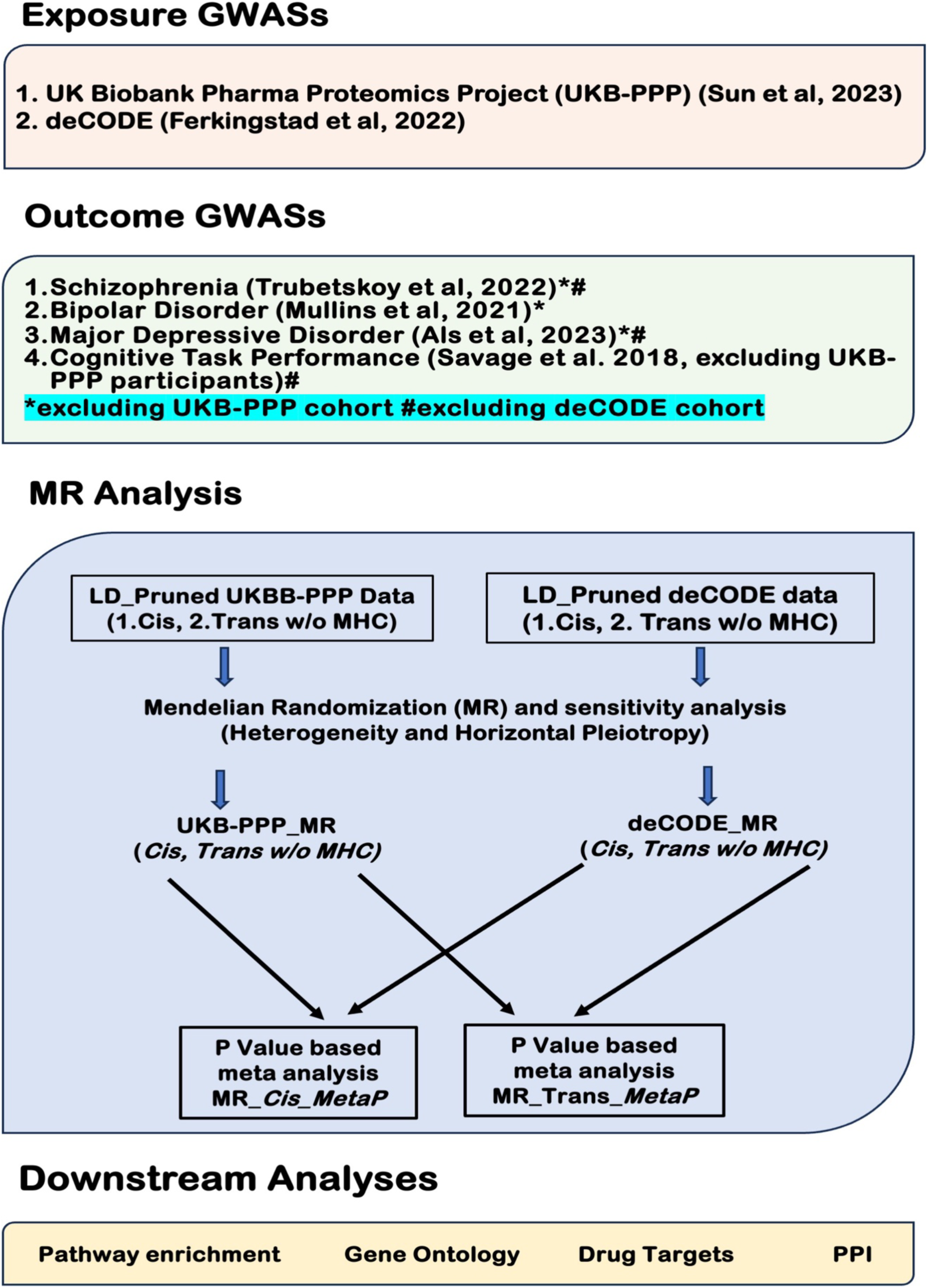
MR analysis workflow.

### Cis association

Mendelian Randomization analysis, employing cis-pQTL as instrumental variables, identified 74 proteins at the Bonferroni threshold described above that may be causally associated with the susceptibility to psychiatric disorders and cognitive ability (Table1, Figure 2). Specifically, 46 of these proteins were genetically predicted to have causal associations with SCZ, 16 with BIP, 8 with MDD, and 17 with CTP (Table1, Supplementary Figure 2 A, B, C, D; Supplementary Table). We note that in Figureure 2, the direction of effect for CTP was reversed to maintain consistency with the interpretation of psychiatric conditions (i.e., poorer cognition in the same direction as increased risk for psychiatric disorder). For example, Figureure 2 shows that higher protein levels of ITIH1 are significantly associated with decreased cognitive performance and increased risk for SCZ and BIP (Figure 2). All of the associated proteins exhibited at least nominal statistical significance (p<0.05) for the correct directionality (i.e., protein level causing the psychiatric or cognitive phenotype) using the MR Steiger approach; moreover, no protein showed significant horizontal pleiotropy (all p>0.05 for MR-Egger intercept test). However, eight proteins that were associated with SCZ and four proteins that were associated with CTP demonstrated nominally significant (p<0.05) levels of heterogeneity (Supplementary Table 2; see forest plots in Supplementary Figures 3A-3BK). Importantly, as demonstrated in Supplementary Table 2, these 12 proteins demonstrated inflated significance levels in the complementary/sensitivity analyses using fixed-effects IVW in the TwoSampleMR^17^ software. In some instances, raw p-values were as many as four orders of magnitude larger using TwoSampleMR than our primary method of determining significance, using multiplicative random effects in the ‘MendelianRandomization’ package. Similarly, many proteins would have been deemed statistically significant by fixed-effects IVW in TwoSampleMR, which did not attain significance in our analysis and are therefore not included in our results. (While there is no universally accepted gold standard amongst MR approaches, we selected the more conservative options available throughout this work.)

**Figure 2.**
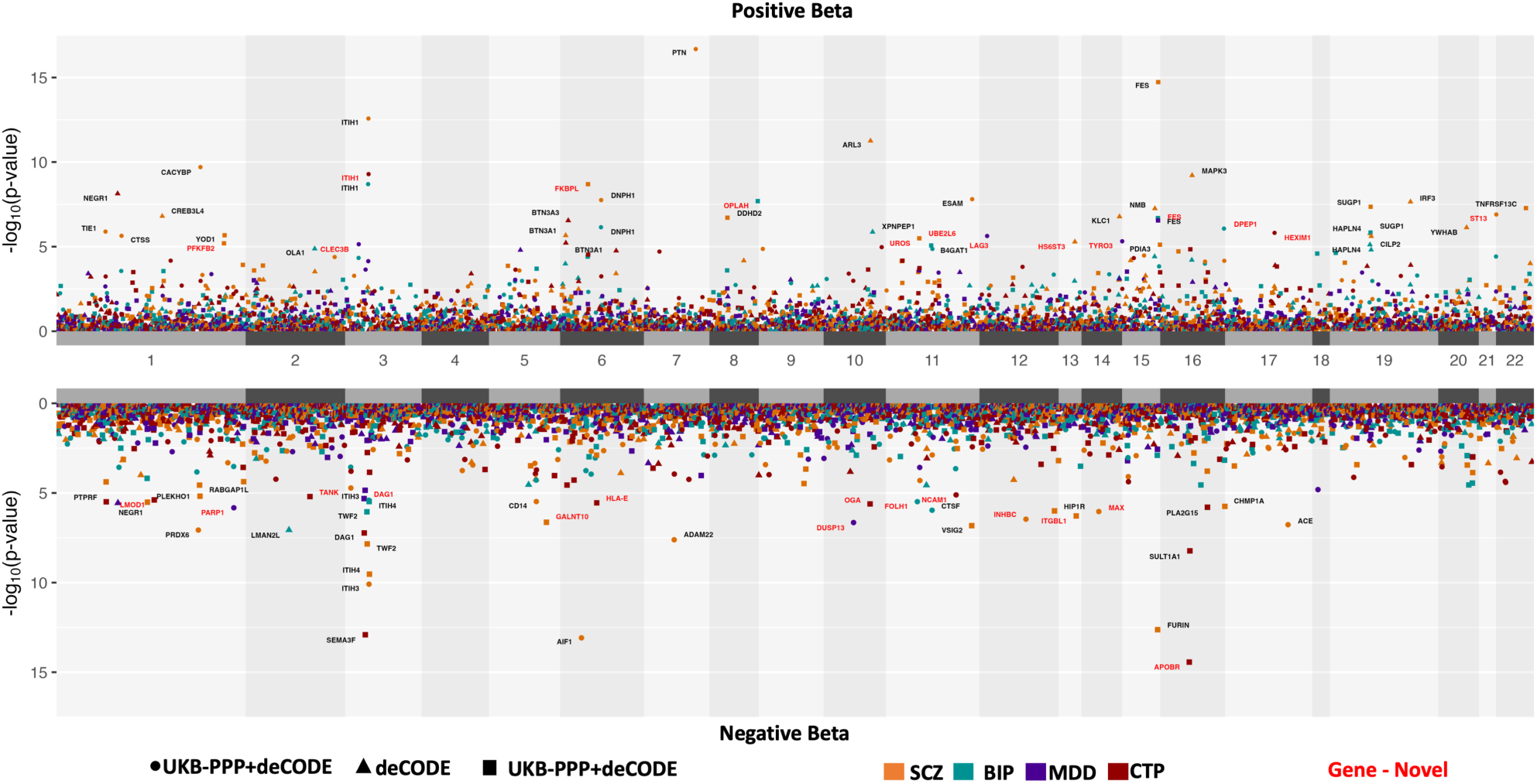
Manhattan plot showing findings from MR analysis for schizophrenia, bipolar disorder, major depressive disorder, and cognitive task performance, employing Cis-pQTLs from UKB-PPP and deCODE dataset as instrumental variables *Note*: Manhattan plots: (Top panel) proteins with positive predictive beta-values (Bottom panel) proteins with negative predictive beta-values relative to the traits investigated. X-axis: genomic coordinates/chromosomes; Y-axis: -log10p values of associations. Traits: Bipolar Disorder (turquoise), Cognitive ability (red), MDD (purple), Schizophrenia (orange). MR/Meta-Analysis: Meta-P (circle), deCODE: Results for deCODE specific MR analysis (triangle), UKB-PPP: Results for UKB-PPP specific MR analysis (square). Proteins based on more than one method, on multiple traits may be represented in the figure.

An extensive literature search was conducted to identify any prior Transcriptome-Wide Association Studies (TWAS), Summary-data-based Mendelian Randomization (SMR), Mendelian Randomization (MR), or colocalization analyses in SCZ, BIP, MDD, and CTP (Supplementary Table 3a-c), to ascertain the novelty of our findings. Notably, we examined literature examining not only blood-based but also brain-based QTL reference datasets; we also extended our search to both gene expression (eQTL) as well as proteomic (pQTL) data, as these can often differ^18^. Our literature review revealed that 9 of the 46 Bonferroni-significant proteins for SCZ had not been reported as statistically significant for the disorder. Similarly, 3 of the 16 proteins for BIP, 7 of 8 for MDD, and nine proteins for CTP had not been previously reported as significant (corrected for multiple comparisons) for that specific phenotype (Table1, Supplementary Table 3c).

If we employ a less strict 5% false discovery rate threshold (FDR<0.05), 210 proteins are genetically predicted to have a causal association with susceptibility to psychiatric disorders and/or cognitive performance. Specifically, 132 proteins were genetically predicted to have causal associations with SCZ, 55 with BIP, 13 with MDD, and 74 with CTP (Table1, Supplementary Table 2). Our literature search revealed that a large proportion of these associations were not previously reported as significant (at FDR<0.05); specifically, 37 of these proteins were for SCZ, 17 for BIP, 9 for MDD, and 50 were novel for CTP (Table1, Supplementary Table 3a-c). Additionally, all of the associated proteins were significant for directionality, and only two associated proteins (one in cognition and one in schizophrenia) were found to have nominally significant (p<0.05) horizontal pleiotropy. Using Cochran’s *Q* statistic derived from the IVW meta-analysis, 19 proteins in SCZ, five proteins in BIP, and 16 proteins in CTP were found to have nominally significant (p<0.05) heterogeneity (for forest plots, see Supplementary Figures 3A-3BK). However, as noted previously, our use of multiplicative random-effects IVW provides conservative p-values in the presence of such heterogeneity.

Our study comprises pQTL data from two cohorts, UKB-PPP and deCODE, which utilize different protein profiling platforms (Olink and SomaScan, respectively). A comprehensive comparative analysis has recently reported a moderate level of correlation of the proteins assessed via these two distinct platforms^19^. Our MR results are consistent with this report, particularly in the context of cis-pQTLs. Pearson correlation analysis, undertaken independently for each condition, revealed coefficients in the range of 0.46 to 0.53 for per-protein effect sizes obtained from MR analysis

(Supplementary Figure 2 A, B, C, D). Among the 74 proteins (PBONF<0.05) identified in our cis-pQTL MR analysis, 27 were assayed in both cohorts. The associations of these 27 proteins with their respective outcome phenotypes (16 for SCZ, 6 for BIP, 5 for MDD, and 4 for CTP) were replicated in both cohorts. Separately, 31 unique proteins were exclusively assayed in the UKB-PPP cohort (19 for SCZ, 6 for BIP, 2 for MDD, and 10 for CTP), and 16 unique proteins (11 for SCZ, 4 for BIP, 1 for MDD, and 3 for CTP) were specifically assayed in the deCODE cohort (Supplementary Table 2). Among the 210 proteins that were found to be associated with a looser threshold (FDR<0.05), 77 proteins were assayed in both cohorts, and all associations were replicated in both cohorts (45 for SCZ, 23 for BIP, 4 for MDD, and 31 for CTP). By contrast, 86 (56 for

SCZ, 20 for BIP, 7 for MDD, and 30 for CTP) and 47 (32 for SCZ, 12 for BIP, 2 for MDD, and 14 for CTP) unique proteins were specifically assayed only in UKB-PPP and deCODE cohort respectively (Supplementary Table 2).

### Trans association

The separate MR analysis employing trans-pQTLs as instrument variables identified 14 proteins meeting the Bonferroni-corrected threshold (PBONF<0.05) that may be causally associated with the susceptibility to psychiatric disorders and cognitive performance (Table1, Supplementary Table 2). Specifically, 6 of these proteins were genetically predicted to have causal associations with SCZ, 3 with BIP, 1 with MDD, and 7 with CTP. (Table1, Supplementary Table 2, Figure 3; Supplementary Figure 2 A, B, C, D). Our systematic review (Table1, Supplementary Table 3a-c) indicated that none of the findings from the trans-pQTL analysis had been previously reported as statistically significant. Moreover, all the identified proteins for schizophrenia, bipolar disorder, and major depressive disorder, including notable proteins such as IL23R and TNS2, have not been previously reported at even nominal or subthreshold levels of statistical significance (Table1, Supplementary Table 3a-c). All proteins associated with the trans-pQTL analyses demonstrated significance (P<0.05) in the MR Steiger test for causal direction, and none exhibited significant horizontal pleiotropy. However, three proteins associated with SCZ, one protein associated with BIP, and four proteins associated with CTP exhibited nominal significance (p<0.05) for heterogeneity (see forest plots in Supplementary Figurers 3A-BK).

**Figure 3.**
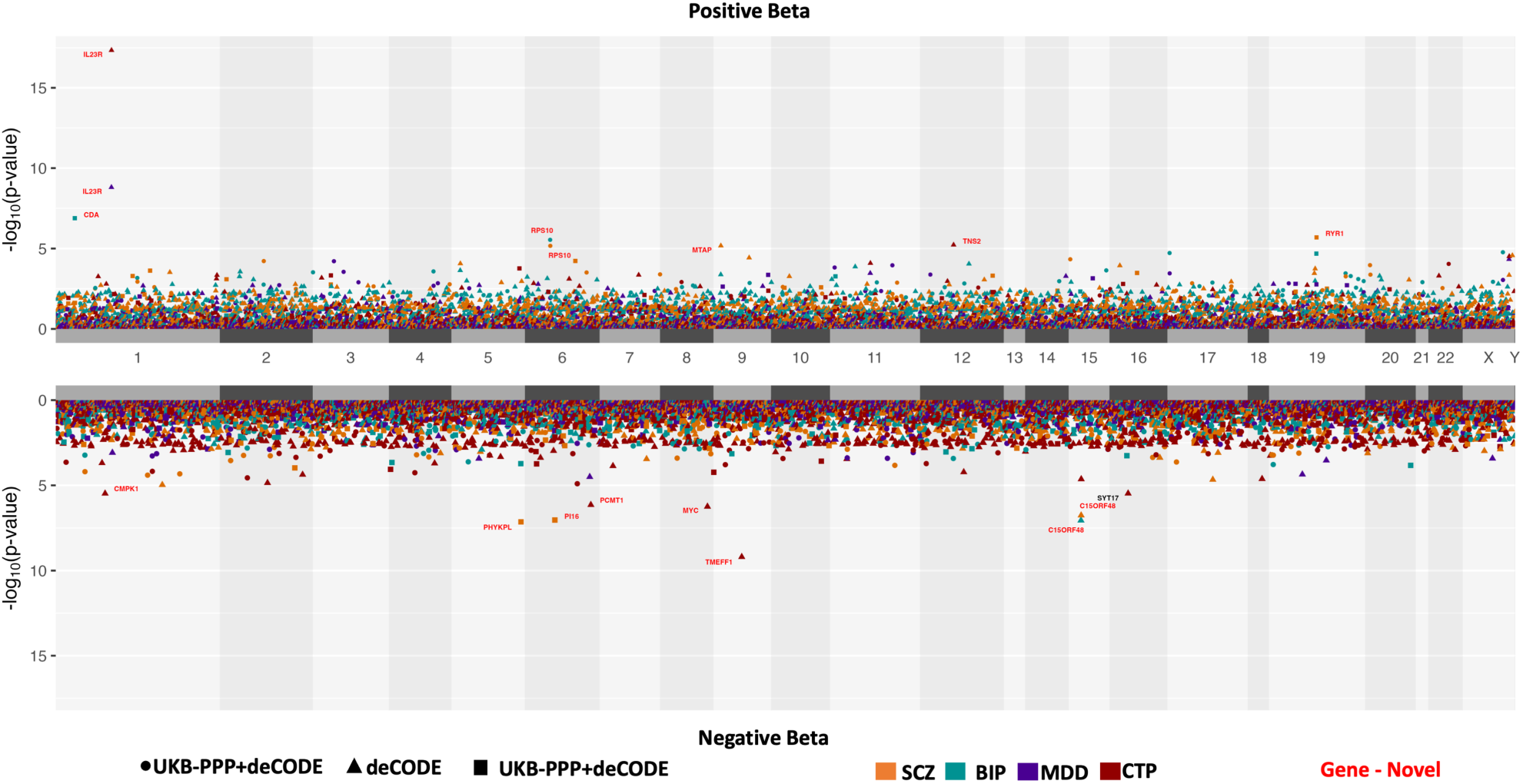
Manhattan plot showing findings from MR analysis for schizophrenia, bipolar disorder, major depressive disorder, and task performance employing Trans-pQTLs from UKB-PPP and deCODE datasets as instrumental variables *Note*: Manhattan plots: (Top panel) proteins with positive predictive beta-values (Bottom panel) proteins with negative predictive beta-values relative to the traits investigated. X-axis: genomic coordinates/chromosomes; Y-axis: -log10p values of associations. Traits: Bipolar Disorder (turquoise), Cognitive ability (red), MDD (purple), Schizophrenia (orange). MR/Meta-Analysis: Meta-P (circle), deCODE: Results for deCODE specific MR analysis (triangle), UKB-PPP: Results for UKB-PPP specific MR analysis (square). Proteins based on more than one method, on multiple traits may be represented in the figure.

At a less stringent threshold (FDR<0.05), associations of a total of 51 proteins were identified for the outcome phenotypes (Table1, Supplementary Table 2). Specifically, 21 proteins were associated with SCZ, 6 with BIP, 1 with MDD, and 28 with CTP. Our review of the literature on prior blood- or brain-based TWAS, SMR, and MR analysis revealed that 49 of our findings were novel. Additionally, all these associations demonstrated significance in the MR Steiger directionality test, and none of the proteins demonstrated significant horizontal pleiotropy. However, six proteins that were associated with CTP and five proteins associated with SCZ were found to have at least nominally significant (P<0.05) heterogeneity (Supplementary Table 2).

As observed previously with the proteins identified utilizing cis-pQTL, a notable portion of the proteins identified using trans-pQTLs also were found to be inflammatory modulators such as CRP, GRIK2, IL1RL2, ISL1, C15ORF48, CDA, F9, SERPINF2, and RYR1 (Supplementary Table 2). In our analysis of CTP, the presence of several complement and interleukins, such as CCL21, CD4, HLA-E, IL1A, and IL23R, stands out as noteworthy (Supplementary Table 2).

Unlike for cis-pQTLs, we observed only modest correlations of MR results across the two proteomics platforms, with varying coefficients across all four phenotypes in the range of 0.09 to 0.15 for per-protein effect sizes (Supplementary Figure 4 A, B, C, D).

Results were consistent with an earlier report that conducted a comprehensive comparative analysis of the proteins assessed via these two distinct platforms^19^. Only 1 protein (RPS10) among the 14 unique proteins (PBONF<0.05) identified in our study was assayed by both deCODE and UKB-PPP platform and it’s associations with SCZ and BIP were replicated in both platforms. In contrast, only 17 proteins (8 for SCZ, 3 for BIP, and 7 for CTP) among the 51 unique proteins (FDR<0.05) were assayed in both platforms (Supplementary Table 2).

### Overlap across phenotypes

We observed considerable evidence for pleiotropic effects of many proteins across phenotypes, providing further insights into the shared etiology among the psychiatric conditions and cognitive ability. In Figure 4A, we utilize cis-pQTL associations that are significant according to Bonferroni correction, showcasing all associations that are significant according to the FDR as well as those that are nominally significant (P<0.01) in our MR analysis for these proteins. For example, ITIH4 demonstrates significant associations with SCZ and BIP, enhancing the interpretability of the FDR-level association with CTP and nominally significant association with MDD. In parallel, TWF2 emerges as another protein with involvement in all four phenotypes (Figure 4 A, B). Interestingly, IL23R, a novel discovery in trans-pqtl analyses for MDD, displays an inverse relationship between cognition and depression (Figure 4 A, B). PARP1 also displays pleiotropic relationships in opposite directions; otherwise, all pleiotropic relationships are concordant for up- or down-regulation.

**Figure 4A.**
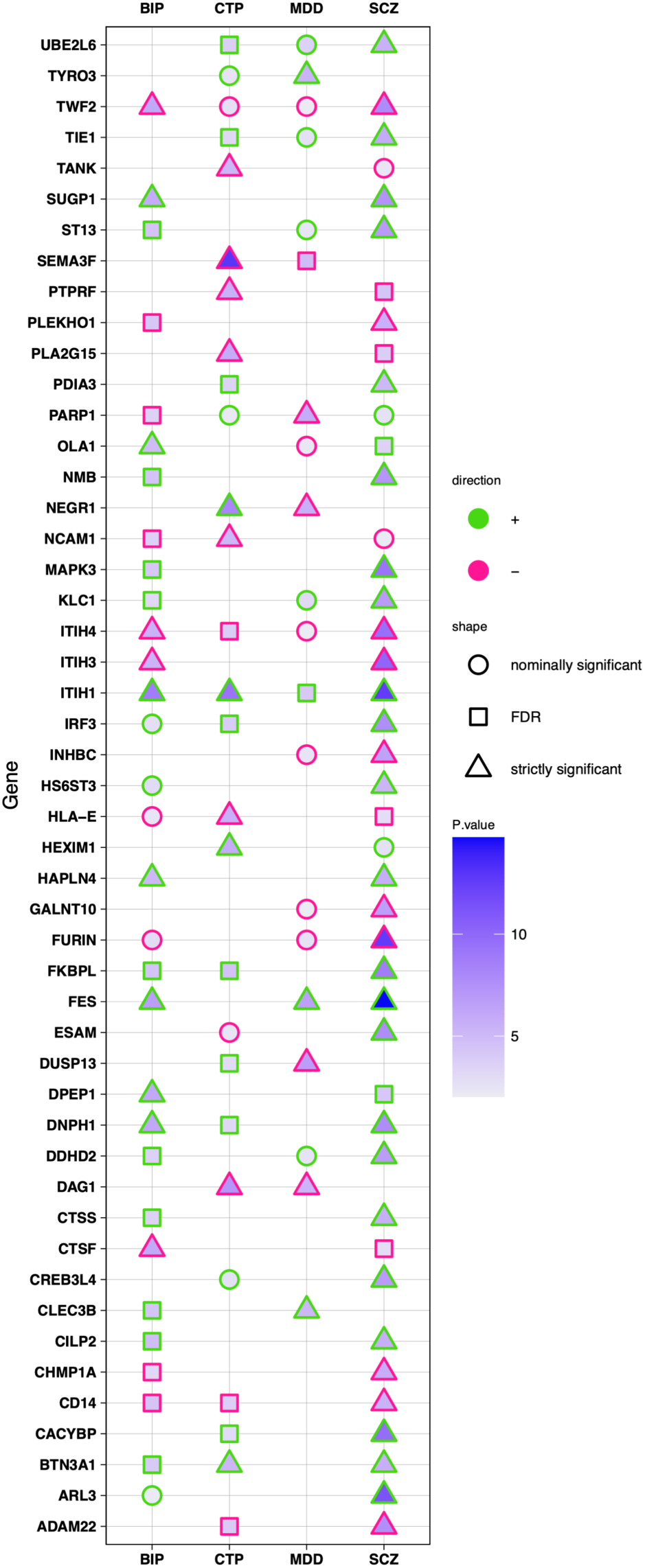
Cis-pQTL MR results across traits. *Note:* Proteins significantly associated with at least one trait are presented in the figure. The strength of MR association p-values is represented by a blue gradient across data values. The strength of association is further annotated for easy reference, nominally significant p < 0.05 (circle), FDR significant p_scz_ < 1.7 x 10^-3^ p_BIP_ < 7.3 x 10^-4^, p_MDD_ < 9 x 10^-9^ p_CTP_ < 1 x 10^-3^ (square), and Bonferroni significant p < 1.33761x 10^-5^ (triangle) are annotated accordingly.

**Figure 4B.**
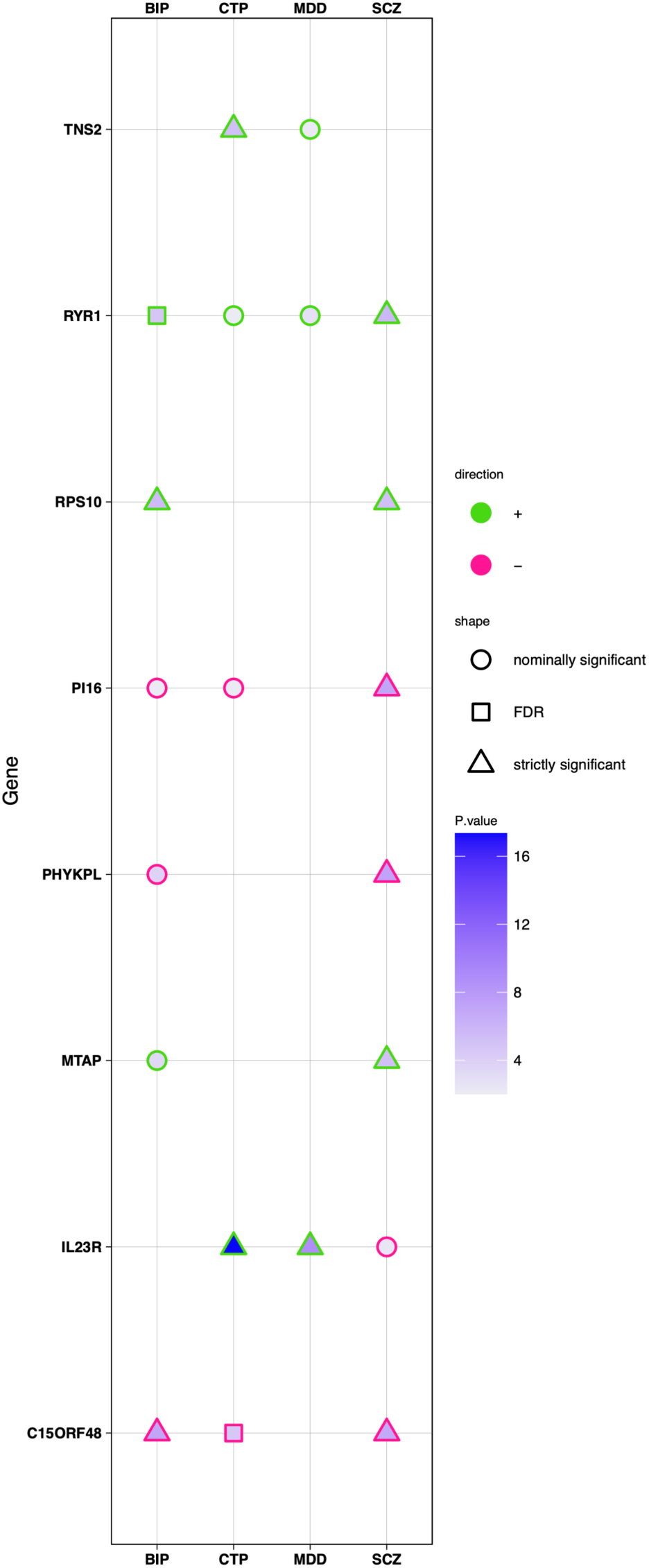
Trans-pQTL MR results across traits. *Note:* Proteins significantly associated with at least one trait are presented in the figure. The strength of MR association p-values is represented by a blue gradient across data values. The strength of association is further annotated for easy reference, nominally significant p < 0.05 (circle), FDR significant p_scz_ < 1.49 x 10^-4^, p_BIP_ < 2.07 x 10^-5^ p_MDD_ < 1.55 x 10^9^ p_CTP_ < 2 x 10^-7^ (square) and Bonferroni significant p < 7.34 x 10^-6^(triangle) are annotated accordingly.

### Pathway enrichment

Pathway enrichment analyses revealed that the circulating proteins significantly associated with SCZ are enriched in immune response pathways such as "Toll Receptor Signaling Pathway,” in addition to enrichment of shared proteins with neurodegenerative disorders such as Alzheimer’s and Parkinson’s (Supplementary Table 4a). In pathway enrichment analysis for BIP-associated proteins, immune-related pathways, platelet-related pathways, and peptidase-mediated proteolysis pathways were observed to be significantly enriched (Supplementary Table 4a). For MDD, in addition to pathways relating to immune response regulation, significant proteins were enriched in signaling pathways related to the MAPK cascade, which is involved in neuronal plasticity, function, and survival^20^ (Supplementary Table 4a). In line with psychiatric disorders, our pathway enrichment analysis for cognitive ability also unveiled several immune response-related pathways and neurodevelopmental and synaptic pathways such as Axon Guidance, Axonogenesis, and Neuron Projection Guidance (Supplementary Table 4a).

### Protein-Protein Interaction

MAPK3, FURIN, ITIH1, ITIH3 and ITIH4 were found to constitute the main hub of the PPI network in SCZ (Supplementary Figure 6A). A previous study of schizophrenia-associated CNV genes also identified MAPK3 as the most prominent network hub in protein–protein interaction networks within the 16p11.2 proximal region^21^. Additionally, TIE1 and ESAM were found to constitute a separate network, potentially related to blood-brain barrier function. The PPI networks for other phenotypes did not reveal many insights, although ITIH1 was found to be an important node in bipolar disorder (Supplementary Figure 6B). BTN3A1 and BTN3A13 were found to constitute an important node in CTP (Supplementary Figure 6 C). No nodes or PPI interaction were identified in MDD (Supplementary Figure 6 D).

### Drug Target Enrichment

Several of the proteins identified in our study, including PDIA3, ITIH4, ITIH3, CD14, AIF1, and CTSS, are targets of established nonsteroidal anti-inflammatory drugs such as Rofecoxib and Celecoxib (Table1, Supplementary Table 4b). SCZ-associated proteins were found to be enriched for known psychiatric agents such as modafinil and valproic acid, as well as calcium channel blockers which have frequently emerged from schizophrenia GWAS analysis^22^. The proteins identified in the SCZ analysis were also enriched for targets of nicotine, suggesting a possible rationale for the use of nicotine as a self-medicating agent among individuals with schizophrenia^23^. In addition to known antipsychotic and mood-stabilizing drugs such as olanzapine, carbamazepine, and valproic acid, proteins identified in bipolar disorder were found to be enriched for targets of amantadine, which is known for its anti-NMDA activity and suggested to be an important therapeutic option for acute bipolar depression (Supplementary Table 4b).

Among the proteins identified in this study for major depressive disorder, NEGR1 and TYRO3 were found to be targets of paroxetine, a widely prescribed selective serotonin reuptake inhibitor, as well as phenelzine, an MAO inhibitor is prescribed for MDD patients who have not responded to other treatments. Additionally, PARP, NEGR1, FES, TYRO3, and DAG1 were found to be targeted by retinoic acid (Supplementary Table 4b). Proteins identified in cognition were enriched for targets of yohimbine, which has been studied for nootropic and anxiety-reducing purposes ^24^ (Supplementary Table 4b).

### Gene-Drug Interaction and Potential Druggability

We identified 7 actionable drug targets for SCZ, 5 for BIP, 3 for MDD, and 4 for CTP (PBONF<0.05) (Table1, Supplementary Table 5a) using only strictly significant proteins obtained from MR analysis utilizing *cis*-pQTLs as genetic instruments. These potential drug targets include FES, MAPK3, ACE, CD14 ITIH3, HLA-E, and NCAM16 (Supplementary Table 5a). Our findings indicate that several of these proteins are targets of drugs already in use for psychiatric indications, such as ITIH3 with clozapine and *ACE* with sertraline. Additionally, a subset of these targets may identify potential opportunities for drug repurposing, including methotrexate (an immune suppressant and antimetabolite) for bipolar disorder and duloxetine (an antidepressant) for cognitive task performance. Perhaps most importantly, almost all the proteins identified in our study, with a few exceptions, were indicated to be druggable (Table1, Supplementary Table 5b).

## Discussion

The present study identified evidence of 109 Bonferroni-corrected causal associations between 88 proteins and one or more of our target phenotypes. Compared to the relevant existing literature across both blood- and brain-based studies of protein levels and/or gene expression, approximately half of these associations are significant for the first time in our study, including numerous pQTLs identified *in trans*. Loosening the statistical threshold to the FDR <0.05 level resulted in several hundred associations with widespread pleiotropic effects detected (Figureure 4A,B; Supplementary Table 2).

It is noteworthy that many of the proteins identified in our study are related to immune function, including interleukins, toll-like receptors, and complement factors (e.g., CF1, CD14, CD40, CD46, HLA-E, CF1, IL1RL2, and IL20RB). Furthermore, our investigation sheds light on several other proteins associated with the regulation of the inflammatory response, such as CREB3L4, CSK, CTSF, CTSS, ICAM5, INHBC, IRF3, ITGAL, and ITGBL1. While prior case-control studies^25^ have often identified heightened levels of pro-inflammatory cytokines such as IL-6, IL-8, and IL-1ꞵ in patients with serious mental illness, it is noteworthy that none of these emerged as significant in our analysis, consistent with evidence that medication^26,27^ and other state-related factors influence the levels of these biomarkers in patients with serious mental illness. Similarly, prior case-control research has replicably observed elevated blood levels of CRP in individuals with schizophrenia; however, these findings may be state-related^28,29^. In this context, it is noteworthy that our results indicated a *reduced* level of circulating CRP was associated with genetic risk for schizophrenia. Moreover, the strongest individual signal in the present study was IL23R; higher levels of IL23R were associated with increased risk of depression and poorer cognition. While elevated IL23 has been associated with depression in patients with psoriatic arthritis^30^ (for which IL23 is a major causal factor)^31^ there have been mixed reports concerning the correlation between IL23 levels in the blood of patients with idiopathic depression^32,33^.

More broadly, blood-based inflammatory markers identified in the present study should be tested as potential early indicators of the risk of developing psychiatric disorders and cognitive impairment. It is worth noting that most prior case-control studies have utilized low-throughput methods to assess circulating proteins; only three very recent studies have utilized the Olink or SomaScan platform to assess a large panel of proteins in serious mental illness^34–36^. Studies in patients at clinical high risk for psychosis, or in the first episode of psychosis, may be particularly informative due to relative lack of cumulative medication exposure, as well as providing information on the potential prognostic value of such blood-based proteomic biomarkers^37^.

In addition to immune and inflammatory proteins, our study was able to implicate many new proteins/genes by leveraging the large sample sizes available in our proteomic reference panels, and comparing our results to the systematic review of prior omics studies (Supplementary Table 3). For example, the apolipoprotein B receptor (APOBR) was the strongest cis-pQTL signal associated with cognitive task performance in the present study, and has not been previously reported in comparable studies. However, circulating levels of ApoB have been associated with risk for Alzheimer’s disease^38,39^ and age-related cognitive decline^40,41^. Similarly, we observed that increased levels of circulating PCMT1, which has been shown to be neuroprotective in animal models of cognitive aging^42^, were causally associated with better cognitive task performance. We also report, for the first time, a causal relationship between reduced levels of folate hydrolase I (FOLH1) and increased risk for bipolar disorder (as well as a nominally significant association with schizophrenia in the same direction). In the brain, this protein catalyzes the breakdown of N-acetylaspartylglutamate into glutamate, thereby serving as a high-level control of neurotransmission and excitotoxicity^43^; reductions in the levels of FOLH1 have been reported in post-mortem brain tissue of patients with schizophrenia^44^. Further studies to examine the relationship between blood levels of FOLH1 and glutamate neurotransmission in schizophrenia and bipolar disorder are warranted. Additionally, reduced levels of syntaxin binding protein 1 (STXBP1, critical to release of neurotransmitters from synaptic vesicles) were associated with schizophrenia. While this association has not been previously reported, it is noteworthy that loss of function mutations in this gene cause a range of neurodevelopmental disorders^45^.

Intriguingly, the pathway enrichment analysis in schizophrenia also reveals shared proteins with neurodegenerative disorders such as Alzheimer’s and Parkinson’s Disease, consistent with a recent brain-based proteomic study^46^ demonstrating unexpected overlap between schizophrenia and neurologic disorders. This raises the possibility of immune protein-mediated neurodegeneration as a potential contributing factor to the risk of schizophrenia. Pathway enrichment analysis for bipolar disorder also found enrichment of peptidase-mediated proteolysis pathways. Regulated proteolysis is a fundamental process for maintaining the health and proper functioning of neurons^47^, as well as for the dynamic remodeling of synapses during synaptic plasticity^48^. Disruptions or defects in any of these proteolytic pathways can have significant consequences, including impairments in neural communication, synaptic stability, and overall brain function, underscoring the critical role of regulated proteolysis in maintaining the proper functioning of the nervous system^47^.

A primary goal of GWAS is the identification of novel drug targets^47^, yet this process is complicated by the lack of specificity of the broad genomic loci typically identified by GWAS. Omics studies, such as the present study’s examination of pQTLs, permits the conversion of GWAS signals into specific drug targets^49^. Our drug target enrichment analysis successfully identified already-approved medications for each of the three psychiatric disease phenotypes (Supplementary Table 4b), providing evidence supporting our approach. In addition to these “positive control” results, it is noteworthy that several other candidate compounds emerging from this analysis have converging mechanistic evidence of therapeutic potential. For example, the drug target enrichment analysis for MDD identified several proteins that are targeted by retinoic acid. It has recently been demonstrated that retinoic acid induces a signaling cascade that produces post-synaptic tuning effects in the hippocampus comparable to those elicited by ketamine (a rapidly-acting antidepressant)^50^. Moreover, Rai14, a molecule downstream of retinoic acid in that signaling pathway, was shown to stabilize and maintain mature dendritic spines, resulting in protection from depression-like behaviors^51^. Our druggability analysis (Supplementary Table 5) can also be utilized to identify potential psychiatric side effects of existing medications with non-psychiatric indications. For example, we confirm a prior report that inhibition of ACE (common in the treatment of hypertension) may be associated with increased risk for schizophrenia^52^.

## Limitations

While the present study was designed to understand the pathophysiology of brain-related phenotypes, our proteomic panel is derived from blood, rather than brain. Consequently, it is likely that we were unable to detect effects of nervous system proteins with limited expression in blood. In this context, however, it is noteworthy that several of our novel findings are proteins that are primarily expressed in brain, including NRCAM and NCAM1, two neural cell adhesion molecules. Moreover, because postmortem brain tissue is difficult to obtain, our reference panel is 1-2 orders of magnitude larger than those available, enhancing power for those proteins that are detectable in blood. While there may be tissue-specific differences in pQTL effects^53,54^, recent well-powered studies suggest that many QTLs are consistent across tissues^55,56^.

The control population utilized in the bipolar disorder GWAS cohort may have some overlap with the cohort used by deCODE to generate pQTL data. However, it is noteworthy that the majority of our findings were observed in the UKB-PPP cohort, which does not share any overlap with the Bipolar Disorder GWAS cohort. Specifically, out of the 61 (55 cis-pQTL associations, 6 trans-pQTL associations) significant proteins (FDR< 0.05), 28 proteins were assayed in both the deCODE and UKB-PPP cohort, with concordant results in both. Additionally, 20 of the 61 proteins were assayed exclusively in the UKB-PPP cohort; only 13 were exclusively identified in the deCODE pQTL study and could not be tested in UKB-PPP. The relatively high concordance between cis-pQTL MR findings in the UKB-PPP and deCODE datasets indicates that technical differences may not hinder meta-analytic studies across the SomaScan and Olink platforms. On the other hand, it is important to note that the MR results derived from trans-pQTL instruments were only weakly correlated across platforms, suggesting that caution should be applied in interpreting these results.

## Methods

### Overview

Our study utilizes protein quantitative trait loci (pQTL) derived from circulating plasma, drawn from two extensive resources: the UK Biobank Pharma Proteomics Project (UKB-PPP) and deCODE Genetics. These resources encompass the profiling of plasma pQTL data for 2,923 proteins from the UKB-PPP cohort that comprises data from 34,557 UK individuals of European ancestry and 4,719 aptamers from the deCODE cohort comprising 35,559 Icelandic individuals of European ancestry. GWAS summary statistics were obtained for European-ancestry subjects from the most recent Psychiatric Genomics Consortium (PGC) reports for SCZ, BIP, and MDD, as well as the largest available non-overlapping dataset for CTP (detailed below). Mendelian randomization was carried out to investigate the proteomic associations for each phenotype (Figure 1, Supplementary Figure 1). Finally, proteins that showed statistically significant causal effects from MR analysis were used for pathway, protein-protein interaction (PPI), and drug target enrichment analysis for each outcome phenotype separately.

## 1. Data Sources and Study Design

### 1.1 Exposure Data Selection

In this study, we employed protein quantitative trait loci (pQTL) data from two genome-wide association studies (GWAS) conducted among individuals of European descent to curate genetic instruments for Mendelian randomization analysis.

#### 1.1.1 UK Biobank Pharma Proteomics Project (UKB-PPP)

This project involves profiling plasma pQTL data for 2,923 proteins derived using the Olink Explore 3072 platform from a cohort of 34,557 UK Biobank participants^57^ of European ancestry.

#### 1.1.2 deCODE Genetics

In addition, we utilized plasma pQTL data obtained from deCODE Genetics, a study that includes 4,719 proteins measured across 35,559 Icelandic individuals. These proteins were quantified using the SOMAscan version 4 assay^58^.

### 1.2. Quality Control and processing of pQTL dataset

Summary statistics from UKB-PPP and deCODE genetics underwent a QC-filtering process, which involved the exclusion of insertion-deletion (INDELs) variants, variants with a minor allele frequency (MAF) below 0.001, palindromic variants with MAF exceeding 0.42, and, for trans-pQTLs, variants in the extended MHC region (Chr6:25-34Mb). QC-filtered summary statistics were then converted to GWAS VCF files, where the alternative allele denotes the effect allele, using the *gwas2vcf* tool (github.com/MRCIEU/gwas2vcf)^59^. After the QC-filtering step, we selected variants that achieve genome-wide significance (p > 5 × 10^−8^) for LD pruning to identify strong, independent genetic instruments. We first separated QC-filtered genome-wide significant variants into cis and trans-pQTLs, where cis-pQTLs are variants within 1 Mb upstream and downstream of the associated protein-coding genes (ensemble 108 annotations) and QC-filtered genome-wide significant variants outside of the cis region are considered trans-pQTLs. LD pruning was then performed for cis and trans-pQTLs separately to identify independent genetic instruments, using the LEUGWASR tool (Options-Clump_kb=10000, Clump_r2=0.01) with UKB-PPP (N=33,000) as LD reference panel (https://github.com/MRCIEU/ieugwasr).

Our MR instruments included a total of 17,317 cis-pQTLs linked to 2,063 unique proteins assessed within the UKB-PPP cohort (Supplementary Table 1a-b) as well as 23,350 cis-pQTLs related to 1,675 unique proteins measured in the deCODE cohort (Supplementary Table 1a-b). Although 847 proteins were assayed on both platforms, yielding a total of 2,891 unique proteins examined in MR using cis-pQTL instruments, we maintained the full denominator of 3,738 analyses for our Bonferroni correction to conservatively account for the fact that we took the lowest available p-value.

**Table 1:** Summary of Results. The following table summarizes the number of significant proteins identified for each of the four outcome phenotypes. While we have emphasized the highest confidence (Bonferroni-significant) results in the text above, we also present results at the FDR level of significance in the Table below, as well as Supplementary Tables 4c, 4d, 5c, and 5d.

| Phen<br>o | cis-pQTL<br>MR<br>results | cis-pQTL<br>Novel | Gene<br>sets<br>(# sets) | Drug<br>Targets<br>Enrichment<br>(# drugs) | Drug<br>Targets<br>(DGIdb) | Druggabl<br>e (DGIdb) | trans-pQTL MR<br>results | trans-pQTL<br>novel |
| --- | --- | --- | --- | --- | --- | --- | --- | --- |
| SCZ | 46 (132) | 9 (43) | 14<br>(132) | 186 (68) | 7 (33) | 37 (105) | 6 (21) | 6 (15) |
| BIP | 16 (55) | 3 (18) | 25 (17) | 190 (82) | 5 (18) | 13 (46) | 3 (6) | 3 (6) |
| MDD | 8 (13) | 7 (10) | 16 (30) | 54 (27) | 3 (3) | 8 (12) | 1 (1) | 1 (1) |
| CTP | 17 (74) | 9 (50) | 12 (61) | 59 (18) | 4 (19) | 13 (57) | 7 (28) | 6 (27) |

*Note:* Numbers in Table represent number of significant results at Bonferroni significance (number of results at FDR<0.05 in parentheses).

Separately, we utilized (as instruments for MR analysis) 31,709 trans-pQTL instruments (excluding the extended MHC region) corresponding to 2,543 unique proteins assessed within the UKB-PPP cohort (Supplementary Table 1-d), as well as 71,763 trans-pQTLs related to 4,268 unique proteins measured in the deCODE cohort (Supplementary Table 1c-d). Among these, 1,527 unique proteins were assayed by both platforms. Despite this overlap, we conservatively controlled for 6,811 tests in our Bonferroni corrections of the trans-pQTL MR results.

### 1.3. Outcome Data

#### 1.3.1. Psychiatric GWAS data

We have obtained the most recent large-scale meta-analyses of GWASs conducted by the Psychiatric Genomics Consortium (PGC) for three complex psychiatric disorders which include schizophrenia (N_case_ = 67,323, N_control_ = 93,456; file name: daner_PGC_SCZ_w3_90_0418b_ukbbdedupe.trios)^60^, bipolar disorder (N_case_ = 40,463, N_control_ = 313,436; file name: daner_bip_pgc3_nm_noukbiobank.gz)^61^, major depressive disorder (N_case_ = 166,773, N_control_ = 507,679; file name: daner_MDDwoBP_20201001_2015iR15iex_HRC_MDDwoBP_iPSYCH2015i_Wray_Fin nGen_MVPaf_2_HRC_MAF01.gz)^62^ as our outcome data for the MR analysis. The downloaded summary statistics for each disorder included only subjects of European ancestry. It excluded any subjects drawn from the UK Biobank to avoid any overlap with the cohort that used data from UKB-PPP. Additionally, the SCZ and MDD cohorts did not include subjects from overlapping with the deCODE pQTL cohort. However, the GWAS of bipolar disorder contained samples from deCODE (N_case_ = 1972, N_control_ = 192,602); summary statistics excluding these individuals were not available, and the extent of overlap of individuals with the deCODE pQTL cohort is not known.

#### 1.3.2. Cognitive Task Performance (CTP) GWAS data

Separately, for Cognitive Task Performance (CTP), we have recomputed GWAS for the largest available sample size of European ancestry subjects, excluding individuals in the UKB-PPP cohort. (It should be noted that this GWAS also contains no individuals from deCODE.) Specifically, we first accessed the genotypic data for “fluid intelligence” (Field ID: 20016) from the UK Biobank (UKB) cohort for all individuals of European ancestry, excluding those assayed on the proteomic platform of UKB-PPP (remaining N=141,123). Our analysis comprised the following key steps: initial sample and genotype quality control, excluding variants with genotyping call rates below 90%, those in Hardy-Weinberg disequilibrium (PHWE < 1 x 10^−1^^5^), and variants with a minor allele frequency <1%. We then performed a GWAS of the “fluid intelligence” phenotype using the regenie pipeline (version 3.2.4)^63^, with covariates including sex, mean age, age^2^, sex by age interaction, sex by age^2^ interaction, and the top 20 principal components.

Subsequently, we meta-analyzed these GWAS summary statistics from the cognitive GWAS of Savage et al. (2018)^64^, excluding all subjects from the UK Biobank (remaining N=74,210; file name: Cognition_Meta_GWAS_without_UKBPP1.tbl). GWAS meta-analysis was performed using METAL software^65^. The resulting GWAS summary statistics (N = 215,333) were employed as outcome data for the CTP trait in our MR analysis.

### 1.4. Processing the GWAS summary statistics

Summary statistics data of the three psychiatric GWASs and CTP meta-GWAS were converted to a GWAS VCF file using the gwas2vcf tool. To ensure consistency of genomic coordinates all the psychiatric GWAS summary statistics were converted into the GRCh38 build to match that of the pQTL data.

## 2. Mendelian randomization (MR) analysis

We performed MR analyses using the ‘MendelianRandomization’ package version 0.8.0^66^ ^67^ within the R environment. SNPs significantly associated with each protein (i.e., pQTLs) were used as instruments. The package includes approaches that compute Wald’s ratio for each SNP instrument’s effect on the outcome phenotype; when multiple SNP instruments are available for a given protein, these effects are combined using the delta-weighted inverse-variance method (IVW-Delta). We applied default parameters ‘MendelianRandomization’, where the function that estimates fixed-effects IVW-Delta was carried out only when fewer than four SNP instruments were available; otherwise, multiplicative random-effects IVW-Delta was employed. Random-effects meta-analysis is more conservative than fixed-effects meta-analysis in the presence of heterogeneity across instruments without sacrificing power in the absence of heterogeneity^68^. Moreover, multiplicative random-effect meta-analysis is more conservative in the presence of pleiotropy^34^ than an additive random-effects approach. (For comparison purposes and as a technical replication, we also performed MR analyses with standard fixed-effects IVW, using the commonly utilized TwoSampleMR package^17^; results of this comparison are available in the supplementary materials.) MR analyses were conducted separately for each pQTL dataset (UKB-PPP and deCODE). For the proteins that are present in both UKB-PPP and deCODE data, we conducted a p-value-based meta-analysis using Fisher’s method implemented in the metap package (https://github.com/cran/metap/blob/master/R/sumlog.R; sumlog function). Analyses were also conducted separately for cis-pQTLs and trans-pQTLs without extended MHC region (will be referred to as trans-pQTL in the later text).

### 2.1. Causal Direction, Heterogeneity, and Horizontal Pleiotropy Test

We have used MR Steiger test^17^ from the TwoSampleMR package^17^ to test for the appropriate causal direction (i.e., protein level causal to psychiatric or cognitive phenotype). To test the robustness and sensitivity of the result, we have used the MR-Egger intercept test^69^ from TwoSampleMR package^17^ to estimate horizontal pleiotropy and Cochran’s *Q* statistic derived from the IVW to estimate heterogeneity^70^.

## 3. Multiple Correction

We applied Bonferroni correction separately for cis-pQTL and trans-pQTL analyses within each phenotype. We further performed False Discovery Rate (FDR; Benjamini-Hochberg) correction on the lowest p-values obtained from the UKB-PPP, deCODE, or metap for cis- and trans-pQTL instruments, separately for all four phenotypes. Specifically, a total of 3,738 proteins had cis-pQTL instruments, and 6,809 proteins had trans-pQTL instruments available, resulting in a Bonferroni-corrected p-value (PBONF) of 1.3x10^-5^ (0.05/3,738) for Mendelian Randomization analysis of proteins using cis-pQTLs, and 7.3x10^-6^ (0.05/6,811) for MR analysis of proteins with trans-pQTLs.

## 4. Systematic literature review

To determine the novelty of our findings, we conducted a comprehensive and systematic literature review using the "advanced search" feature on PubMed, encompassing all outcome phenotypes considered in our study. Studies examining molecular phenotypes in relation to psychiatric GWAS have primarily focused on gene expression quantitative trait loci (eQTLs), so these were included in our search along with studies of pQTLs. (It should be noted that transcriptomic and proteomic data can often differ ^18^). Moreover, such studies have frequently examined brain-based molecular phenotypes, which have the advantage of direct assessment of neural tissue. However, brain-based studies have the disadvantage of relatively small sample sizes compared to our pQTL reference panels of ∼35,000 individuals each. Thus, our search strategy on PubMed was as follows:

Include the following terms in their title or abstracts: ("pqtl" or "pqtls" or "expression quantitative trait loci" or "protein quantitative trait loci" or "eqtl" or "eqtls" or "proteome" or "proteomics" or "proteomic" or "transcriptome" or "transcriptomic" or "transcriptomics" or "qtl" or "proteomes" or “transcriptomes" or "proteogenomic" or "expression") AND ("TWAS" or "transcriptome-wide association study" or "transcriptome wide association study" or "SMR" or "Summary data based mendelian randomization" or "mendelian randomization" or "MR" or "Mendelian randomisation" or "cis").

The initial search resulted in 43,226 publications found. We then refined our search within the 43,226 publications that include any one or more following terms in their title:

"schizophrenia" or "bipolar” or "depression" or "depressive" or "cognition" or "cognitive" or "intelligence" or "education" or "psychiatric" or "neurological" or "common disorders" or "common diseases" or "common trait" or "complex phenotypes" or "complex traits" or "complex trait" or "complex disorders" or "complex diseases" or "brain" or "neuropsychiatric".

This refinement resulted in 998 publications. We then removed any reviews, papers without human data, papers with data from induced pluripotent stem cells or organoids or tissues other than brain or blood, papers without available data, and preprints. 50 relevant publications remained - these selected articles were thoroughly reviewed to evaluate the novelty of our work. The novelty of our findings was evaluated using two distinct thresholds. Proteins deemed significant at the stringent Bonferroni level (P < 0.05) in our study were classified as novel if they lacked prior investigation or exhibited a P-value > 0.000015 or a PP4 interaction score less than 0.7 in the included literature. Similarly, proteins reaching significance at the less stringent 5% false discovery rate threshold (FDR < 0.05) in our study were categorized as novel if there were no no prior investigation on them or if they displayed a P-value > 0.001 or a PP4<0.7 within the included studies.

## 5. Pathway Enrichment

We performed gene-set enrichment analysis, examining only those proteins identified with the highest confidence level: specifically, proteins that were strictly significant (PBONF < 0.05) in the cis-pQTL analyses for each phenotype. We used a web-based tool, EnrichR^71^, specifically using its integrated gene-set libraries from KEGG, Reactome, MSigDB, and Gene Ontology (Cellular component, Molecular Function, and Biological Process).

## 6. Protein-protein Interaction

For these same Bonferroni-significant proteins emerging from the cis-pQTL analyses, a protein-protein interaction network for each outcome phenotype was constructed using STRINGDB (https://string-db.org/)^72^. This online tool incorporates a broad range of data sources to derive both known and predicted protein-protein interactions from both direct (physical) and indirect (functional) associations. Network images are derived using a spring model, with nodes modeled as masses and edges as springs; the final position of the nodes in the image is computed by minimizing the ’energy’ of the system.

## 7. Drug Target Enrichment

Further, for these same Bonferroni-significant proteins emerging from the cis-pQTL analyses, we performed a Drug Target enrichment analysis in EnrichR, specifically using its integrated libraries from DSigDB, DrugMatriX, and CMAP.

## 8. Gene-Drug Interaction and Potential Druggability

To further examine the druggability of these significant proteins emerging from Mendelian Randomization, we utilized DGIdb4.0^73^ (www.dgidb.org) to identify approved compounds, immunotherapies, and known chemical compounds that could interact with these proteins. Additionally, we used DGIdb4.0 to identify potential druggable genes that are known or predicted to interact with drugs. DGIdb is an online tool that uses a combination of expert curation and text-mining to extract drug-gene interactions from resources such as DrugBank, PharmGKB, ChEMBL, Drug Target Commons.

Furthermore, DGIdb can categorize genes as potentially druggable based on their presence in selected pathways, molecular functions, and gene families from the Gene Ontology, the Human Protein Atlas, IDG, "druggable genome" lists from Hopkins and Groom^74^ and Russ and Lampel^75^, among others.

## Data and Code Availability

1. Outcome GWAS summary statistics can be downloaded from SCZ: received from PGC consortium - will be made available upon publication BIP: https://Figureshare.com/ndownloader/files/40036705 MDD: https://ipsych.dk/fileadmin/ipsych.dk/Downloads/daner_MDDwoBP_20201001_2015iR15iex_HRC_MDDwoBP_iPSYCH2015i_Wray_FinnGen_MVPaf_2_HRC_MAF01.gz CTP: Received from UKBB - will be made available upon publication

1. Mendelian Randomization (MR) package (GitHub link: https://github.com/cran/MendelianRandomization )
2. TwoSampleMR package (GitHub link: https://mrcieu.github.io/TwoSampleMR/index.html )
3. Meta-analysis package (Github link: https://github.com/cran/metap/blob/master/R/sumlog.R)
4. EnricR (Webportal link: https://maayanlab.cloud/Enrichr/)
5. StringDB - Protein-Protein Interaction (Webportal link: https://string-db.org/ )
6. LEUGWASR package (Github link: https://github.com/MRCIEU/ieugwasr)

## Supporting information

High resolution Figures and Supplementary Figures

## Data Availability

All data produced in the present study are available upon reasonable request to the authors.

## Acknowledgements

The use of UKB data (WES, genome-wide genotyping and phenotypic data) in the current study is approved under application no. 26041. U.B., J.J., M.L., and T.L. were supported by the National Institute of Mental Health of the National Institutes of Health (NIH) under award no. R01MH117646 (T.L., principal investigator). The content is solely the responsibility of the authors and does not necessarily represent the official views of the NIH.

