## Supplementary material for "Large-Scale Mendelian Randomization Study Reveals Circulating Blood-based Proteomic Biomarkers for Psychopathology and Cognitive Task Performance": High resolution Figures and Supplementary Figures: Supplementary_Fig1_MR.pdf

### Exposure GWASs

1. UK Biobank Pharma Proteomics Project (UKB-PPP) (Sun et al, 2023)
2. deCODE (Ferkingstad et al, 2022)

Pre-Processing and QC

Conversion of summary statistics to VCF (Alt allele= effect allele)

Tool: gwas2vcf (github.com/MRCIEU/gwas2vcf)

Removal of
1.INDELs
2.Variants with MAF<0.001
3.Palindromic Variants (MAF> 0.42)
4. pQTLs with P>5E-8

Separating Cis and Trans pQTLS

pQTLS at 1 mb Upstream and Downstream of a Gene defined by ensemble 108

Remaining pQTLs without MHC (Chr6:25000000-34000000)

LD Pruning

*Tool: LEUGWASR; Options- Clump_kb=10000, Clump_r²=0.01, Reference Panel=Biogen UKBB cohort (N=50000)*

Cis pQTLs

Trans pQTLs without MHC

### Outcome GWASs

1. Schizophrenia (Trubetskoy et al, 2022)*#
2. Bipolar Disorder (Mullins et al, 2021)*
3. Major Depressive Disorder (Als et al, 2023)*#
4. Cognitive Task Performance (Savage et al. 2018, excluding UKB-PPP participants)#

*excluding UKB-PPP cohort #excluding deCODE cohort

Pre-Processing

Conversion of summary statistics to VCF → Conversion to GrcH38 build

Tool: gwas2vcf (github.com/MRCIEU/gwas2vcf)

Tool: bcftools (samtools.github.io/bcftools)

### MR Analysis

LD_Pruned UKBB-PPP Data (1.Cis, 2.Trans w/o MHC)

LD_Pruned deCODE data (1.Cis, 2. Trans w/o MHC)

*Tool: gwas2vcf (github/MRCIEU/gwasvcf/; Option: tag_r²=0.8)*

Proxy variant ($r^2$> 0.8) identification if variant in exposure GWAS not present in outcome GWAS

*Tool: TwoSampleMR (mrcieu.github.io/TwoSampleMR/; Option: harmonise_data)*

Harmonization of the exposure and outcome data

*github.com/sb452/MendelianRandomization*

*Mendelian Randomization* (IVW_Delta)

UKB-PPP_MR (*Cis, Trans w/o MHC)*

deCODE_MR (*Cis, Trans w/o MHC)*

### MR Sensitivity Analysis

MR results from UKBB-PPP & deCODE data (*Cis, Trans w/o MHC from each dataset)*

*Tool: TwoSampleMR Option: mr_heterogeneity)*

Heterogeneity

Horizontal Pleiotropy

### Meta Analysis

UKB-PPP_MR_*Cis* + deCODE_MR_*Cis*

UKB-PPP_MR_*Trans* + deCODE_MR_*Trans*

Meta Analysis

*Tool: metap cran.r-project.org/web/packages/metap/ Option: allmetap("sumlog")*

MR_*Cis_MetaP*

MR_Trans_*MetaP*

### Multiple Correction

Lowest P value from MR analysis using Cis-pQTLs (UKB-PPP/deCODE/METAP) for each outcome

Bonferroni (0.05/3738) & FDR (0.05*P value Rank/3738)

Lowest P value from MR analysis using deCODE-pQTLs (UKB-PPP/deCODE/METAP) for each outcome

Bonferroni (0.05/6811) & FDR (0.05*P value Rank/6811)

### Downstream Analyses

Proteins with Cis-Bpnf P <0.05 *from each outcome*

Pathway enrichment

*EnrichR*

Gene Ontology

Drug Targets

*StringDB*

PPI
