## Supplementary material for "Large-Scale Mendelian Randomization Study Reveals Circulating Blood-based Proteomic Biomarkers for Psychopathology and Cognitive Task Performance": High resolution Figures and Supplementary Figures: Supplementary_Fig_3A-BK_MR.pdf

**Supplementary Fig 3:** Forest plots showing distribution of beta values across pQTLs for the proteins that showed significant heterogeneity or horizontal pleiotropy

#### A. AIF1_SCZ_Cis-pQTL

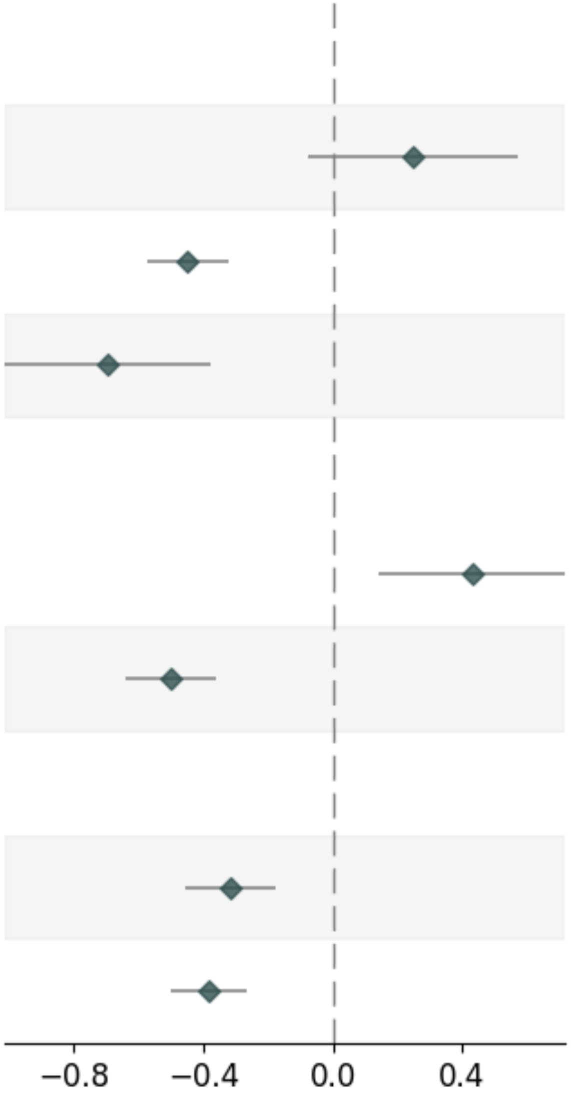


| Variable | Pvalue | Beta | Up_CI | Low_CI | nSNP | MAF |
|---|---|---|---|---|---|---|
| **deCODE_SingleSNP** | | | | | | |
| deCODE _6_31595147_C_T | 1.32e-01 | 0.247 | 0.569 | -0.074 | _ | 0.0766 |
| deCODE _6_31635814_A_G | 1.48e-12 | -0.447 | -0.323 | -0.57 | _ | 0.4514 |
| deCODE _6_32610037_G_T | 1.96e-05 | -0.694 | -0.375 | -1.012 | _ | 0.2627 |
| **UKB-PPP_SingleSNP** | | | | | | |
| UKB-PPP_6_31525184_A_G | 3.40e-03 | 0.43 | 0.718 | 0.142 | _ | 0.3482 |
| UKB-PPP_6_31635814_A_G | 1.48e-12 | -0.502 | -0.363 | -0.64 | _ | 0.4307 |
| **combined** | | | | | | |
| UKB-PPP | 7.63e-06 | -0.316 | -0.178 | -0.455 | 2 | _ |
| deCODE | 3.15e-10 | -0.382 | -0.263 | -0.501 | 3 | _ |

#### B. ARL3_SCZ_Cis-pQTL

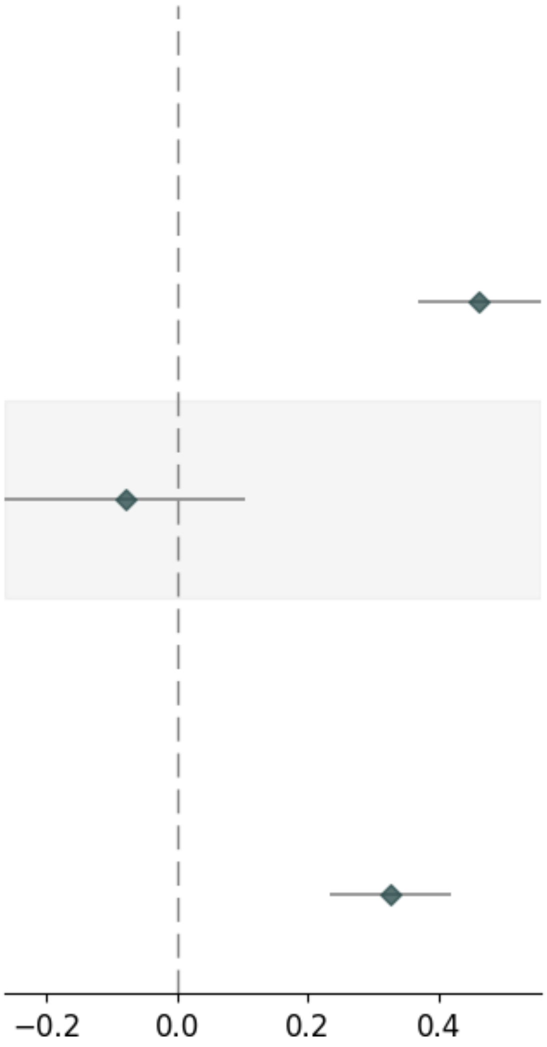


| Variable | | Pvalue | Beta | Up_CI | Low_CI | nSNP | MAF |
|---|---|---|---|---|---|---|---|
| **deCODE_SingleSNP** | | | | | | | |
| deCODE | 10_102676884_C_T | 9.47e-22 | 0.463 | 0.558 | 0.368 | _ | 0.1063 |
| deCODE | 10_102786426_A_G | 3.99e-01 | -0.079 | 0.105 | -0.264 | _ | 0.2869 |
| **combined** | | | | | | | |
| deCODE | | 5.60e-12 | 0.326 | 0.419 | 0.234 | 2 | _ |

**C. CGREF1_SCZ_Cis-pQTL**

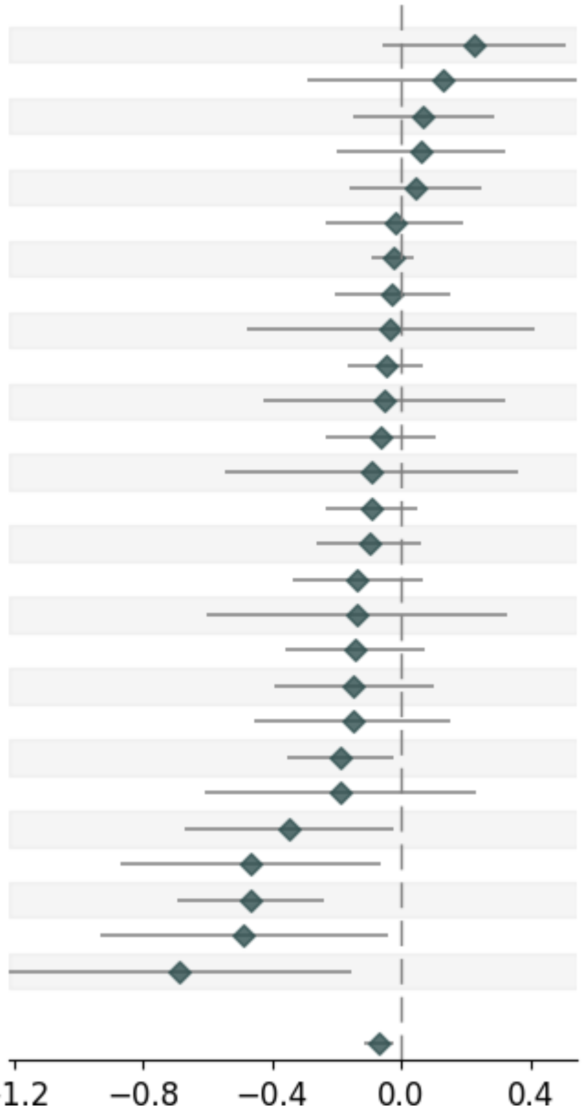


| Variable | Pvalue | Beta | Up_CI | Low_CI | nSNP | MAF |
|---|---|---|---|---|---|---|
| **UKB-PPP_SingleSNP** | | | | | | |
| UKB-PPP_2_27017106_A_T | 1.22e-01 | 0.223 | 0.506 | -0.06 | _ | 0.0105 |
| UKB-PPP_2_27070121_G_A | 5.51e-01 | 0.127 | 0.543 | -0.29 | _ | 0.0113 |
| UKB-PPP_2_26964963_T_C | 5.42e-01 | 0.068 | 0.289 | -0.152 | _ | 0.0138 |
| UKB-PPP_2_27446270_A_G | 6.55e-01 | 0.06 | 0.321 | -0.202 | _ | 0.015 |
| UKB-PPP_2_26916935_G_T | 6.74e-01 | 0.043 | 0.245 | -0.158 | _ | 0.0178 |
| UKB-PPP_2_26991122_T_C | 8.49e-01 | -0.021 | 0.192 | -0.233 | _ | 0.0133 |
| UKB-PPP_2_26715626_G_A | 3.93e-01 | -0.027 | 0.035 | -0.09 | _ | 0.0483 |
| UKB-PPP_2_27081683_A_G | 7.60e-01 | -0.028 | 0.15 | -0.206 | _ | 0.0142 |
| UKB-PPP_2_26873467_G_A | 8.75e-01 | -0.036 | 0.409 | -0.48 | _ | 0.0161 |
| UKB-PPP_2_26950446_G_A | 4.13e-01 | -0.048 | 0.067 | -0.164 | _ | 0.0432 |
| UKB-PPP_2_26809799_G_A | 7.87e-01 | -0.052 | 0.323 | -0.426 | _ | 0.0165 |
| UKB-PPP_2_26959446_G_A | 4.42e-01 | -0.066 | 0.103 | -0.235 | _ | 0.0156 |
| UKB-PPP_2_27247034_T_A | 6.90e-01 | -0.092 | 0.362 | -0.546 | _ | 0.0203 |
| UKB-PPP_2_27011881_G_A | 2.04e-01 | -0.093 | 0.051 | -0.237 | _ | 0.0243 |
| UKB-PPP_2_26879679_T_C | 2.27e-01 | -0.101 | 0.063 | -0.264 | _ | 0.0198 |
| UKB-PPP_2_27171912_C_T | 1.82e-01 | -0.136 | 0.064 | -0.335 | _ | 0.0204 |
| UKB-PPP_2_27142724_G_T | 5.58e-01 | -0.139 | 0.326 | -0.605 | _ | 0.0254 |
| UKB-PPP_2_27252791_G_A | 1.98e-01 | -0.142 | 0.074 | -0.357 | _ | 0.0111 |
| UKB-PPP_2_27223831_G_A | 2.41e-01 | -0.148 | 0.099 | -0.395 | _ | 0.0122 |
| UKB-PPP_2_27116290_G_T | 3.33e-01 | -0.15 | 0.153 | -0.453 | _ | 0.0194 |
| UKB-PPP_2_27067098_A_C | 2.60e-02 | -0.187 | -0.022 | -0.352 | _ | 0.0188 |
| UKB-PPP_2_26767213_A_G | 3.73e-01 | -0.191 | 0.229 | -0.61 | _ | 0.0244 |
| UKB-PPP_2_27070173_C_T | 3.47e-02 | -0.346 | -0.025 | -0.668 | _ | 0.0194 |
| UKB-PPP_2_26920408_A_G | 2.33e-02 | -0.465 | -0.063 | -0.866 | _ | 0.0141 |
| UKB-PPP_2_27660372_C_T | 5.25e-05 | -0.465 | -0.24 | -0.691 | _ | 0.0469 |
| UKB-PPP_2_27796957_G_A | 3.24e-02 | -0.487 | -0.041 | -0.932 | _ | 0.0205 |
| UKB-PPP_2_27137905_G_A | 1.10e-02 | -0.685 | -0.157 | -1.213 | _ | 0.0138 |
| **combined** | | | | | | |
| UKB-PPP | 1.72e-03 | -0.071 | -0.027 | -0.115 | 27 | _ |

#### D. CD14_SCZ_Cis-pQTL

| Variable | Pvalue | Beta | Up_CI | Low_CI | nSNP | MAF |
|---|---|---|---|---|---|---|
| **deCODE_SingleSNP** | | | | | | |
| deCODE _5_141287515_G_A | 2.29e-01 | 0.157 | 0.413 | -0.099 | _ | 0.0375 |
| deCODE _5_140633982_T_G | 3.29e-04 | -0.132 | -0.06 | -0.205 | _ | 0.3074 |
| deCODE _5_140936673_A_G | 2.67e-01 | -0.181 | 0.139 | -0.502 | _ | 0.0157 |
| deCODE _5_139853212_G_A | 1.93e-01 | -0.236 | 0.12 | -0.593 | _ | 0.0486 |
| **UKB-PPP_SingleSNP** | | | | | | |
| UKB-PPP_5_141067739_T_C | 6.76e-03 | 0.454 | 0.783 | 0.126 | _ | 0.0519 |
| UKB-PPP_5_140360869_C_T | 1.47e-05 | -0.145 | -0.08 | -0.211 | _ | 0.2335 |
| UKB-PPP_5_141476782_A_G | 2.53e-01 | -0.26 | 0.186 | -0.706 | _ | 0.0148 |
| **combined** | | | | | | |
| deCODE | 8.06e-03 | -0.118 | -0.031 | -0.206 | 4 | _ |
| UKB-PPP | 1.07e-04 | -0.128 | -0.063 | -0.192 | 3 | _ |

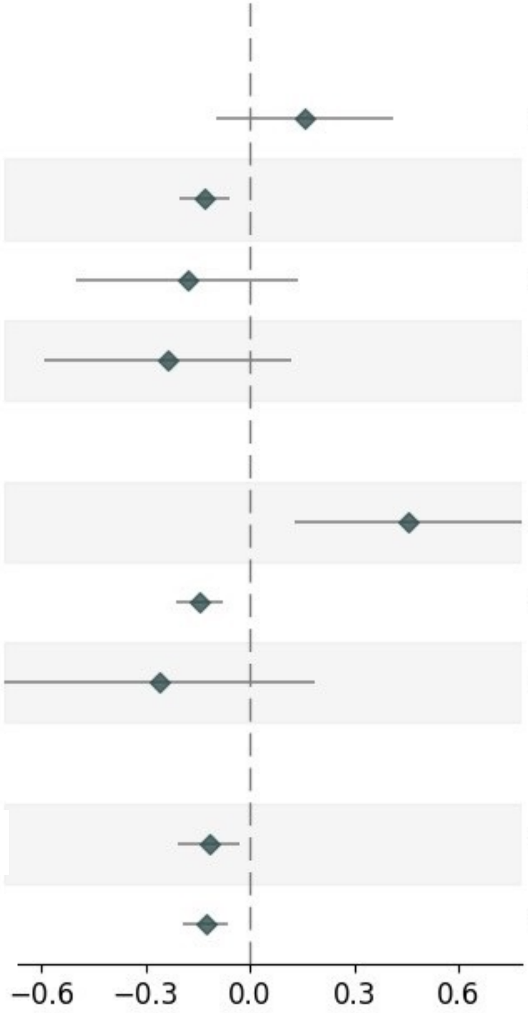

### E. CFI_SCZ_Cis-pQTL

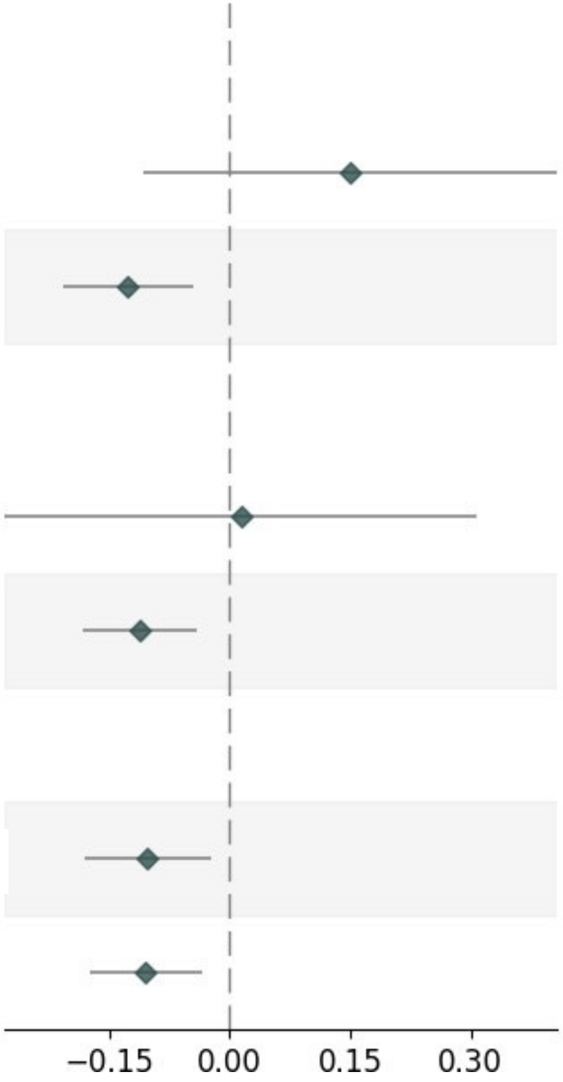


| Variable | Pvalue | Beta | Up_CI | Low_CI | nSNP | MAF |
|---|---|---|---|---|---|---|
| **deCODE_SingleSNP** | | | | | | |
| deCODE 4_109827389_T_A | 2.53e-01 | 0.15 | 0.408 | -0.107 | _ | 0.1467 |
| deCODE 4_109737911_T_C | 2.10e-03 | -0.127 | -0.046 | -0.208 | _ | 0.4827 |
| **UKB-PPP_SingleSNP** | | | | | | |
| UKB-PPP_4_110045298_A_G | 9.23e-01 | 0.014 | 0.308 | -0.28 | _ | 0.0392 |
| UKB-PPP_4_109737911_T_C | 2.10e-03 | -0.112 | -0.041 | -0.184 | _ | 0.4837 |
| **combined** | | | | | | |
| deCODE | 1.00e-02 | -0.102 | -0.024 | -0.18 | 2 | _ |
| UKB-PPP | 3.19e-03 | -0.105 | -0.035 | -0.175 | 2 | _ |

### F. CTSS_SCZ_Cis-pQTL

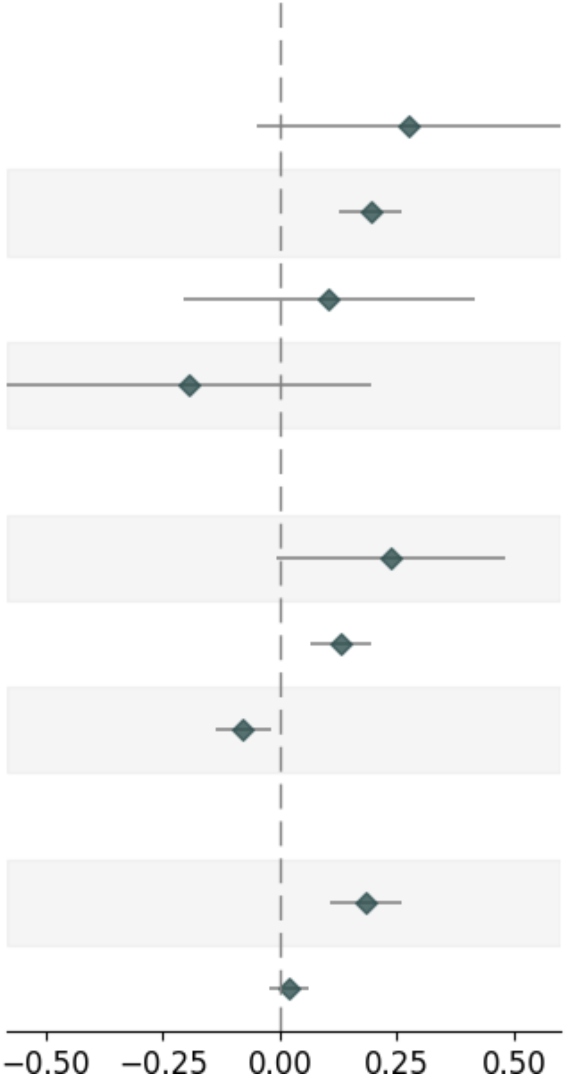


| Variable | Pvalue | Beta | Up_CI | Low_CI | nSNP | MAF |
|---|---|---|---|---|---|---|
| **deCODE_SingleSNP** | | | | | | |
| deCODE _1_150846616_T_A | 9.75e-02 | 0.275 | 0.601 | -0.05 | _ | 0.0591 |
| deCODE _1_150050850_G_A | 1.73e-08 | 0.195 | 0.262 | 0.127 | _ | 0.0821 |
| deCODE _1_151722519_C_T | 5.12e-01 | 0.104 | 0.416 | -0.207 | _ | 0.1066 |
| deCODE _1_150006012_C_T | 3.33e-01 | -0.192 | 0.197 | -0.582 | _ | 0.0685 |
| **UKB-PPP_SingleSNP** | | | | | | |
| UKB-PPP_1_151378584_G_A | 5.48e-02 | 0.238 | 0.48 | -0.005 | _ | 0.129 |
| UKB-PPP_1_150312297_G_T | 9.23e-05 | 0.13 | 0.195 | 0.065 | _ | 0.1237 |
| UKB-PPP_1_150644223_A_G | 8.17e-03 | -0.078 | -0.02 | -0.135 | _ | 0.4045 |
| **combined** | | | | | | |
| deCODE | 2.30e-06 | 0.183 | 0.259 | 0.107 | 4 | _ |
| UKB-PPP | 3.69e-01 | 0.02 | 0.062 | -0.023 | 3 | _ |

#### G. CXCL14_SCZ_Cis-pQTL

| Variable | Pvalue | Beta | Up_CI | Low_CI | nSNP | MAF |
|---|---|---|---|---|---|---|
| **UKB-PPP_SingleSNP** | | | | | | |
| UKB-PPP_5_135608457_A_G | 4.82e-02 | -0.22 | -0.002 | -0.438 | _ | 0.3573 |
| UKB-PPP_5_135694015_G_A | 3.26e-06 | -0.798 | -0.462 | -1.134 | _ | 0.2319 |
| **combined** | | | | | | |
| UKB-PPP | 3.27e-04 | -0.357 | -0.162 | -0.551 | 2 | _ |

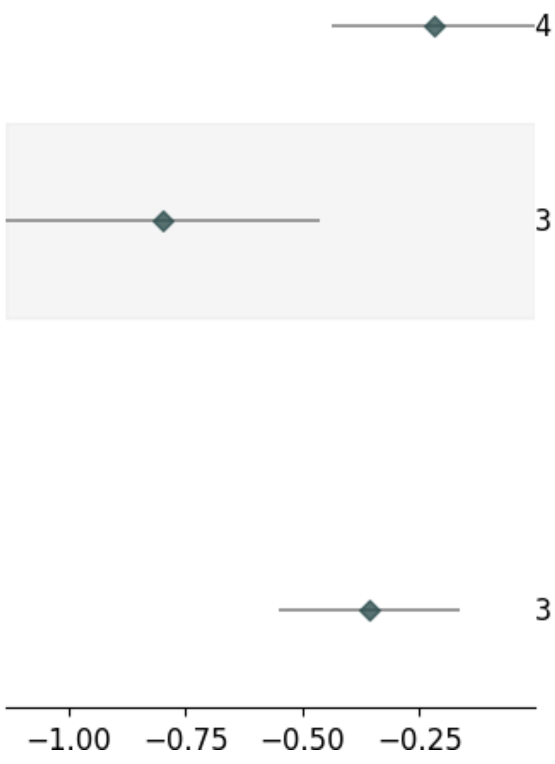

#### H. DPEP1_SCZ_Cis-pQTL

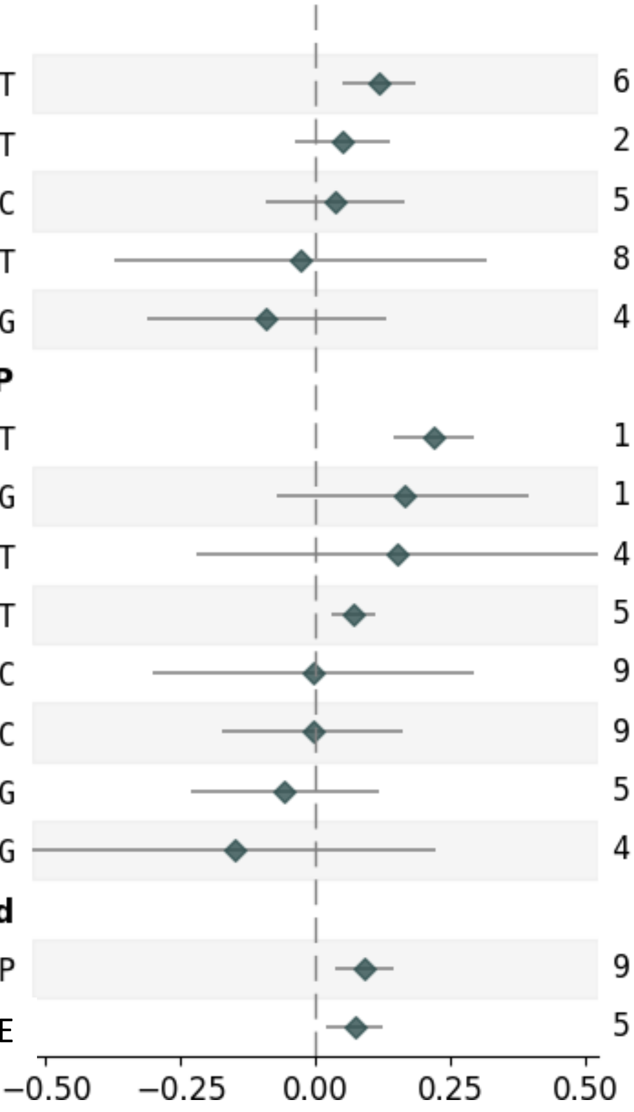


| Variable | Pvalue | Beta | Up_CI | Low_CI | nSNP | MAF |
|---|---|---|---|---|---|---|
| **deCODE_SingleSNP** | | | | | | |
| deCODE_16_89936274_C_T | 6.39e-04 | 0.119 | 0.187 | 0.051 | _ | 0.3865 |
| deCODE_16_89614569_C_T | 2.68e-01 | 0.049 | 0.137 | -0.038 | _ | 0.2105 |
| deCODE_16_89317317_T_C | 5.76e-01 | 0.036 | 0.164 | -0.091 | _ | 0.3437 |
| deCODE_16_88922955_G_T | 8.77e-01 | -0.027 | 0.319 | -0.374 | _ | 0.0076 |
| deCODE_16_88731220_A_G | 4.20e-01 | -0.091 | 0.13 | -0.313 | _ | 0.2386 |
| **UKB-PPP_SingleSNP** | | | | | | |
| UKB-PPP_16_89808220_C_T | 1.02e-08 | 0.22 | 0.295 | 0.145 | _ | 0.4198 |
| UKB-PPP_16_90104403_A_G | 1.70e-01 | 0.164 | 0.397 | -0.07 | _ | 0.0725 |
| UKB-PPP_16_88924495_C_T | 4.28e-01 | 0.151 | 0.525 | -0.222 | _ | 0.0246 |
| UKB-PPP_16_89579615_C_T | 5.23e-04 | 0.071 | 0.111 | 0.031 | _ | 0.2029 |
| UKB-PPP_16_88624200_A_C | 9.80e-01 | -0.004 | 0.294 | -0.302 | _ | 0.0838 |
| UKB-PPP_16_89220830_T_C | 9.53e-01 | -0.005 | 0.163 | -0.173 | _ | 0.0729 |
| UKB-PPP_16_89817719_A_G | 5.15e-01 | -0.058 | 0.117 | -0.232 | _ | 0.0259 |
| UKB-PPP_16_89428268_A_G | 4.28e-01 | -0.151 | 0.222 | -0.524 | _ | 0.0119 |
| **combined** | | | | | | |
| UKB-PPP | 9.03e-04 | 0.091 | 0.145 | 0.037 | 8 | _ |
| deCODE | 5.80e-03 | 0.074 | 0.126 | 0.021 | 5 | _ |

#### I. GPHA2_SCZ_Cis-pQTL

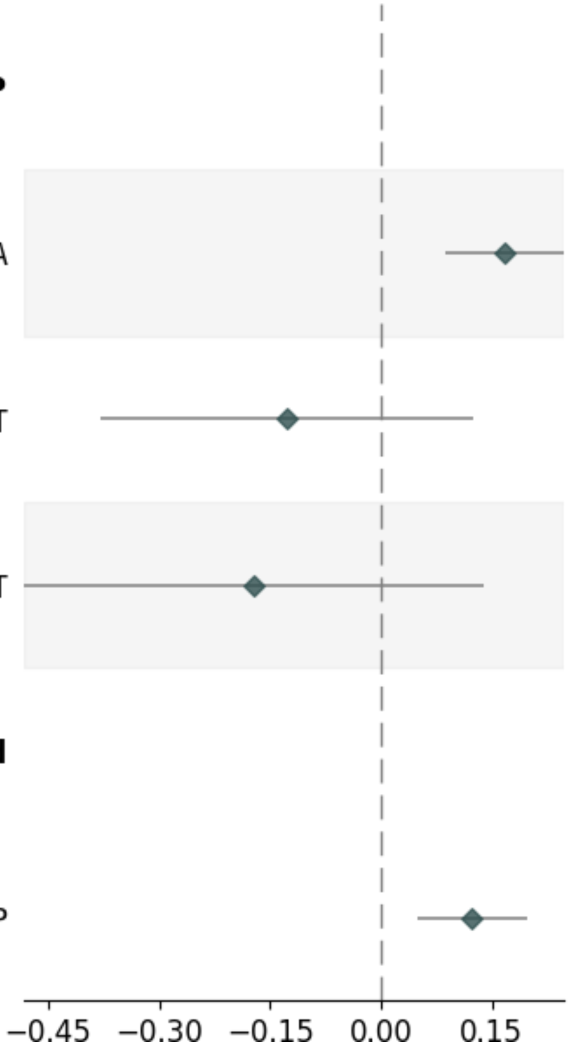


| Variable | Pvalue | Beta | Up_CI | Low_CI | nSNP | MAF |
|---|---|---|---|---|---|---|
| **UKB-PPP_SingleSNP** | | | | | | |
| UKB-PPP_11_64962847_C_A | 3.66e-05 | 0.168 | 0.248 | 0.088 | _ | 0.0461 |
| UKB-PPP_11_64963297_C_T | 3.26e-01 | -0.126 | 0.126 | -0.379 | _ | 0.0158 |
| UKB-PPP_11_65052392_C_T | 2.79e-01 | -0.171 | 0.139 | -0.482 | _ | 0.0253 |
| **combined** | | | | | | |
| UKB-PPP | 1.20e-03 | 0.123 | 0.198 | 0.049 | 3 | _ |

#### J. HLA-E_SCZ_Cis-pQTL

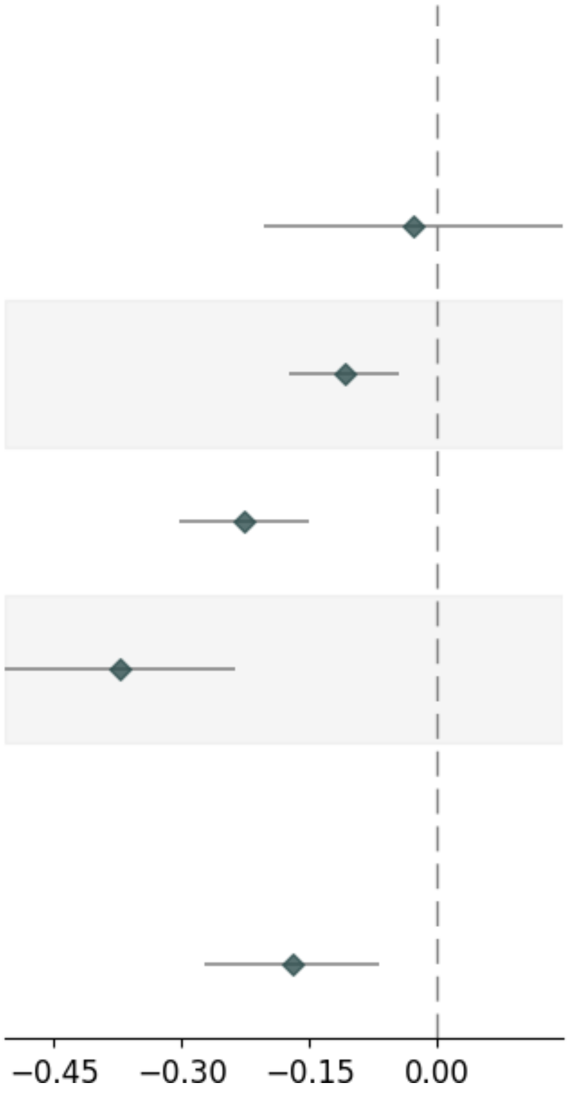


| Variable | Pvalue | Beta | Up_CI | Low_CI | nSNP | MAF |
|---|---|---|---|---|---|---|
| **UKB-PPP_SingleSNP** | | | | | | |
| UKB-PPP_6_31294684_T_C | 7.58e-01 | -0.028 | 0.148 | -0.204 | _ | 0.068 |
| UKB-PPP_6_31270982_C_T | 8.84e-04 | -0.109 | -0.045 | -0.173 | _ | 0.4994 |
| UKB-PPP_6_31225746_C_T | 5.55e-09 | -0.227 | -0.151 | -0.303 | _ | 0.3523 |
| UKB-PPP_6_31389166_A_T | 6.60e-08 | -0.372 | -0.237 | -0.507 | _ | 0.1906 |
| **combined** | | | | | | |
| UKB-PPP | 1.28e-03 | -0.17 | -0.067 | -0.273 | 4 | _ |

#### K. ITGAL_SCZ_Cis-pQTL

| Variable | Pvalue | Beta | Up_CI | Low_CI | nSNP | MAF |
|---|---|---|---|---|---|---|
| **UKB-PPP_SingleSNP** | | | | | | |
| UKB-PPP_16_30471219_T_G | 2.63e-02 | -0.122 | -0.014 | -0.23 | _ | 0.4844 |
| UKB-PPP_16_29967744_A_G | 7.57e-10 | -1.013 | -0.69 | -1.336 | _ | 0.4911 |
| **combined** | | | | | | |
| UKB-PPP | 1.70e-03 | -0.169 | -0.063 | -0.275 | 2 | _ |

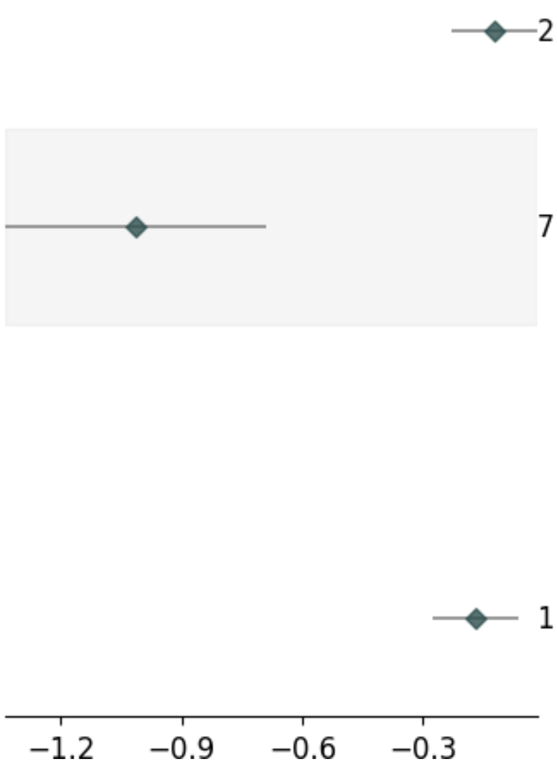

**L. ITIH1_SCZ_Cis-pQTL**

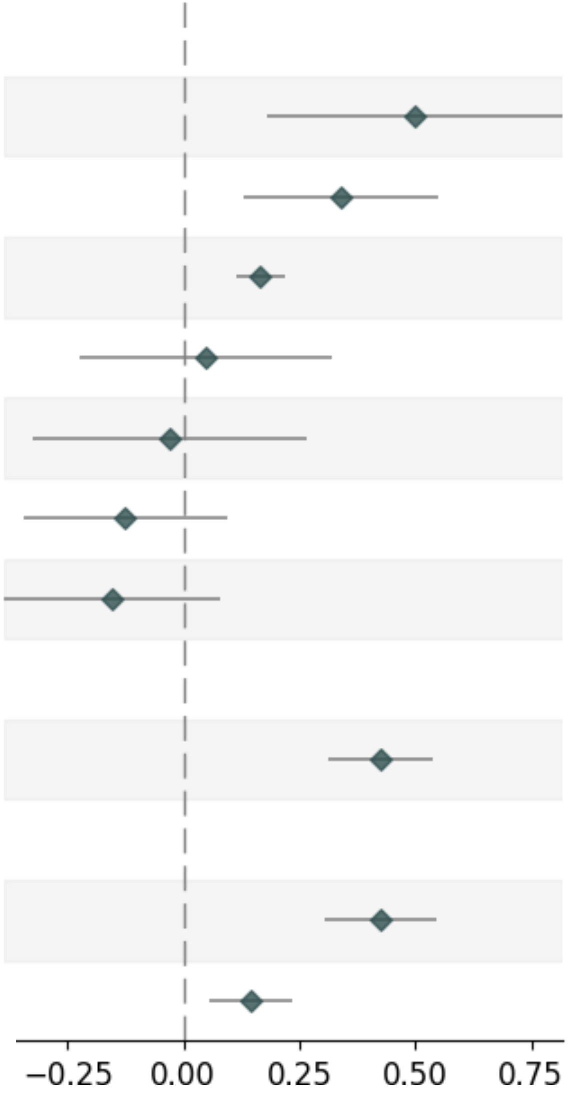


| Variable | Pvalue | Beta | Up_CI | Low_CI | nSNP | MAF |
|---|---|---|---|---|---|---|
| **deCODE_SingleSNP** | | | | | | |
| deCODE _3_53178767_C_T | 2.22e-03 | 0.499 | 0.818 | 0.179 | _ | 0.0173 |
| deCODE _3_52741044_C_T | 1.54e-03 | 0.339 | 0.548 | 0.129 | _ | 0.0683 |
| deCODE _3_52180624_A_G | 6.57e-10 | 0.165 | 0.218 | 0.113 | _ | 0.3635 |
| deCODE _3_53014314_A_G | 7.29e-01 | 0.048 | 0.32 | -0.224 | _ | 0.0189 |
| deCODE _3_53210133_G_A | 8.50e-01 | -0.028 | 0.267 | -0.324 | _ | 0.0101 |
| deCODE _3_53739410_T_C | 2.65e-01 | -0.125 | 0.095 | -0.344 | _ | 0.1557 |
| deCODE _3_51837747_C_T | 1.96e-01 | -0.154 | 0.079 | -0.386 | _ | 0.0101 |
| **UKB-PPP_SingleSNP** | | | | | | |
| UKB-PPP_3_52786965_A_T | 8.48e-14 | 0.426 | 0.538 | 0.314 | _ | 0.3572 |
| **combined** | | | | | | |
| UKB-PPP | 6.36e-12 | 0.426 | 0.547 | 0.304 | 1 | _ |
| deCODE | 1.26e-03 | 0.146 | 0.234 | 0.057 | 7 | _ |

#### M. TIH3_SCZ_Cis-pQTL

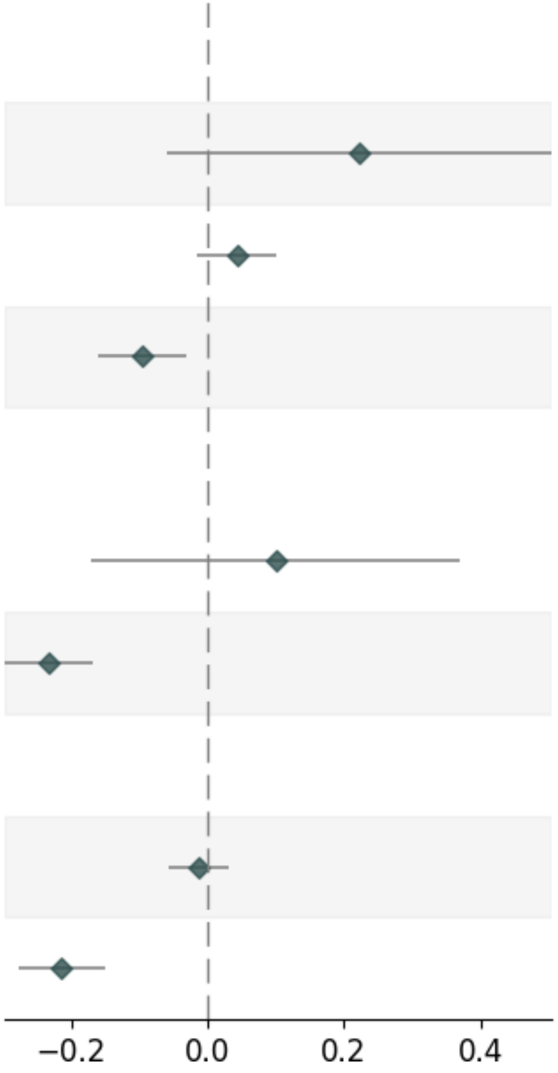


| Variable | Pvalue | Beta | Up_CI | Low_CI | nSNP | MAF |
|---|---|---|---|---|---|---|
| **Decode_SingleSNP** | | | | | | |
| Decode_3_53738091_A_G | 1.21e-01 | 0.223 | 0.505 | -0.059 | _ | 0.0344 |
| Decode_3_53676323_T_C | 1.52e-01 | 0.043 | 0.102 | -0.016 | _ | 0.029 |
| Decode_3_52814191_C_T | 4.43e-03 | -0.095 | -0.03 | -0.16 | _ | 0.0872 |
| **UKB-PPP_SingleSNP** | | | | | | |
| UKB-PPP_3_53157050_C_A | 4.70e-01 | 0.1 | 0.37 | -0.171 | _ | 0.0401 |
| UKB-PPP_3_52243429_A_G | 2.15e-12 | -0.233 | -0.168 | -0.297 | _ | 0.3174 |
| **combined** | | | | | | |
| Decode | 5.54e-01 | -0.013 | 0.03 | -0.057 | 3 | _ |
| UKB-PPP | 8.20e-11 | -0.214 | -0.149 | -0.278 | 2 | _ |

### N. ITIH4_SCZ_Cis-pQTL

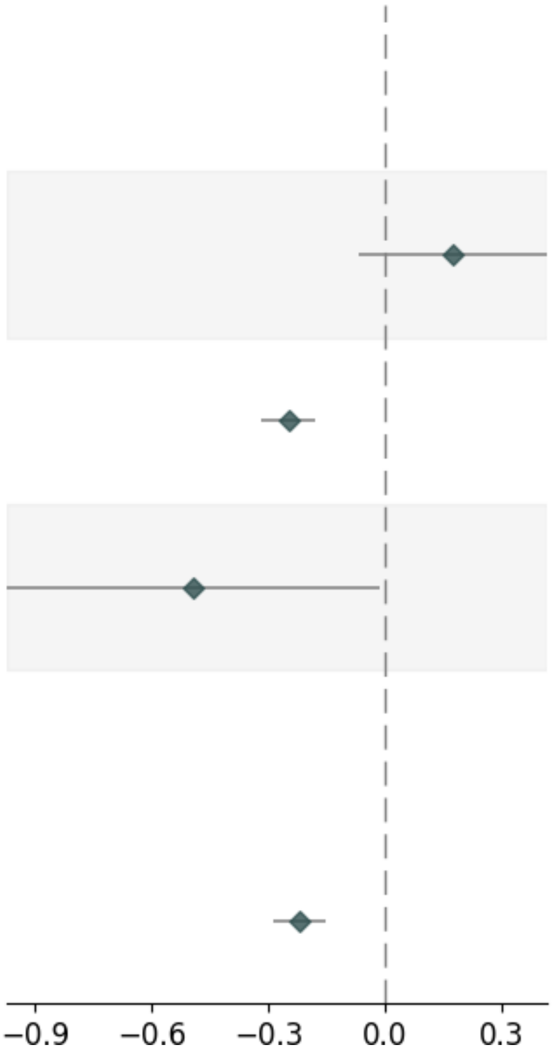


| Variable | Pvalue | Beta | Up_CI | Low_CI | nSNP | MAF |
|---|---|---|---|---|---|---|
| **UKB-PPP_SingleSNP** | | | | | | |
| UKB-PPP_3_53029815_G_A | 1.58e-01 | 0.174 | 0.416 | -0.068 | _ | 0.0106 |
| UKB-PPP_3_52785311_T_A | 3.67e-12 | -0.249 | -0.179 | -0.319 | _ | 0.3953 |
| UKB-PPP_3_53007170_T_C | 4.28e-02 | -0.494 | -0.016 | -0.972 | _ | 0.0184 |
| **combined** | | | | | | |
| UKB-PPP | 2.97e-10 | -0.22 | -0.152 | -0.289 | 3 | _ |

#### O. NCR3LG1_SCZ_Cis-pQTL

| Variable | Pvalue | Beta | Up_CI | Low_CI | nSNP | MAF |
|---|---|---|---|---|---|---|
| **UKB-PPP_SingleSNP** | | | | | | |
| UKB-PPP_11_17379587_G_A | 4.74e-02 | 0.124 | 0.247 | 0.001 | _ | 0.0802 |
| UKB-PPP_11_17017480_T_C | 8.33e-04 | 0.119 | 0.189 | 0.049 | _ | 0.4581 |
| UKB-PPP_11_17517860_A_T | 1.34e-01 | -0.139 | 0.043 | -0.321 | _ | 0.291 |
| **combined** | | | | | | |
| UKB-PPP | 1.40e-03 | 0.095 | 0.153 | 0.037 | 3 | _ |

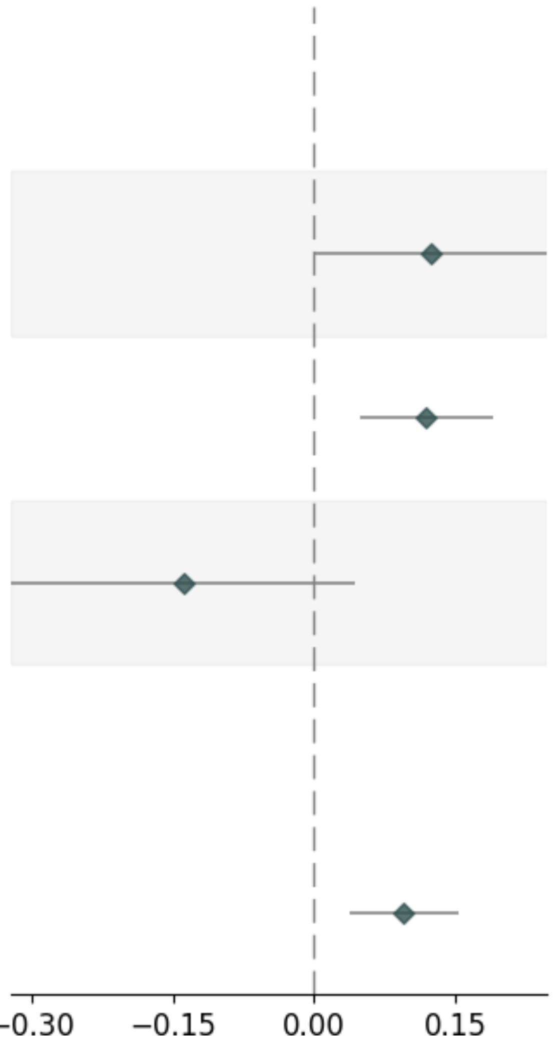

#### P. NMB_SCZ_Cis-pQTL

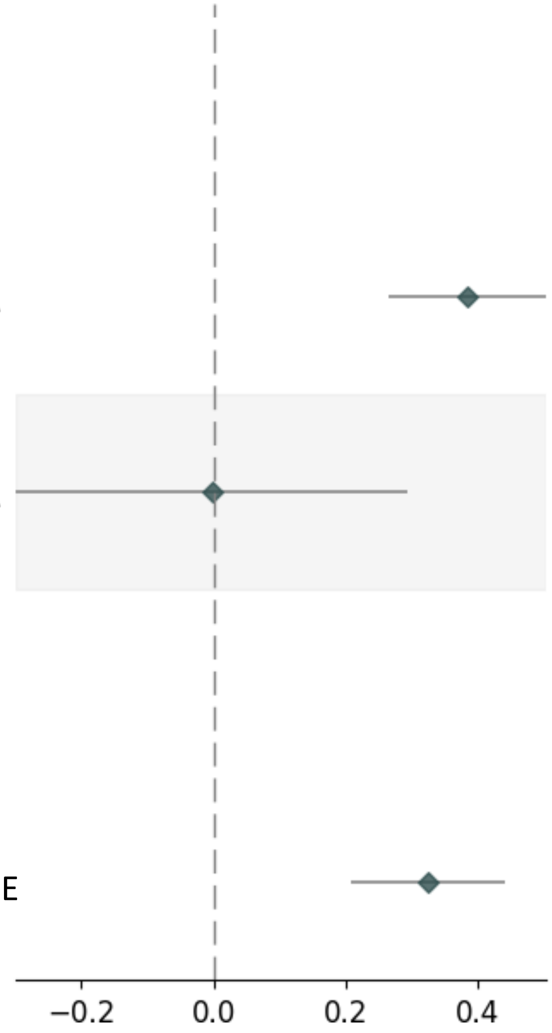


| Variable | Pvalue | Beta | Up_CI | Low_CI | nSNP | MAF |
|---|---|---|---|---|---|---|
| **deCODE_SingleSNP** | | | | | | |
| deCODE _15_84761911_T_A | 2.28e-10 | 0.384 | 0.503 | 0.266 | _ | 0.3321 |
| deCODE _15_83683730_G_A | 9.88e-01 | -0.002 | 0.294 | -0.298 | _ | 0.1301 |
| **combined** | | | | | | |
| deCODE | 5.58e-08 | 0.324 | 0.441 | 0.207 | 2 | _ |

### Q. SERPING1_SCZ_Cis-pQTL

| Variable | Pvalue | Beta | Up_CI | Low_CI | nSNP | MAF |
|---|---|---|---|---|---|---|
| **deCODE_SingleSNP** | | | | | | |
| deCODE_11_57070939_G_A | 9.25e-01 | 0.009 | 0.186 | -0.169 | _ | 0.0388 |
| deCODE_11_57467314_G_A | 1.28e-02 | -0.069 | -0.015 | -0.123 | _ | 0.1098 |
| deCODE_11_58031711_C_T | 1.83e-04 | -0.166 | -0.079 | -0.252 | _ | 0.2454 |
| **UKB-PPP_SingleSNP** | | | | | | |
| UKB-PPP_11_57598250_T_C | 6.00e-02 | 0.097 | 0.197 | -0.004 | _ | 0.049 |
| UKB-PPP_11_57653985_C_A | 1.81e-07 | -0.278 | -0.174 | -0.383 | _ | 0.3238 |
| **combined** | | | | | | |
| UKB-PPP | 3.65e-02 | -0.079 | -0.005 | -0.152 | 2 | _ |
| deCODE | 9.98e-05 | -0.089 | -0.044 | -0.134 | 3 | _ |

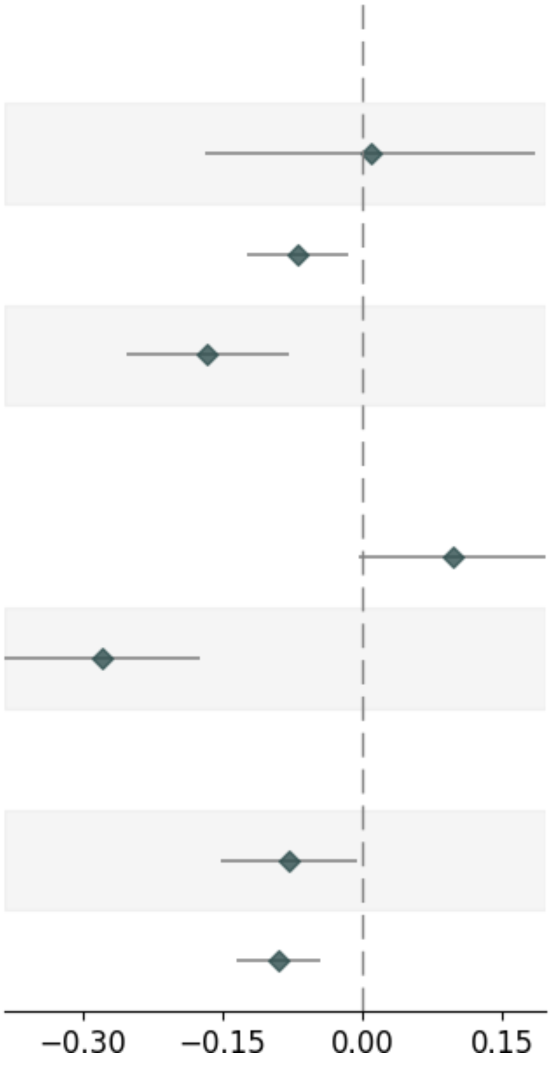

#### R. SIRPA_SCZ_Cis-pQTL (UKB-PPP)

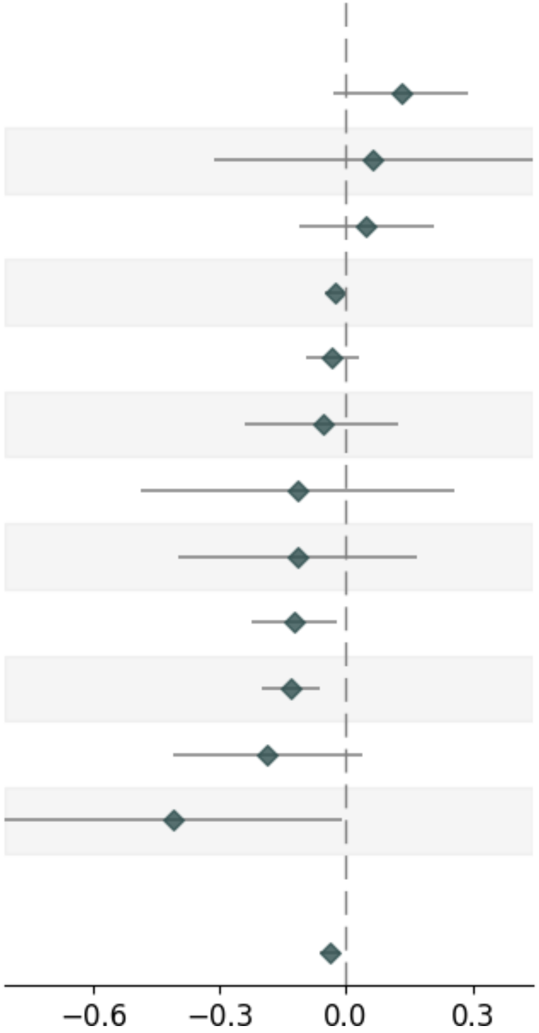


| Variable | Pvalue | Beta | Up_CI | Low_CI | nSNP | MAF |
|---|---|---|---|---|---|---|
| **UKB-PPP_SingleSNP** | | | | | | |
| UKB-PPP_20_1930044_G_A | 1.11e-01 | 0.13 | 0.291 | -0.03 | _ | 0.0478 |
| UKB-PPP_20_1194130_G_T | 7.37e-01 | 0.065 | 0.444 | -0.314 | _ | 0.1827 |
| UKB-PPP_20_1803759_G_A | 5.47e-01 | 0.049 | 0.21 | -0.112 | _ | 0.2437 |
| UKB-PPP_20_1902308_T_C | 1.45e-02 | -0.027 | -0.005 | -0.049 | _ | 0.1767 |
| UKB-PPP_20_1888701_T_C | 3.23e-01 | -0.032 | 0.031 | -0.095 | _ | 0.2753 |
| UKB-PPP_20_2050237_C_T | 5.43e-01 | -0.056 | 0.125 | -0.238 | _ | 0.0431 |
| UKB-PPP_20_1986052_T_C | 5.47e-01 | -0.114 | 0.258 | -0.487 | _ | 0.0137 |
| UKB-PPP_20_1998527_A_C | 4.24e-01 | -0.115 | 0.167 | -0.396 | _ | 0.0185 |
| UKB-PPP_20_1570839_C_T | 1.84e-02 | -0.121 | -0.02 | -0.222 | _ | 0.3543 |
| UKB-PPP_20_1815815_G_T | 1.66e-04 | -0.13 | -0.062 | -0.198 | _ | 0.4105 |
| UKB-PPP_20_2026077_G_A | 1.01e-01 | -0.187 | 0.037 | -0.41 | _ | 0.012 |
| UKB-PPP_20_2179590_C_T | 4.49e-02 | -0.409 | -0.009 | -0.809 | _ | 0.0157 |
| **combined** | | | | | | |
| UKB-PPP | 4.42e-03 | -0.038 | -0.012 | -0.064 | 12 | _ |

### S. SIRPA_SCZ_Cis-pQTL(DECODE)

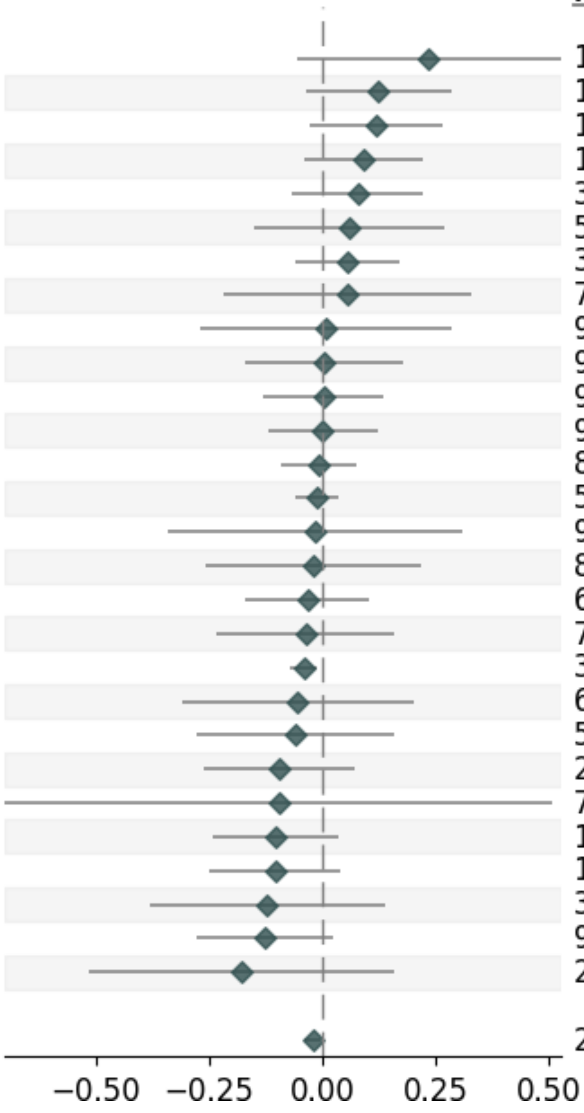


| Variable | Pvalue | Beta | Up_CI | Low_CI | nSNP | MAF |
|---|---|---|---|---|---|---|
| **deCODE_SingleSNP** | | | | | | |
| deCODE_20_1017639_C_T | 1.13e-01 | 0.235 | 0.527 | -0.056 | _ | 0.0112 |
| deCODE_20_1142143_A_G | 1.32e-01 | 0.123 | 0.283 | -0.037 | _ | 0.053 |
| deCODE_20_1930044_G_A | 1.11e-01 | 0.118 | 0.263 | -0.027 | _ | 0.044 |
| deCODE_20_1405219_T_C | 1.78e-01 | 0.091 | 0.223 | -0.041 | _ | 0.273 |
| deCODE_20_2307823_C_T | 3.03e-01 | 0.077 | 0.223 | -0.069 | _ | 0.1343 |
| deCODE_20_1631194_C_T | 5.79e-01 | 0.06 | 0.27 | -0.151 | _ | 0.0042 |
| deCODE_20_2009971_T_C | 3.38e-01 | 0.056 | 0.171 | -0.059 | _ | 0.0198 |
| deCODE_20_1031106_G_A | 7.03e-01 | 0.053 | 0.327 | -0.22 | _ | 0.0199 |
| deCODEe_20_920028_G_A | 9.50e-01 | 0.009 | 0.286 | -0.268 | _ | 0.0267 |
| deCODE_20_2771148_G_T | 9.71e-01 | 0.003 | 0.177 | -0.17 | _ | 0.0603 |
| deCODE_20_1934491_A_G | 9.71e-01 | 0.002 | 0.136 | -0.131 | _ | 0.0076 |
| deCODE_20_1871454_C_A | 9.82e-01 | 0.001 | 0.121 | -0.119 | _ | 0.0774 |
| deCODE_20_1795547_T_C | 8.40e-01 | -0.009 | 0.075 | -0.092 | _ | 0.1765 |
| deCODE_20_1936563_G_A | 5.82e-01 | -0.013 | 0.034 | -0.061 | _ | 0.0679 |
| deCODE_20_976467_A_C | 9.19e-01 | -0.017 | 0.307 | -0.341 | _ | 0.0268 |
| deCODE_20_1927548_G_A | 8.72e-01 | -0.019 | 0.218 | -0.257 | _ | 0.0072 |
| deCODE_20_2009783_T_G | 6.28e-01 | -0.034 | 0.104 | -0.172 | _ | 0.0284 |
| deCODE_20_2114071_A_G | 7.00e-01 | -0.038 | 0.157 | -0.234 | _ | 0.019 |
| deCODE_20_1899904_C_T | 3.40e-03 | -0.042 | -0.014 | -0.07 | _ | 0.1041 |
| deCODE_20_2656919_G_A | 6.79e-01 | -0.054 | 0.201 | -0.309 | _ | 0.0104 |
| deCODE_20_2052585_C_T | 5.87e-01 | -0.06 | 0.157 | -0.277 | _ | 0.0123 |
| deCODE_20_2591187_A_G | 2.60e-01 | -0.096 | 0.071 | -0.263 | _ | 0.0332 |
| deCODE_20_2271504_T_A | 7.54e-01 | -0.097 | 0.508 | -0.701 | _ | 0.026 |
| deCODE_20_2335283_C_T | 1.50e-01 | -0.103 | 0.037 | -0.243 | _ | 0.4422 |
| deCODE_20_1190465_A_G | 1.57e-01 | -0.105 | 0.04 | -0.249 | _ | 0.0919 |
| deCODE_20_900450_G_A | 3.54e-01 | -0.122 | 0.137 | -0.381 | _ | 0.0862 |
| deCODE_20_971192_C_T | 9.45e-02 | -0.128 | 0.022 | -0.277 | _ | 0.0176 |
| deCODE_20_894452_C_T | 2.94e-01 | -0.18 | 0.157 | -0.517 | _ | 0.0275 |
| **combined** | | | | | | |
| deCODE | 2.28e-02 | -0.022 | -0.003 | -0.042 | 28 | _ |

### T. VSIG2_SCZ_Cis-pQTL

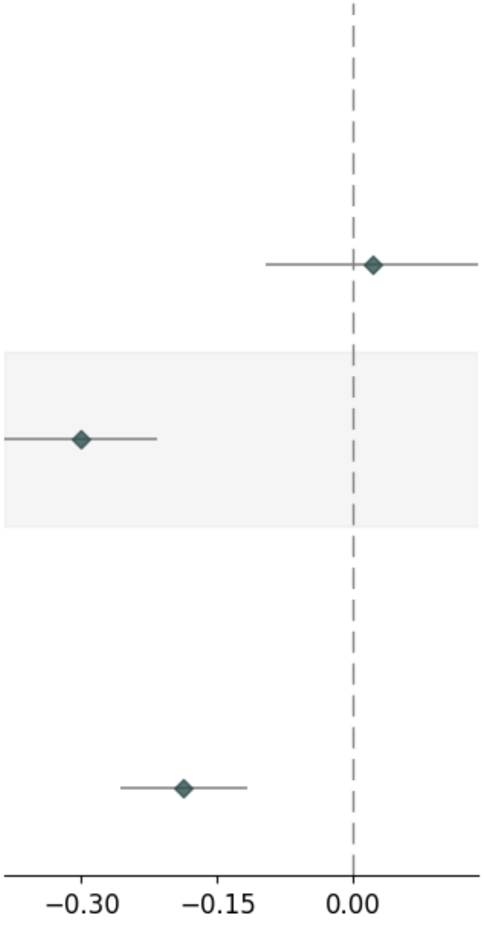


| Variable | Pvalue | Beta | Up_CI | Low_CI | nSNP | MAF |
|---|---|---|---|---|---|---|
| **UKB-PPP_SingleSNP** | | | | | | |
| UKB-PPP_11_124731504_A_G | 7.29e-01 | 0.021 | 0.138 | -0.097 | _ | 0.0398 |
| UKB-PPP_11_124750251_G_T | 1.87e-12 | -0.301 | -0.217 | -0.384 | _ | 0.2559 |
| **combined** | | | | | | |
| UKB-PPP | 1.50e-07 | -0.187 | -0.117 | -0.257 | 2 | _ |

#### U. CTSS_BIP_Cis-pQTL

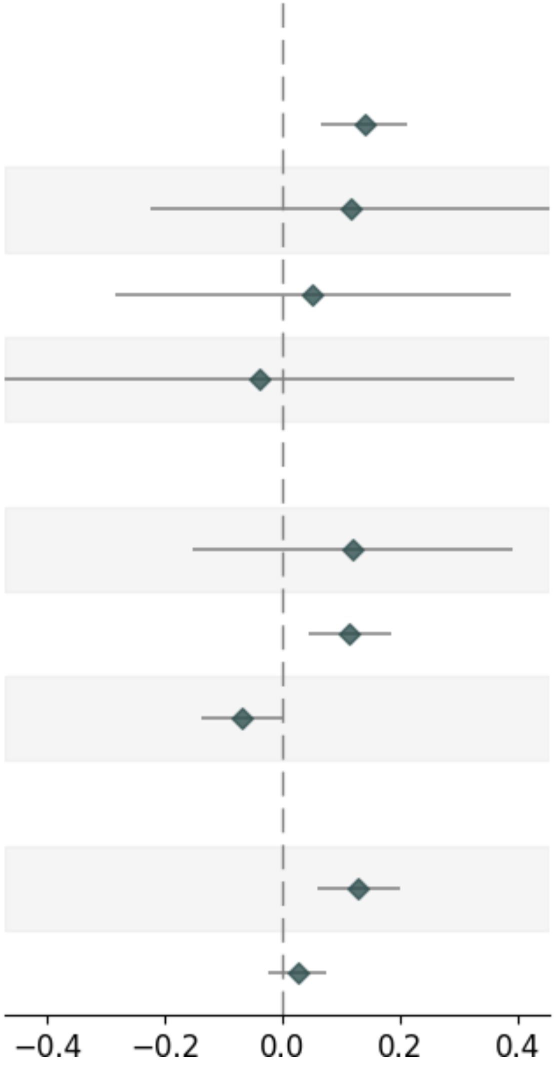


| Variable | Pvalue | Beta | Up_CI | Low_CI | nSNP | MAF |
|---|---|---|---|---|---|---|
| **deCODE_SingleSNP** | | | | | | |
| deCODE_1_150050850_G_A | 2.17e-04 | 0.14 | 0.214 | 0.066 | _ | 0.0821 |
| deCODE_1_151722519_C_T | 5.01e-01 | 0.116 | 0.456 | -0.223 | _ | 0.1066 |
| deCODE_1_150846616_T_A | 7.63e-01 | 0.052 | 0.388 | -0.284 | _ | 0.0591 |
| deCODE_1_150006012_C_T | 8.58e-01 | -0.039 | 0.394 | -0.472 | _ | 0.0685 |
| **UKB-PPP_SingleSNP** | | | | | | |
| UKB-PPP_1_151378584_G_A | 3.87e-01 | 0.12 | 0.393 | -0.152 | _ | 0.129 |
| UKB-PPP_1_150312297_G_T | 1.14e-03 | 0.115 | 0.185 | 0.046 | _ | 0.1237 |
| UKB-PPP_1_150644223_A_G | 5.28e-02 | -0.069 | 0.001 | -0.138 | _ | 0.4045 |
| **combined** | | | | | | |
| deCODE | 2.75e-04 | 0.13 | 0.2 | 0.06 | 4 | _ |
| UKB-PPP | 2.96e-01 | 0.026 | 0.074 | -0.023 | 3 | _ |

#### V. ITIH4_BIP_Cis-pQTL

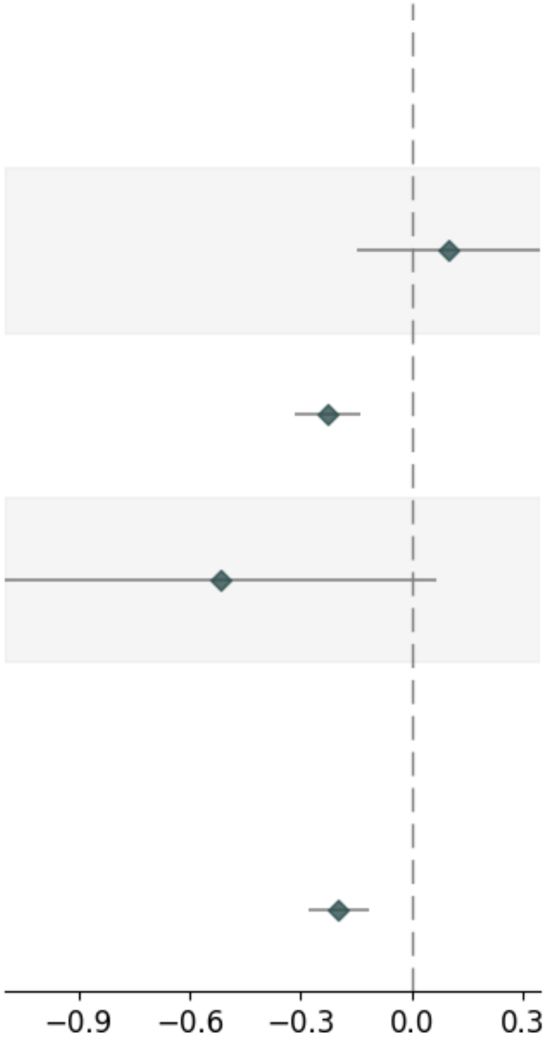


| Variable | Pvalue | Beta | Up_CI | Low_CI | nSNP | MAF |
|---|---|---|---|---|---|---|
| **UKB-PPP_SingleSNP** | | | | | | |
| UKB-PPP_3_53029815_G_A | 4.37e-01 | 0.099 | 0.347 | -0.15 | _ | 0.0106 |
| UKB-PPP_3_52785311_T_A | 3.58e-07 | -0.228 | -0.14 | -0.316 | _ | 0.3953 |
| UKB-PPP_3_53007170_T_C | 8.33e-02 | -0.514 | 0.068 | -1.097 | _ | 0.0184 |
| **combined** | | | | | | |
| UKB-PPP | 3.32e-06 | -0.197 | -0.114 | -0.28 | 3 | _ |

**W. MAPK3_BIP _Cis-pQTL**

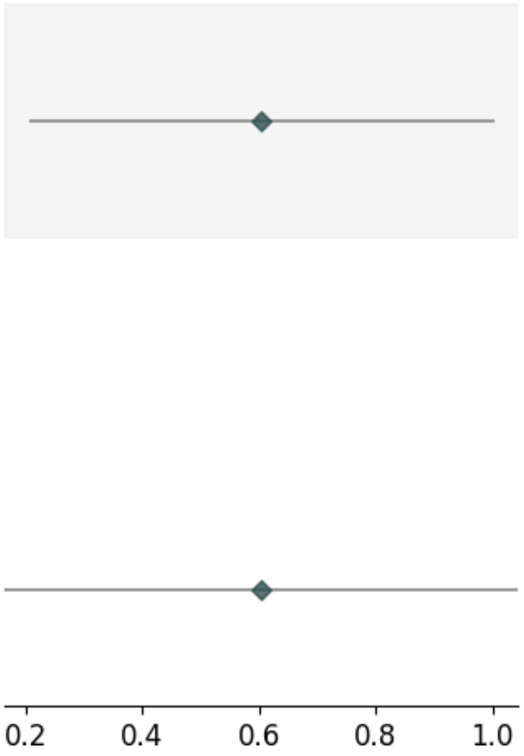


| Variable | Pvalue | Beta | Up_CI | Low_CI | nSNP | MAF |
|---|---|---|---|---|---|---|
| **deCODE_SingleSNP** | | | | | | |
| deCODE_14_103541163_C_T | 3.02e-03 | 0.604 | 1.003 | 0.205 | _ | 0.2806 |
| **combined** | | | | | | |
| deCODE | 7.17e-03 | 0.604 | 1.044 | 0.164 | 1 | _ |

### X.MLN_BIP_Cis-pQTL

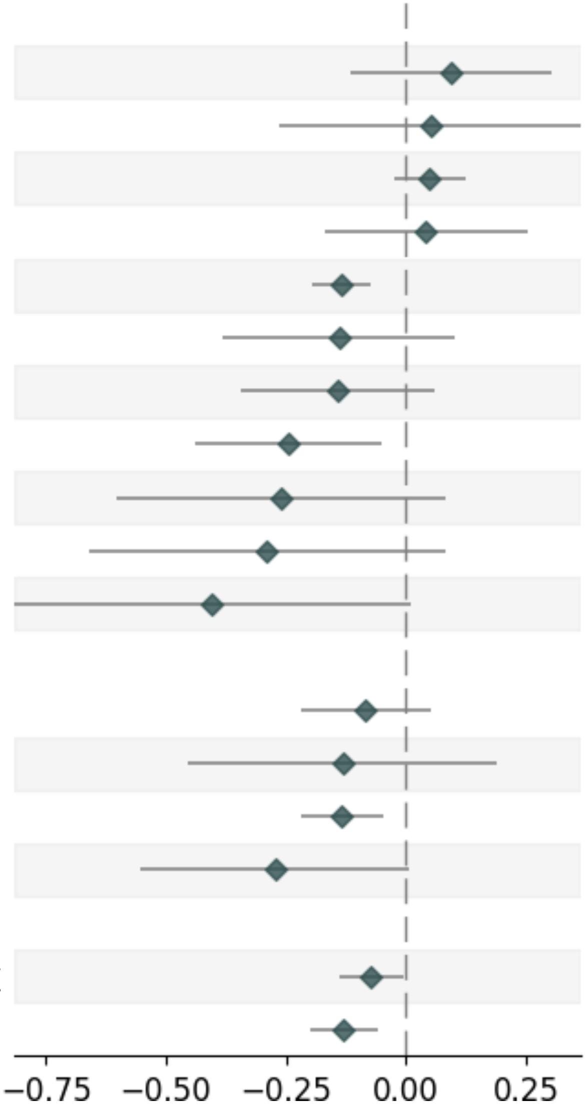


| Variable | Pvalue | Beta | Up_CI | Low_CI | nSNP | MAF |
|---|---|---|---|---|---|---|
| **deCODE_SingleSNP** | | | | | | |
| deCODE_6_32822282_T_C | 3.77e-01 | 0.094 | 0.302 | -0.114 | _ | 0.0272 |
| deCODE_6_34269213_G_A | 7.49e-01 | 0.051 | 0.364 | -0.262 | _ | 0.0361 |
| deCODE_6_33823453_C_T | 2.03e-01 | 0.048 | 0.123 | -0.026 | _ | 0.1939 |
| deCODE_6_33041397_A_T | 7.01e-01 | 0.041 | 0.252 | -0.17 | _ | 0.3113 |
| deCODE_6_33682966_A_G | 1.07e-05 | -0.135 | -0.075 | -0.196 | _ | 0.1857 |
| deCODE_6_34070252_T_C | 2.54e-01 | -0.14 | 0.1 | -0.38 | _ | 0.0182 |
| deCODE_6_32836604_A_T | 1.68e-01 | -0.141 | 0.059 | -0.342 | _ | 0.0602 |
| deCODE_6_33947994_C_T | 1.34e-02 | -0.244 | -0.051 | -0.437 | _ | 0.4638 |
| deCODE_6_32962902_G_A | 1.37e-01 | -0.26 | 0.083 | -0.602 | _ | 0.0719 |
| deCODE_6_33111758_C_T | 1.26e-01 | -0.289 | 0.081 | -0.659 | _ | 0.0309 |
| deCODE_6_32925384_G_A | 5.51e-02 | -0.403 | 0.009 | -0.814 | _ | 0.0177 |
| **UKB-PPP_SingleSNP** | | | | | | |
| UKB-PPP_6_33760579_G_A | 2.27e-01 | -0.084 | 0.052 | -0.22 | _ | 0.0342 |
| UKB-PPP_6_33606457_G_A | 4.19e-01 | -0.132 | 0.189 | -0.454 | _ | 0.4352 |
| UKB-PPP_6_33689073_C_T | 2.35e-03 | -0.133 | -0.047 | -0.219 | _ | 0.4487 |
| UKB-PPP_6_33902029_G_A | 5.43e-02 | -0.273 | 0.005 | -0.552 | _ | 0.345 |
| **combined** | | | | | | |
| deCODE | 3.53e-02 | -0.072 | -0.005 | -0.139 | 11 | _ |
| UKB-PPP | 2.51e-04 | -0.129 | -0.06 | -0.198 | 4 | _ |

#### Y. NCAM1_BIP_Cis-pQTL

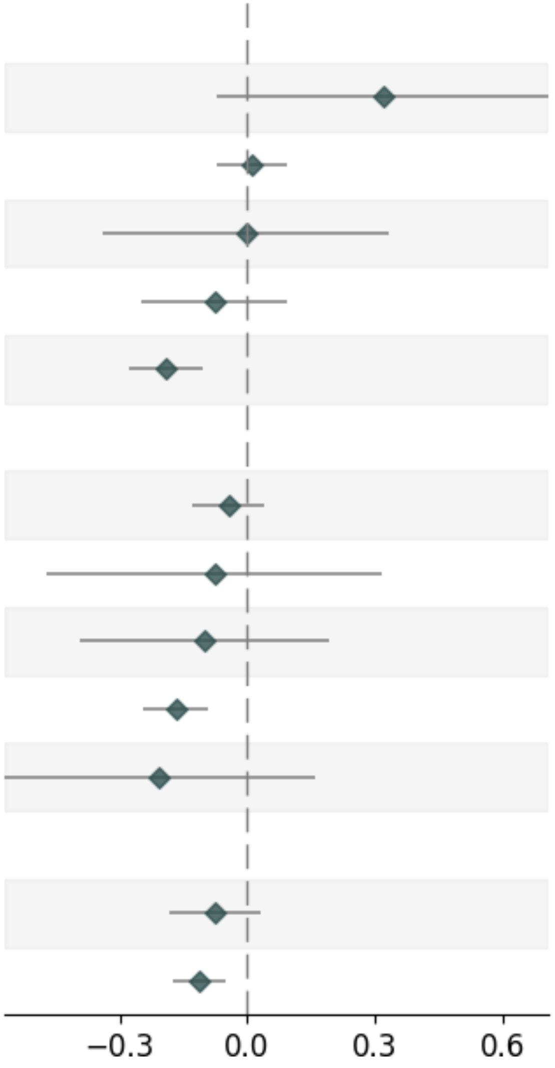


| Variable | Pvalue | Beta | Up_CI | Low_CI | nSNP | MAF |
|---|---|---|---|---|---|---|
| **deCODE_SingleSNP** | | | | | | |
| Decode_11_113649477_C_T | 1.10e-01 | 0.318 | 0.708 | -0.072 | _ | 0.0136 |
| Decode_11_113262954_T_G | 8.24e-01 | 0.009 | 0.093 | -0.074 | _ | 0.1301 |
| Decode_11_113248134_C_T | 9.84e-01 | -0.004 | 0.333 | -0.34 | _ | 0.4001 |
| Decode_11_112772253_C_T | 3.65e-01 | -0.078 | 0.091 | -0.248 | _ | 0.472 |
| Decode_11_113105212_C_T | 1.03e-05 | -0.192 | -0.106 | -0.277 | _ | 0.1876 |
| **UKB-PPP_SingleSNP** | | | | | | |
| UKB-PPP_11_112768683_T_C | 3.12e-01 | -0.044 | 0.041 | -0.129 | _ | 0.4877 |
| UKB-PPP_11_113580333_C_T | 7.00e-01 | -0.077 | 0.316 | -0.471 | _ | 0.0854 |
| UKB-PPP_11_113264096_C_T | 5.00e-01 | -0.101 | 0.192 | -0.394 | _ | 0.0611 |
| UKB-PPP_11_113044643_T_C | 1.84e-05 | -0.168 | -0.091 | -0.244 | _ | 0.2004 |
| UKB-PPP_11_112570055_C_T | 2.65e-01 | -0.206 | 0.157 | -0.569 | _ | 0.4047 |
| **combined** | | | | | | |
| Decode | 1.58e-01 | -0.077 | 0.03 | -0.183 | 5 | _ |
| UKB-PPP | 2.26e-04 | -0.113 | -0.053 | -0.173 | 5 | _ |

### Z. APOC1_CTP_Cis-pQTL

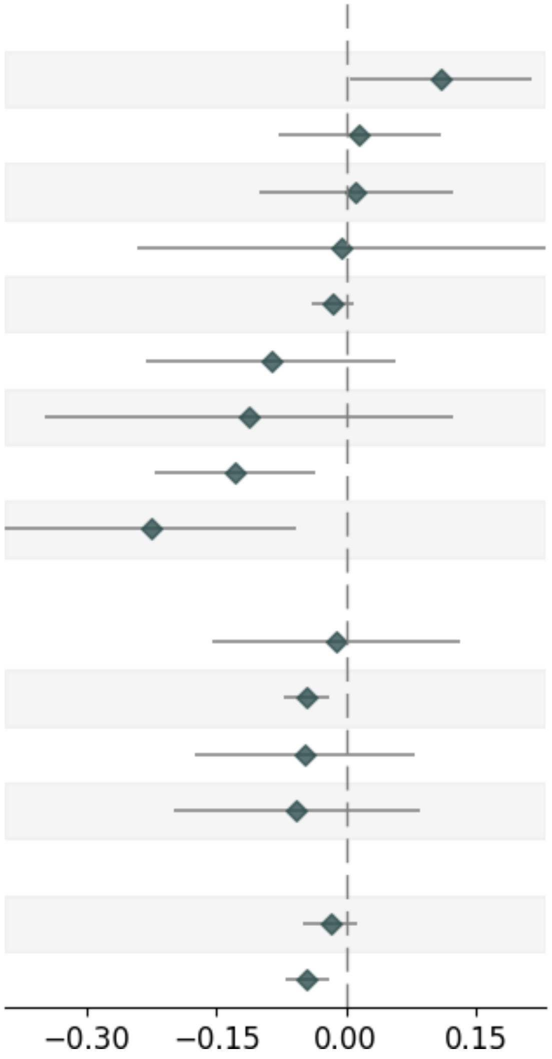


| Variable | Pvalue | Beta | Up_CI | Low_CI | nSNP | MAF |
|---|---|---|---|---|---|---|
| **deCODE_SingleSNP** | | | | | | |
| deCODE_19_45232745_T_C | 4.05e-02 | 0.109 | 0.214 | 0.005 | _ | 0.367 |
| deCODE_19_45175587_G_T | 7.49e-01 | 0.015 | 0.109 | -0.079 | _ | 0.0121 |
| deCODE_19_44753944_G_T | 8.49e-01 | 0.011 | 0.123 | -0.101 | _ | 0.0512 |
| deCODE_19_43966527_C_T | 9.69e-01 | -0.005 | 0.231 | -0.241 | _ | 0.0158 |
| deCODE_19_44928401_A_G | 2.20e-01 | -0.015 | 0.009 | -0.039 | _ | 0.4648 |
| deCODE_19_44890259_C_T | 2.36e-01 | -0.087 | 0.057 | -0.231 | _ | 0.0143 |
| deCODE_19_44441276_G_A | 3.51e-01 | -0.113 | 0.124 | -0.349 | _ | 0.0161 |
| deCODE_19_44436209_C_T | 6.57e-03 | -0.129 | -0.036 | -0.221 | _ | 0.15 |
| deCODE_19_44442012_G_A | 8.49e-03 | -0.226 | -0.058 | -0.394 | _ | 0.0063 |
| **UKB-PPP_SingleSNP** | | | | | | |
| UKB-PPP_19_45011597_C_T | 8.71e-01 | -0.012 | 0.131 | -0.154 | _ | 0.0209 |
| UKB-PPP_19_44911194_T_C | 8.35e-04 | -0.046 | -0.019 | -0.072 | _ | 0.3603 |
| UKB-PPP_19_44983783_A_G | 4.67e-01 | -0.047 | 0.08 | -0.174 | _ | 0.0126 |
| UKB-PPP_19_44993253_G_A | 4.26e-01 | -0.058 | 0.085 | -0.2 | _ | 0.0097 |
| **combined** | | | | | | |
| deCODE | 2.53e-01 | -0.018 | 0.013 | -0.049 | 9 | _ |
| UKB-PPP | 5.40e-04 | -0.045 | -0.019 | -0.07 | 4 | _ |

#### AA. BCHE_CTP_Cis-pQTL

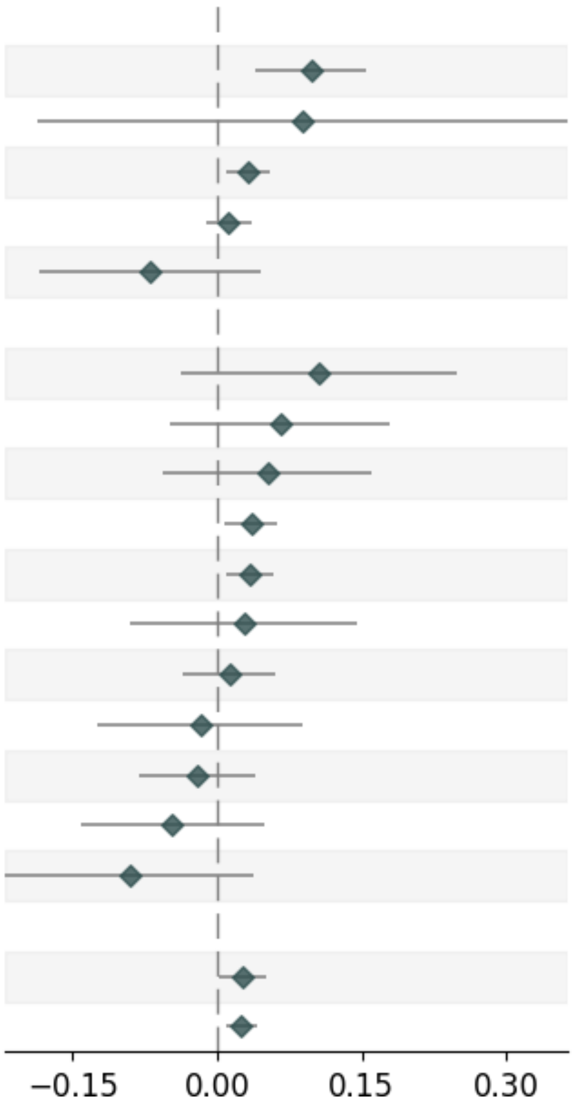


| Variable | Pvalue | Beta | Up_CI | Low_CI | nSNP | MAF |
|---|---|---|---|---|---|---|
| **deCODE_SingleSNP** | | | | | | |
| deCODE_3_165637186_T_C | 8.72e-04 | 0.097 | 0.154 | 0.04 | _ | 0.2971 |
| deCODE_3_165267390_C_T | 5.29e-01 | 0.089 | 0.364 | -0.187 | _ | 0.0413 |
| deCODE_3_165718306_C_T | 4.16e-03 | 0.032 | 0.054 | 0.01 | _ | 0.0344 |
| deCODE_3_165914891_T_C | 3.42e-01 | 0.011 | 0.035 | -0.012 | _ | 0.2049 |
| deCODE_3_166219244_T_C | 2.39e-01 | -0.069 | 0.046 | -0.184 | _ | 0.011 |
| **UKB-PPP_SingleSNP** | | | | | | |
| UKB-PPP_3_165241077_A_G | 1.50e-01 | 0.105 | 0.249 | -0.038 | _ | 0.0052 |
| UKB-PPP_3_165463346_T_C | 2.69e-01 | 0.065 | 0.179 | -0.05 | _ | 0.0081 |
| UKB-PPP_3_166237485_G_A | 3.50e-01 | 0.052 | 0.16 | -0.057 | _ | 0.0079 |
| UKB-PPP_3_165917062_T_C | 1.21e-02 | 0.035 | 0.062 | 0.008 | _ | 0.0304 |
| UKB-PPP_3_165717854_G_A | 5.29e-03 | 0.034 | 0.058 | 0.01 | _ | 0.459 |
| UKB-PPP_3_166054790_T_C | 6.45e-01 | 0.028 | 0.145 | -0.09 | _ | 0.0183 |
| UKB-PPP_3_165767696_C_A | 5.98e-01 | 0.013 | 0.06 | -0.035 | _ | 0.0084 |
| UKB-PPP_3_166679267_A_G | 7.50e-01 | -0.017 | 0.089 | -0.124 | _ | 0.0322 |
| UKB-PPP_3_165480065_T_G | 4.96e-01 | -0.021 | 0.039 | -0.081 | _ | 0.0566 |
| UKB-PPP_3_165317696_C_T | 3.39e-01 | -0.047 | 0.049 | -0.142 | _ | 0.0101 |
| UKB-PPP_3_166552672_C_A | 1.68e-01 | -0.091 | 0.038 | -0.22 | _ | 0.0113 |
| **combined** | | | | | | |
| deCODE | 3.66e-02 | 0.026 | 0.05 | 0.002 | 5 | _ |
| UKB-PPP | 1.79e-03 | 0.025 | 0.041 | 0.009 | 11 | _ |

**AB. CD14_CTP_Cis-pQTL**

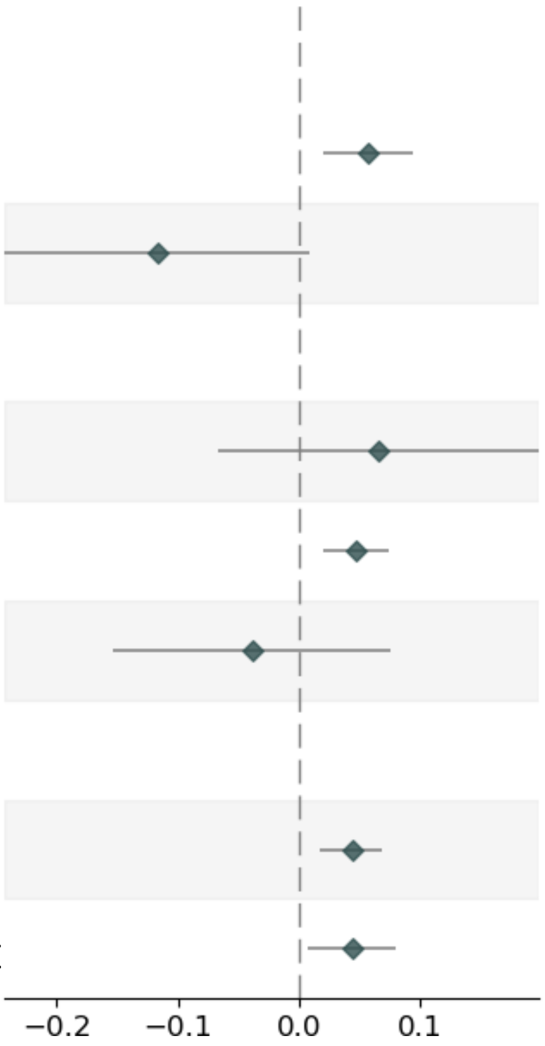


| Variable | Pvalue | Beta | Up_CI | Low_CI | nSNP | MAF |
|---|---|---|---|---|---|---|
| **deCODE_SingleSNP** | | | | | | |
| deCODE_5_140633982_T_G | 2.77e-03 | 0.057 | 0.094 | 0.02 | _ | 0.3074 |
| deCODE_5_141287515_G_A | 6.92e-02 | -0.117 | 0.009 | -0.243 | _ | 0.0375 |
| **UKB-PPP_SingleSNP** | | | | | | |
| UKB-PPP_5_141476782_A_G | 3.31e-01 | 0.066 | 0.199 | -0.067 | _ | 0.0148 |
| UKB-PPP_5_140360869_C_T | 5.40e-04 | 0.047 | 0.074 | 0.02 | _ | 0.2335 |
| UKB-PPP_5_141067739_T_C | 5.12e-01 | -0.038 | 0.076 | -0.153 | _ | 0.0519 |
| **combined** | | | | | | |
| UKB-PPP | 8.88e-04 | 0.044 | 0.069 | 0.018 | 3 | _ |
| deCODE | 1.80e-02 | 0.044 | 0.08 | 0.007 | 2 | _ |

### AC. ERBB3_CTP_Cis-pQTL

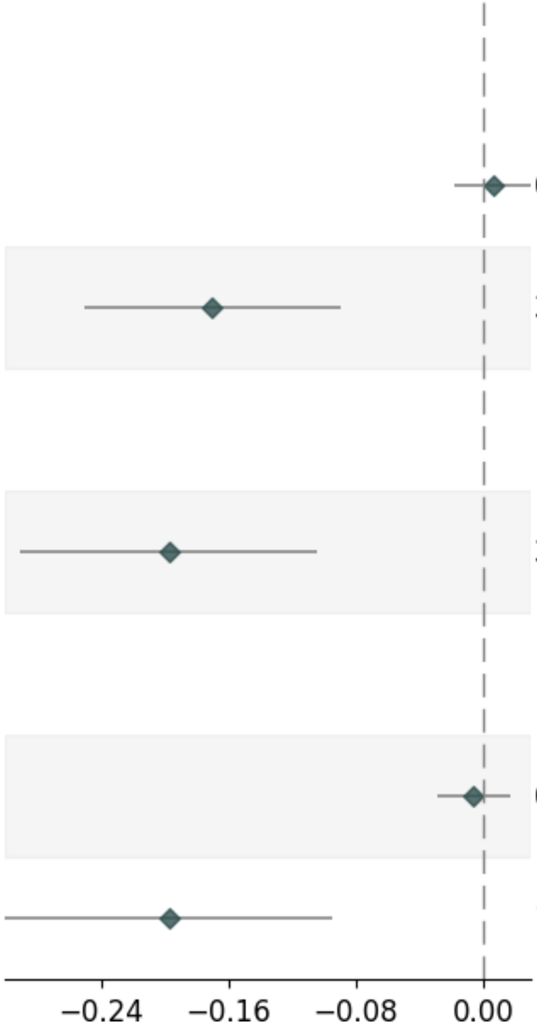


| Variable | Pvalue | Beta | Up_CI | Low_CI | nSNP | MAF |
|---|---|---|---|---|---|---|
| **deCODE_SingleSNP** | | | | | | |
| deCODE_12_56095610_A_G | 6.11e-01 | 0.006 | 0.03 | -0.018 | _ | 0.0011 |
| deCODE_12_55991795_C_T | 3.44e-05 | -0.17 | -0.089 | -0.25 | _ | 0.4517 |
| **UKB-PPP_SingleSNP** | | | | | | |
| UKB-PPP_12_55991795_C_T | 3.44e-05 | -0.197 | -0.104 | -0.291 | _ | 0.4381 |
| **combined** | | | | | | |
| deCODE | 6.15e-01 | -0.006 | 0.017 | -0.029 | 2 | _ |
| UKB-PPP | 1.57e-04 | -0.197 | -0.095 | -0.3 | 1 | _ |

#### AD. HBEGF_CTP_Cis-pQTL

| Variable | Pvalue | Beta | Up_CI | Low_CI | nSNP | MAF |
|---|---|---|---|---|---|---|
| **deCODE_SingleSNP** | | | | | | |
| deCODE_5_140157337_A_G | 4.61e-07 | 0.309 | 0.429 | 0.189 | _ | 0.4388 |
| **UKB-PPP_SingleSNP** | | | | | | |
| UKB-PPP_5_140275366_G_A | 5.93e-08 | 0.399 | 0.544 | 0.255 | _ | 0.444 |
| UKB-PPP_5_140612586_G_A | 6.28e-03 | -0.181 | -0.051 | -0.31 | _ | 0.2898 |
| **combined** | | | | | | |
| deCODE | 1.18e-04 | 0.309 | 0.467 | 0.152 | 1 | _ |
| UKB-PPP | 8.43e-01 | 0.012 | 0.127 | -0.104 | 2 | _ |

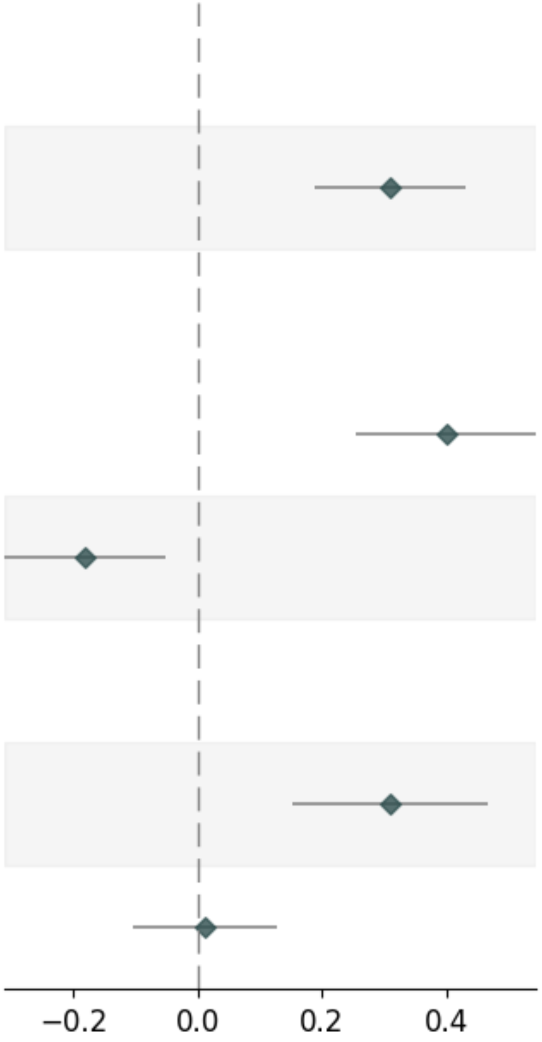

### AE. ICAM5_CTP_Cis-pQTL_DECODE

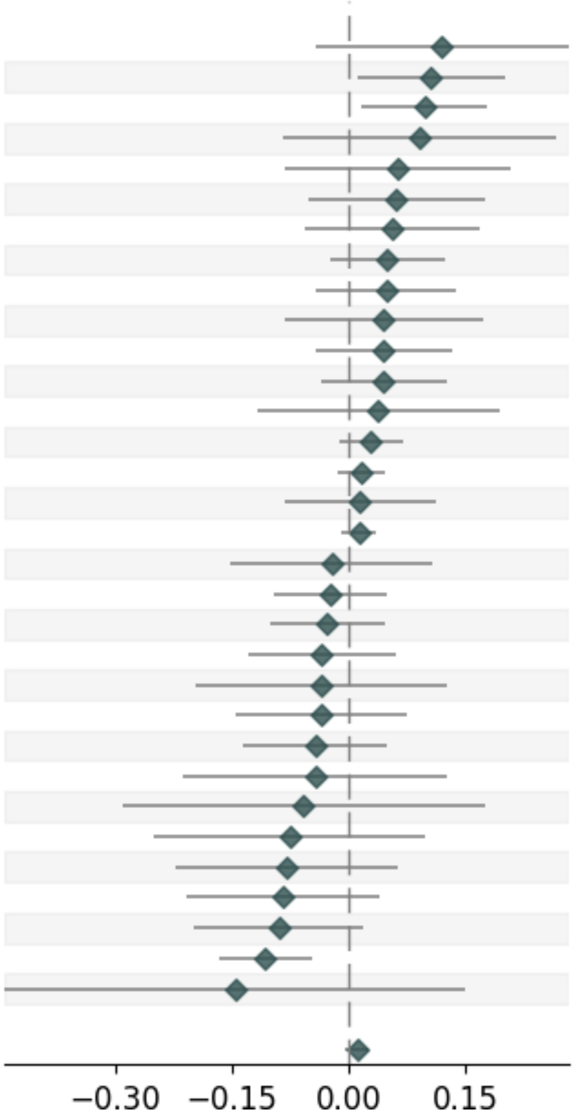


| Variable | Pvalue | Beta | Up_CI | Low_CI | nSNP | MAF |
|---|---|---|---|---|---|---|
| deCODE_SingleSNP | | | | | | |
| deCODE_19_9667048_C_A | 1.49e-01 | 0.12 | 0.283 | -0.043 | _ | 0.0121 |
| deCODE_19_10201842_C_T | 2.79e-02 | 0.106 | 0.201 | 0.011 | _ | 0.034 |
| deCODE_19_10298107_G_A | 1.89e-02 | 0.097 | 0.178 | 0.016 | _ | 0.0059 |
| deCODE_19_10123751_C_A | 3.08e-01 | 0.092 | 0.267 | -0.084 | _ | 0.0135 |
| deCODE_19_11084147_G_A | 3.91e-01 | 0.063 | 0.207 | -0.081 | _ | 0.0628 |
| deCODE_19_9623181_C_T | 2.92e-01 | 0.061 | 0.174 | -0.052 | _ | 0.0103 |
| deCODE_19_10371063_T_G | 3.19e-01 | 0.057 | 0.168 | -0.055 | _ | 0.0269 |
| deCODE_19_10208502_A_G | 1.87e-01 | 0.05 | 0.124 | -0.024 | _ | 0.0073 |
| deCODE_19_11186157_G_A | 3.02e-01 | 0.048 | 0.138 | -0.043 | _ | 0.0047 |
| deCODE_19_10212300_G_A | 4.90e-01 | 0.045 | 0.172 | -0.083 | _ | 0.0187 |
| deCODE_19_10738626_T_C | 3.13e-01 | 0.045 | 0.133 | -0.042 | _ | 0.0203 |
| deCODE_19_9885167_C_T | 2.78e-01 | 0.044 | 0.125 | -0.036 | _ | 0.1538 |
| deCODE_19_9443199_G_A | 6.29e-01 | 0.038 | 0.193 | -0.117 | _ | 0.0103 |
| deCODE_19_10325886_C_T | 1.54e-01 | 0.029 | 0.069 | -0.011 | _ | 0.1305 |
| deCODE_19_10163778_T_C | 3.09e-01 | 0.016 | 0.047 | -0.015 | _ | 0.025 |
| deCODE_19_10249480_C_T | 7.66e-01 | 0.015 | 0.113 | -0.083 | _ | 0.0105 |
| deCODE_19_10272297_G_A | 2.50e-01 | 0.013 | 0.036 | -0.009 | _ | 0.0589 |
| deCODE_19_11186179_A_G | 7.42e-01 | -0.022 | 0.108 | -0.152 | _ | 0.0858 |
| deCODE_19_10255216_C_T | 5.27e-01 | -0.024 | 0.05 | -0.097 | _ | 0.0054 |
| deCODE_19_10724367_T_C | 4.64e-01 | -0.027 | 0.046 | -0.1 | _ | 0.0145 |
| deCODE_19_9971108_G_A | 4.85e-01 | -0.034 | 0.061 | -0.128 | _ | 0.0095 |
| deCODE_19_10174408_T_C | 6.73e-01 | -0.035 | 0.127 | -0.197 | _ | 0.0095 |
| deCODE_19_10622406_C_T | 5.30e-01 | -0.035 | 0.075 | -0.145 | _ | 0.036 |
| deCODE_19_10293588_A_G | 3.59e-01 | -0.043 | 0.049 | -0.136 | _ | 0.0082 |
| deCODE_19_10330152_G_A | 6.20e-01 | -0.043 | 0.127 | -0.213 | _ | 0.021 |
| deCODE_19_9871921_C_T | 6.26e-01 | -0.058 | 0.175 | -0.29 | _ | 0.0139 |
| deCODE_19_10738372_T_G | 3.96e-01 | -0.076 | 0.099 | -0.251 | _ | 0.0161 |
| deCODE_19_9404759_C_A | 2.74e-01 | -0.08 | 0.063 | -0.223 | _ | 0.0054 |
| deCODE_19_10817405_C_T | 1.80e-01 | -0.085 | 0.039 | -0.209 | _ | 0.0052 |
| deCODE_19_10895917_C_T | 1.07e-01 | -0.09 | 0.019 | -0.199 | _ | 0.0077 |
| deCODE_19_11017689_C_T | 4.98e-04 | -0.107 | -0.047 | -0.167 | _ | 0.0098 |
| deCODE_19_10077499_C_T | 3.32e-01 | -0.146 | 0.149 | -0.441 | _ | 0.0046 |
| **combined** | | | | | | |
| deCODE | 1.75e-01 | 0.011 | 0.026 | -0.005 | 32 | _ |

### AF. ICAM5_CTP_Cis-pQTL_UKBB-PPP

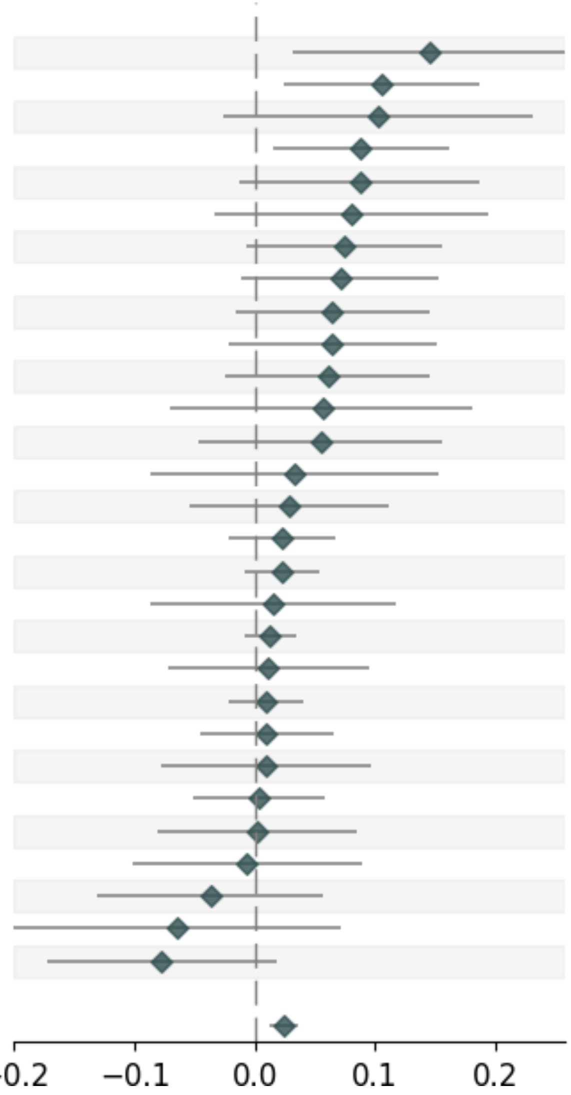

| Variable | Pvalue | Beta | Up_CI | Low_CI | nSNP | MAF |
|---|---|---|---|---|---|---|
| **UKB-PPP_SingleSNP** | | | | | | |
| UKB-PPP_19_10354167_G_A | 1.18e-02 | 0.145 | 0.258 | 0.032 | _ | 0.008 |
| UKB-PPP_19_10168183_G_T | 1.12e-02 | 0.105 | 0.186 | 0.024 | _ | 0.0094 |
| UKB-PPP_19_10467485_C_T | 1.19e-01 | 0.102 | 0.231 | -0.026 | _ | 0.0053 |
| UKB-PPP_19_10298107_G_A | 1.89e-02 | 0.088 | 0.162 | 0.015 | _ | 0.0063 |
| UKB-PPP_19_10265313_C_T | 8.76e-02 | 0.087 | 0.186 | -0.013 | _ | 0.0137 |
| UKB-PPP_19_10459155_C_T | 1.66e-01 | 0.08 | 0.194 | -0.033 | _ | 0.0086 |
| UKB-PPP_19_10347287_G_A | 7.41e-02 | 0.074 | 0.155 | -0.007 | _ | 0.0114 |
| UKB-PPP_19_10195878_G_A | 8.84e-02 | 0.071 | 0.153 | -0.011 | _ | 0.0189 |
| UKB-PPP_19_10300004_C_T | 1.19e-01 | 0.064 | 0.145 | -0.016 | _ | 0.0127 |
| UKB-PPP_19_10608379_G_A | 1.46e-01 | 0.064 | 0.151 | -0.022 | _ | 0.0103 |
| UKB-PPP_19_10160530_G_A | 1.57e-01 | 0.061 | 0.146 | -0.024 | _ | 0.0133 |
| UKB-PPP_19_10114395_A_G | 3.87e-01 | 0.056 | 0.181 | -0.07 | _ | 0.0054 |
| UKB-PPP_19_10060589_C_T | 2.92e-01 | 0.055 | 0.156 | -0.047 | _ | 0.0127 |
| UKB-PPP_19_10299506_C_T | 5.87e-01 | 0.033 | 0.153 | -0.087 | _ | 0.0067 |
| UKB-PPP_19_10212300_G_A | 4.90e-01 | 0.029 | 0.112 | -0.054 | _ | 0.0098 |
| UKB-PPP_19_10163778_T_C | 3.09e-01 | 0.023 | 0.067 | -0.021 | _ | 0.0186 |
| UKB-PPP_19_10325886_C_T | 1.54e-01 | 0.023 | 0.054 | -0.009 | _ | 0.1116 |
| UKB-PPP_19_10249480_C_T | 7.66e-01 | 0.015 | 0.117 | -0.086 | _ | 0.0165 |
| UKB-PPP_19_10272297_G_A | 2.50e-01 | 0.013 | 0.035 | -0.009 | _ | 0.0538 |
| UKB-PPP_19_10103827_G_A | 7.89e-01 | 0.011 | 0.095 | -0.072 | _ | 0.0145 |
| UKB-PPP_19_10171310_C_T | 5.27e-01 | 0.01 | 0.041 | -0.021 | _ | 0.0159 |
| UKB-PPP_19_10273970_C_T | 7.26e-01 | 0.01 | 0.066 | -0.046 | _ | 0.0053 |
| UKB-PPP_19_10309650_T_C | 8.39e-01 | 0.009 | 0.096 | -0.078 | _ | 0.0077 |
| UKB-PPP_19_10250069_A_T | 9.10e-01 | 0.003 | 0.058 | -0.051 | _ | 0.0139 |
| UKB-PPP_19_10287455_A_C | 9.61e-01 | 0.002 | 0.085 | -0.081 | _ | 0.0051 |
| UKB-PPP_19_10322313_T_C | 8.91e-01 | -0.007 | 0.089 | -0.102 | _ | 0.0067 |
| UKB-PPP_19_10107715_C_T | 4.33e-01 | -0.037 | 0.056 | -0.131 | _ | 0.0165 |
| UKB-PPP_19_10293588_A_G | 3.59e-01 | -0.064 | 0.072 | -0.2 | _ | 0.0057 |
| UKB-PPP_19_10456649_C_T | 1.13e-01 | -0.078 | 0.018 | -0.173 | _ | 0.0167 |
| **combined** | | | | | | |
| UKB-PPP | 7.43e-05 | 0.024 | 0.036 | 0.012 | 29 | _ |

### AG. ITIH4_CTP_Cis-pQTL

| Variable | Pvalue | Beta | Up_CI | Low_CI | nSNP | MAF |
|---|---|---|---|---|---|---|
| **UKB-PPP_SingleSNP** | | | | | | |
| UKB-PPP_3_52785311_T_A | 9.98e-07 | 0.081 | 0.113 | 0.049 | _ | 0.3953 |
| UKB-PPP_3_53007170_T_C | 7.51e-01 | 0.025 | 0.179 | -0.129 | _ | 0.0184 |
| UKB-PPP_3_53029815_G_A | 8.01e-02 | -0.071 | 0.008 | -0.15 | _ | 0.0106 |
| **combined** | | | | | | |
| UKB-PPP | 1.42e-04 | 0.058 | 0.088 | 0.028 | 3 | _ |

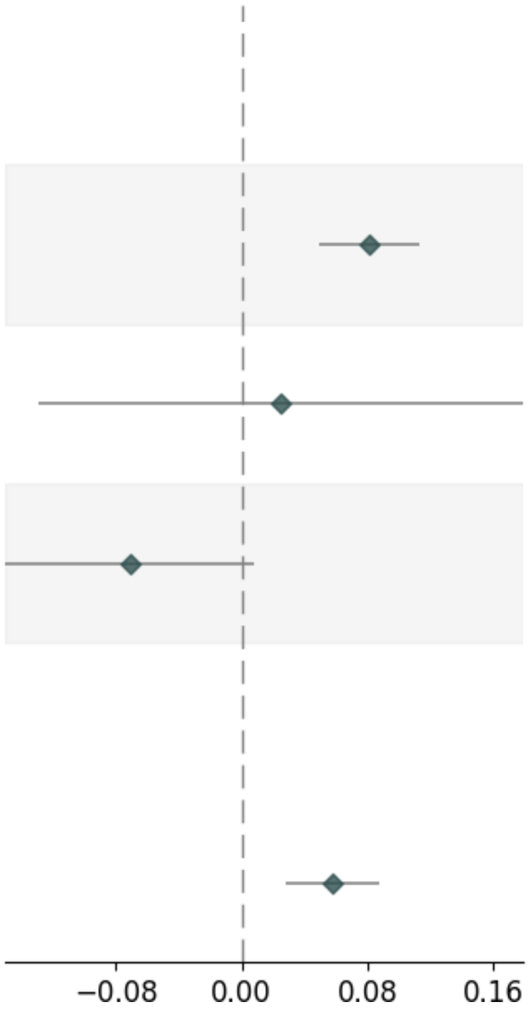

#### AH. NAGK_CTP_Cis-pQTL

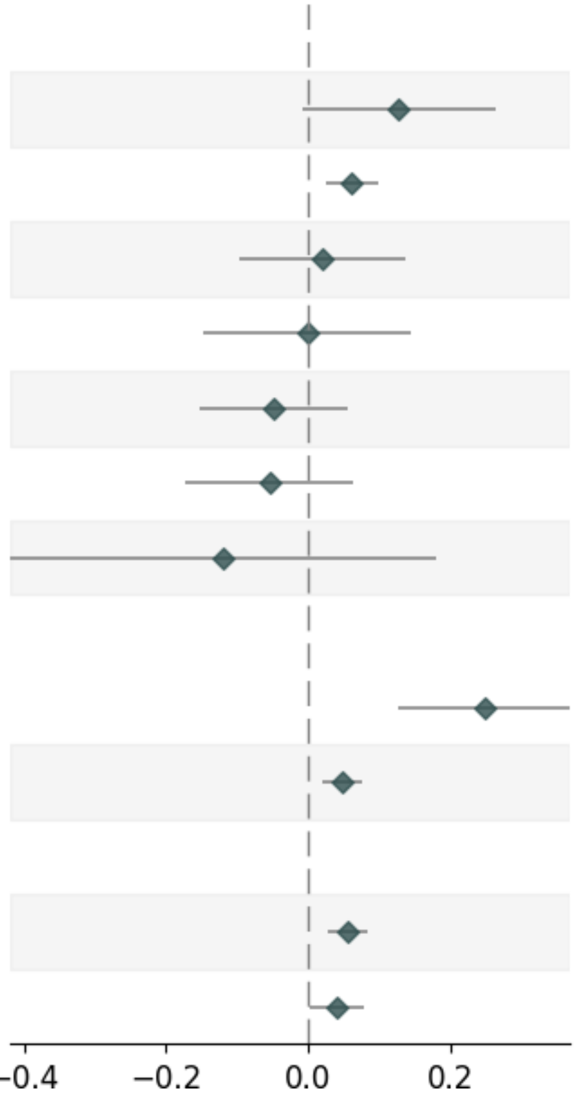


| Variable | Pvalue | Beta | Up_CI | Low_CI | nSNP | MAF |
|---|---|---|---|---|---|---|
| **deCODE_SingleSNP** | | | | | | |
| deCODE_2_71114512_A_C | 6.65e-02 | 0.127 | 0.263 | -0.009 | _ | 0.1754 |
| deCODE_2_71065392_T_C | 1.02e-03 | 0.062 | 0.099 | 0.025 | _ | 0.3755 |
| deCODE_2_72037670_G_A | 7.36e-01 | 0.02 | 0.136 | -0.096 | _ | 0.0052 |
| deCODE_2_71916310_T_C | 9.84e-01 | -0.001 | 0.144 | -0.147 | _ | 0.0698 |
| deCODE_2_70828835_T_G | 3.55e-01 | -0.049 | 0.055 | -0.152 | _ | 0.0264 |
| deCODE_2_71507405_T_C | 3.70e-01 | -0.054 | 0.064 | -0.172 | _ | 0.038 |
| deCODE_2_70751193_C_T | 4.35e-01 | -0.119 | 0.18 | -0.419 | _ | 0.0306 |
| **UKB-PPP_SingleSNP** | | | | | | |
| UKB-PPP_2_71404938_T_C | 5.97e-05 | 0.248 | 0.369 | 0.127 | _ | 0.2986 |
| UKB-PPP_2_71065392_T_C | 1.02e-03 | 0.047 | 0.076 | 0.019 | _ | 0.3423 |
| **combined** | | | | | | |
| UKB-PPP | 1.05e-04 | 0.055 | 0.083 | 0.027 | 2 | _ |
| deCODE | 4.17e-02 | 0.04 | 0.078 | 0.002 | 7 | _ |

### AI. NEGR1_CTP_Cis-pQTL

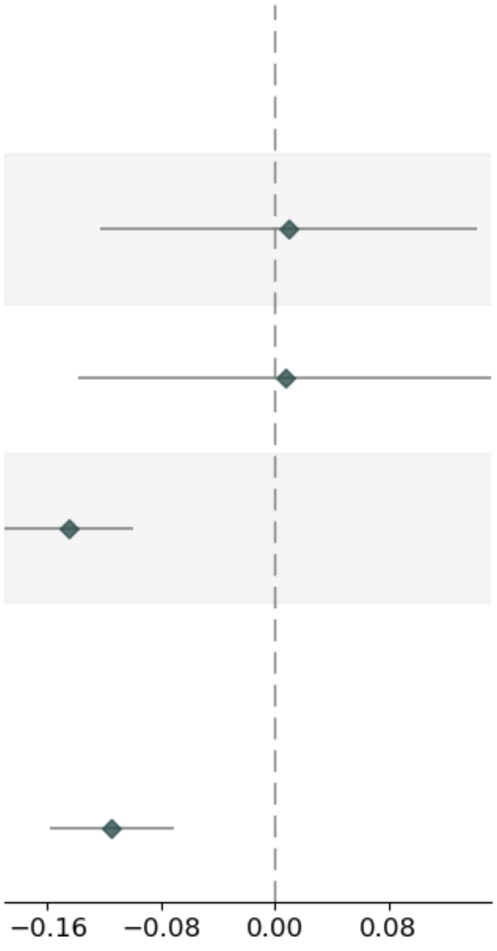


| Variable | Pvalue | Beta | Up_CI | Low_CI | nSNP | MAF |
|---|---|---|---|---|---|---|
| **deCODE_SingleSNP** | | | | | | |
| deCODE_1_71980481_A_G | 8.89e-01 | 0.009 | 0.142 | -0.123 | _ | 0.2649 |
| deCODE_1_70578253_C_T | 9.28e-01 | 0.007 | 0.152 | -0.138 | _ | 0.007 |
| deCODE_1_72243359_G_A | 3.32e-10 | -0.145 | -0.1 | -0.19 | _ | 0.4604 |
| **combined** | | | | | | |
| deCODE | 2.53e-07 | -0.115 | -0.071 | -0.158 | 3 | _ |

### AJ. PLA2G15_CTP_Cis-pQTL

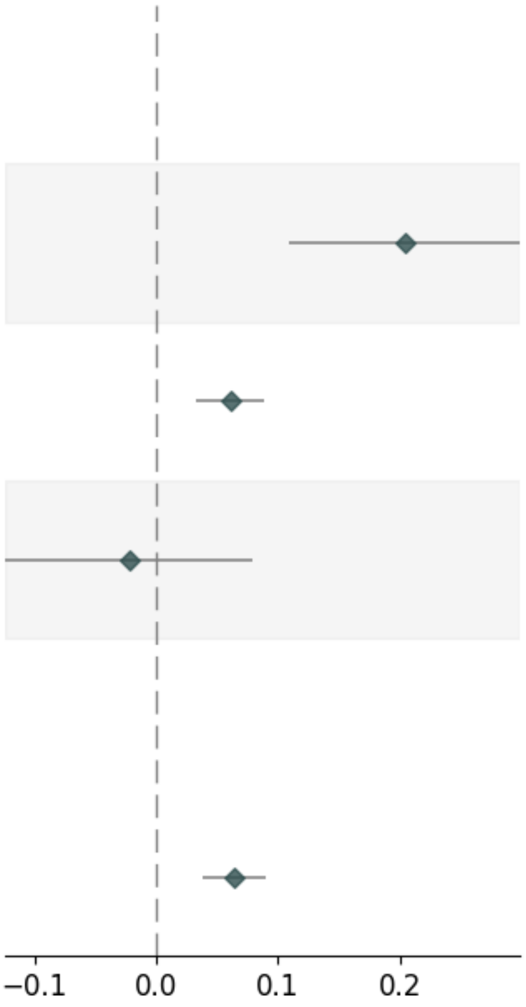


| Variable | Pvalue | Beta | Up_CI | Low_CI | nSNP | MAF |
|---|---|---|---|---|---|---|
| **UKB-PPP_SingleSNP** | | | | | | |
| UKB-PPP_16_68488494_C_A | 2.59e-05 | 0.204 | 0.299 | 0.109 | _ | 0.2222 |
| UKB-PPP_16_68128113_G_A | 1.65e-05 | 0.061 | 0.088 | 0.033 | _ | 0.1153 |
| UKB-PPP_16_67892482_A_G | 6.67e-01 | -0.022 | 0.079 | -0.124 | _ | 0.0062 |
| **combined** | | | | | | |
| UKB-PPP | 1.60e-06 | 0.064 | 0.09 | 0.038 | 3 | _ |

### AK. SCG3_CTP_Cis-pQTL

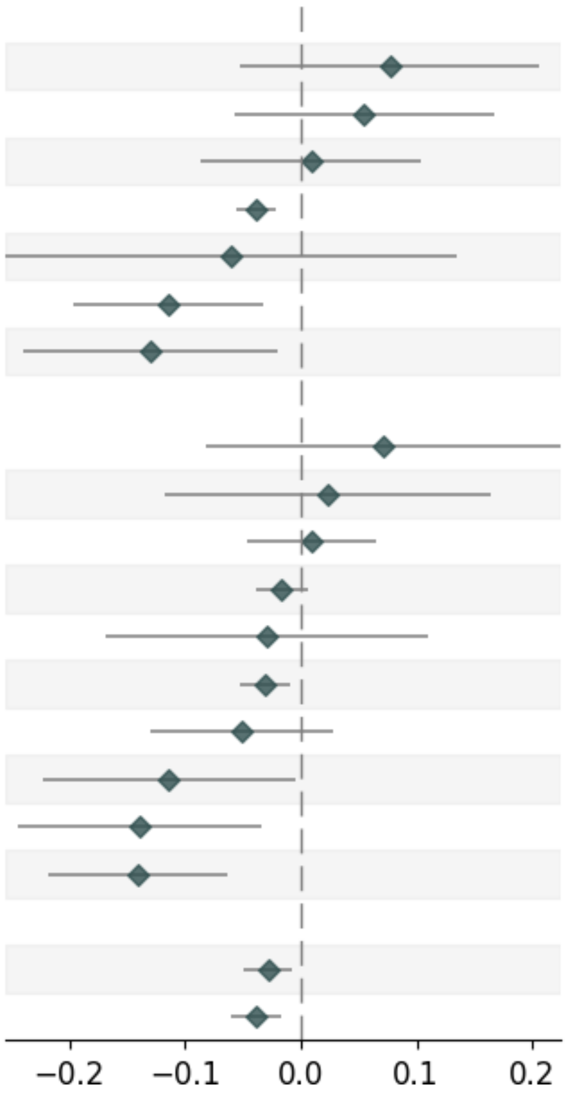


| Variable | Pvalue | Beta | Up_CI | Low_CI | nSNP | MAF |
|---|---|---|---|---|---|---|
| **deCODE_SingleSNP** | | | | | | |
| deCODE_15_52127441_A_G | 2.39e-01 | 0.077 | 0.206 | -0.052 | _ | 0.0041 |
| deCODE_15_50873983_T_C | 3.45e-01 | 0.054 | 0.167 | -0.058 | _ | 0.0074 |
| deCODE_15_51022847_C_T | 8.50e-01 | 0.009 | 0.104 | -0.086 | _ | 0.0251 |
| deCODE_15_51403538_A_T | 2.97e-06 | -0.039 | -0.022 | -0.055 | _ | 0.1662 |
| deCODE_15_52425548_C_T | 5.42e-01 | -0.06 | 0.134 | -0.255 | _ | 0.0078 |
| deCODE_15_52416482_G_A | 6.14e-03 | -0.114 | -0.033 | -0.196 | _ | 0.1332 |
| deCODE_15_52022492_G_A | 2.04e-02 | -0.13 | -0.02 | -0.24 | _ | 0.0066 |
| **UKB-PPP_SingleSNP** | | | | | | |
| UKB-PPP_15_51291789_T_G | 3.62e-01 | 0.071 | 0.225 | -0.082 | _ | 0.0148 |
| UKB-PPP_15_52375492_T_C | 7.50e-01 | 0.023 | 0.164 | -0.118 | _ | 0.0065 |
| UKB-PPP_15_51427651_A_T | 7.48e-01 | 0.009 | 0.065 | -0.047 | _ | 0.0566 |
| UKB-PPP_15_51391668_C_T | 1.43e-01 | -0.017 | 0.006 | -0.039 | _ | 0.3004 |
| UKB-PPP_15_52580124_G_A | 6.77e-01 | -0.03 | 0.11 | -0.169 | _ | 0.0923 |
| UKB-PPP_15_51720904_G_A | 6.20e-03 | -0.031 | -0.009 | -0.053 | _ | 0.3922 |
| UKB-PPP_15_50891514_G_A | 2.08e-01 | -0.051 | 0.028 | -0.13 | _ | 0.0103 |
| UKB-PPP_15_50691346_C_T | 4.19e-02 | -0.114 | -0.004 | -0.223 | _ | 0.2122 |
| UKB-PPP_15_51183612_T_C | 9.68e-03 | -0.139 | -0.034 | -0.244 | _ | 0.0071 |
| UKB-PPP_15_50686870_C_A | 3.28e-04 | -0.141 | -0.064 | -0.218 | _ | 0.013 |
| **combined** | | | | | | |
| UKB-PPP | 6.67e-03 | -0.028 | -0.008 | -0.049 | 10 | _ |
| deCODE | 5.26e-04 | -0.038 | -0.017 | -0.06 | 7 | _ |

#### AL. SEMA3F_CTP_Cis-pQTL

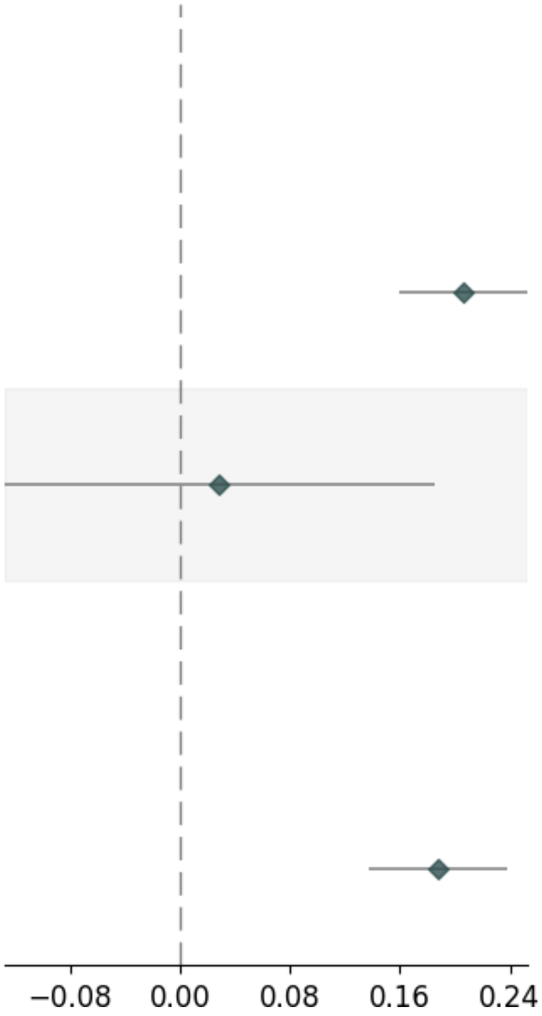


| Variable | Pvalue | Beta | Up_CI | Low_CI | nSNP | MAF |
|---|---|---|---|---|---|---|
| **UKB-PPP_SingleSNP** | | | | | | |
| UKB-PPP_3_50137139_C_T | 9.00e-18 | 0.206 | 0.253 | 0.159 | _ | 0.3556 |
| UKB-PPP_3_50087849_T_C | 7.20e-01 | 0.029 | 0.185 | -0.127 | _ | 0.0257 |
| **combined** | | | | | | |
| UKB-PPP | 1.24e-13 | 0.188 | 0.238 | 0.138 | 2 | _ |

### AM. SIRPA_CTP_Cis-pQTL_UKB-PPP

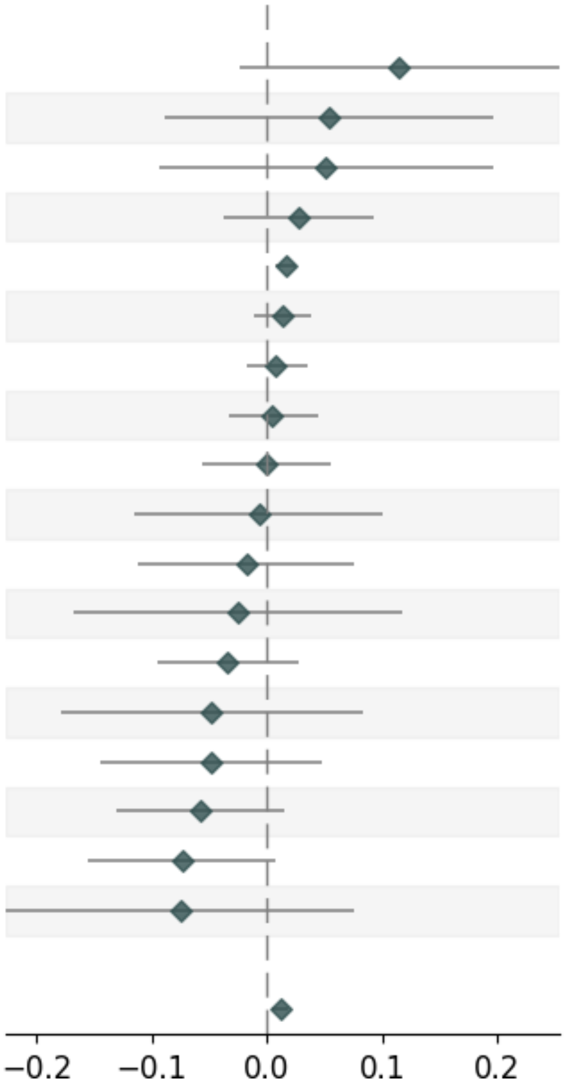


| Variable | Pvalue | Beta | Up_CI | Low_CI | nSNP | MAF |
|---|---|---|---|---|---|---|
| **UKB-PPP_SingleSNP** | | | | | | |
| UKB-PPP_20_2179590_C_T | 1.06e-01 | 0.115 | 0.254 | -0.024 | _ | 0.0157 |
| UKB-PPP_20_2092409_C_T | 4.62e-01 | 0.054 | 0.197 | -0.089 | _ | 0.0075 |
| UKB-PPP_20_2165113_C_T | 4.85e-01 | 0.051 | 0.196 | -0.093 | _ | 0.0086 |
| UKB-PPP_20_1930044_G_A | 4.11e-01 | 0.028 | 0.093 | -0.038 | _ | 0.0478 |
| UKB-PPP_20_1902308_T_C | 6.22e-04 | 0.016 | 0.025 | 0.007 | _ | 0.1767 |
| UKB-PPP_20_1888701_T_C | 2.59e-01 | 0.014 | 0.039 | -0.011 | _ | 0.2753 |
| UKB-PPP_20_1815815_G_T | 5.45e-01 | 0.008 | 0.035 | -0.018 | _ | 0.4105 |
| UKB-PPP_20_1570839_C_T | 7.88e-01 | 0.005 | 0.044 | -0.033 | _ | 0.3543 |
| UKB-PPP_20_2050237_C_T | 9.78e-01 | -0.001 | 0.055 | -0.056 | _ | 0.0431 |
| UKB-PPP_20_1986052_T_C | 9.04e-01 | -0.007 | 0.101 | -0.115 | _ | 0.0137 |
| UKB-PPP_20_1981084_C_T | 7.11e-01 | -0.018 | 0.076 | -0.111 | _ | 0.0123 |
| UKB-PPP_20_1701698_G_A | 7.29e-01 | -0.025 | 0.117 | -0.167 | _ | 0.0106 |
| UKB-PPP_20_1803759_G_A | 2.83e-01 | -0.034 | 0.028 | -0.095 | _ | 0.2437 |
| UKB-PPP_20_1194130_G_T | 4.75e-01 | -0.048 | 0.083 | -0.178 | _ | 0.1827 |
| UKB-PPP_20_1998527_A_C | 3.16e-01 | -0.049 | 0.047 | -0.144 | _ | 0.0185 |
| UKB-PPP_20_1908497_G_A | 1.20e-01 | -0.058 | 0.015 | -0.131 | _ | 0.0052 |
| UKB-PPP_20_2026077_G_A | 7.69e-02 | -0.073 | 0.008 | -0.155 | _ | 0.012 |
| UKB-PPP_20_1941574_C_T | 3.28e-01 | -0.075 | 0.075 | -0.226 | _ | 0.0069 |
| **combined** | | | | | | |
| UKB-PPP | 3.76e-03 | 0.012 | 0.019 | 0.004 | 18 | _ |

#### AN. SIRPA_CTP_Cis-pQTL_DECODE_Part1

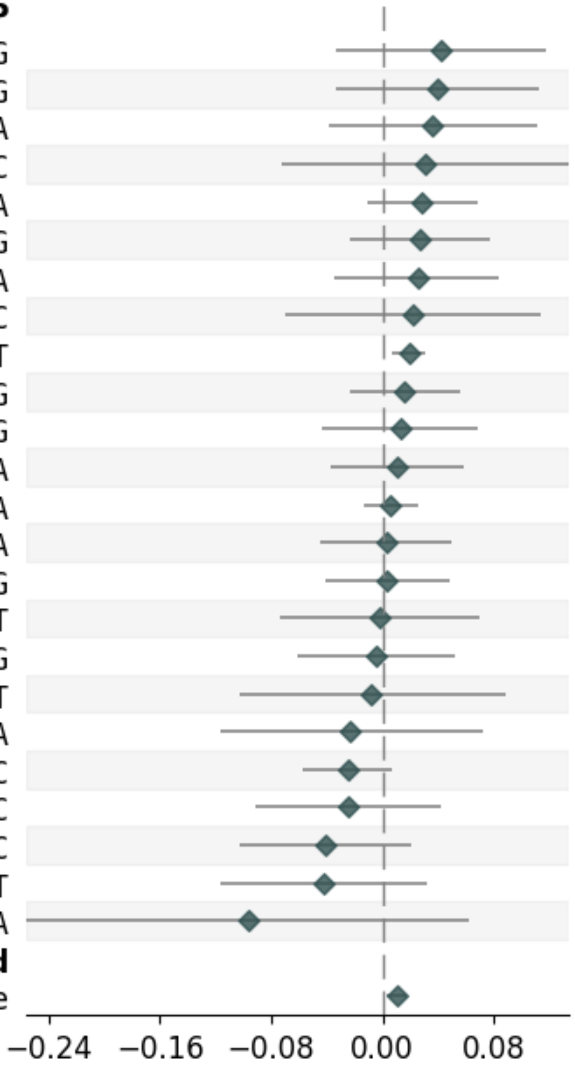


| Variable | Pvalue | Beta | Up_CI | Low_CI | nSNP | MAF |
|---|---|---|---|---|---|---|
| **Decode_SingleSNP** | | | | | | |
| Decode_20_1157969_A_G | 2.74e-01 | 0.042 | 0.117 | -0.033 | _ | 0.0258 |
| Decode_20_2114071_A_G | 2.91e-01 | 0.04 | 0.113 | -0.034 | _ | 0.019 |
| Decode_20_1927548_G_A | 3.47e-01 | 0.036 | 0.111 | -0.039 | _ | 0.0072 |
| Decode_20_1191805_T_C | 5.63e-01 | 0.031 | 0.134 | -0.073 | _ | 0.0229 |
| Decode_20_1871454_C_A | 1.57e-01 | 0.028 | 0.068 | -0.011 | _ | 0.0774 |
| Decode_20_1190465_A_G | 2.82e-01 | 0.027 | 0.077 | -0.023 | _ | 0.0919 |
| Decode_20_1930044_G_A | 4.11e-01 | 0.025 | 0.084 | -0.035 | _ | 0.044 |
| Decode_20_2072291_T_C | 6.39e-01 | 0.022 | 0.114 | -0.07 | _ | 0.0321 |
| Decode_20_1899904_C_T | 1.65e-03 | 0.019 | 0.031 | 0.007 | _ | 0.1041 |
| Decode_20_1934491_A_G | 4.40e-01 | 0.016 | 0.056 | -0.024 | _ | 0.0076 |
| Decode_20_1908019_A_G | 6.48e-01 | 0.013 | 0.069 | -0.043 | _ | 0.0126 |
| Decode_20_1910795_G_A | 6.63e-01 | 0.011 | 0.058 | -0.037 | _ | 0.0145 |
| Decode_20_1936563_G_A | 5.70e-01 | 0.006 | 0.026 | -0.014 | _ | 0.0679 |
| Decode_20_1898505_G_A | 9.15e-01 | 0.003 | 0.05 | -0.045 | _ | 0.0108 |
| Decode_20_2009783_T_G | 8.84e-01 | 0.003 | 0.048 | -0.041 | _ | 0.0284 |
| Decode_20_2052585_C_T | 9.50e-01 | -0.002 | 0.07 | -0.074 | _ | 0.0123 |
| Decode_20_1142143_A_G | 8.84e-01 | -0.004 | 0.052 | -0.061 | _ | 0.053 |
| Decode_20_1017639_C_T | 8.74e-01 | -0.008 | 0.088 | -0.103 | _ | 0.0112 |
| Decode_20_1031106_G_A | 6.39e-01 | -0.023 | 0.072 | -0.117 | _ | 0.0199 |
| Decode_20_1795547_T_C | 1.23e-01 | -0.025 | 0.007 | -0.058 | _ | 0.1765 |
| Decode_20_2009971_T_C | 4.62e-01 | -0.025 | 0.042 | -0.092 | _ | 0.0198 |
| Decode_20_1405219_T_C | 1.89e-01 | -0.041 | 0.02 | -0.103 | _ | 0.273 |
| Decode_20_1631194_C_T | 2.66e-01 | -0.042 | 0.032 | -0.117 | _ | 0.0042 |
| Decode_20_2271504_T_A | 2.30e-01 | -0.097 | 0.062 | -0.256 | _ | 0.026 |
| **combined** | | | | | | |
| Decode | 6.57e-03 | 0.01 | 0.017 | 0.003 | 48 | _ |

### AO. SIRPA_CTP_Cis-pQTL_DECODE_2

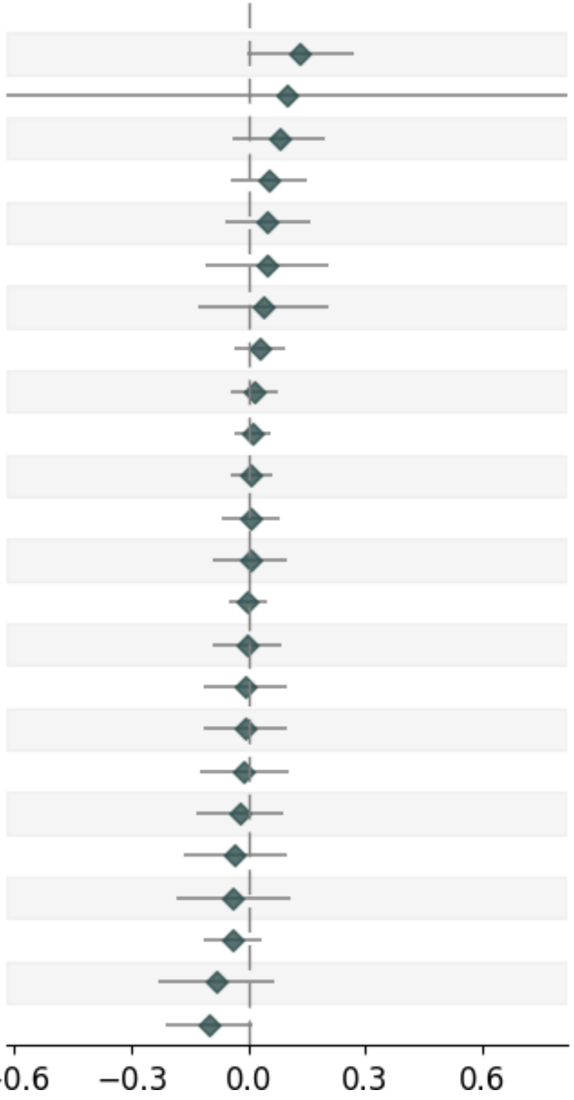


| Variable | Pvalue | Beta | Up_CI | Low_CI | nSNP | MAF |
|---|---|---|---|---|---|---|
| **deCODE_SingleSNP** | | | | | | |
| deCODE_20_2530335_G_A | 5.92e-02 | 0.132 | 0.268 | -0.005 | _ | 0.0106 |
| deCODE_20_2689159_A_G | 7.87e-01 | 0.099 | 0.818 | -0.62 | _ | 0.0045 |
| deCODE_20_981440_G_A | 2.00e-01 | 0.078 | 0.197 | -0.041 | _ | 0.0114 |
| deCODE_20_2518347_T_C | 2.85e-01 | 0.053 | 0.151 | -0.044 | _ | 0.0071 |
| deCODE_20_944970_C_A | 3.86e-01 | 0.049 | 0.159 | -0.061 | _ | 0.0061 |
| deCODE_20_998825_T_C | 5.73e-01 | 0.045 | 0.203 | -0.112 | _ | 0.0068 |
| deCODE_20_2744225_C_T | 6.74e-01 | 0.036 | 0.203 | -0.131 | _ | 0.0099 |
| deCODE_20_904459_C_T | 3.70e-01 | 0.029 | 0.092 | -0.034 | _ | 0.0107 |
| deCODE_20_971192_C_T | 6.42e-01 | 0.014 | 0.073 | -0.045 | _ | 0.0176 |
| deCODE_20_2307823_C_T | 6.18e-01 | 0.012 | 0.058 | -0.034 | _ | 0.1343 |
| deCODE_20_2591187_A_G | 8.05e-01 | 0.007 | 0.059 | -0.046 | _ | 0.0332 |
| deCODE_20_2771148_G_T | 9.02e-01 | 0.005 | 0.079 | -0.07 | _ | 0.0603 |
| deCODE_20_900450_G_A | 9.16e-01 | 0.005 | 0.1 | -0.09 | _ | 0.0862 |
| deCODE_20_2335283_C_T | 9.51e-01 | -0.002 | 0.046 | -0.049 | _ | 0.4422 |
| deCODE_20_2727181_C_T | 9.59e-01 | -0.002 | 0.085 | -0.09 | _ | 0.0068 |
| deCODE_20_894452_C_T | 8.74e-01 | -0.008 | 0.097 | -0.114 | _ | 0.0275 |
| deCODE_20_920028_G_A | 8.69e-01 | -0.009 | 0.096 | -0.114 | _ | 0.0267 |
| deCODE_20_2557821_C_A | 8.51e-01 | -0.011 | 0.103 | -0.124 | _ | 0.0111 |
| deCODE_20_976467_A_C | 7.02e-01 | -0.022 | 0.089 | -0.132 | _ | 0.0268 |
| deCODE_20_2537520_G_A | 6.06e-01 | -0.035 | 0.097 | -0.167 | _ | 0.0104 |
| deCODE_20_2630355_G_A | 5.90e-01 | -0.04 | 0.107 | -0.187 | _ | 0.0111 |
| deCODE_20_2656919_G_A | 2.93e-01 | -0.04 | 0.034 | -0.114 | _ | 0.0104 |
| deCODE_20_2669967_G_A | 2.71e-01 | -0.083 | 0.065 | -0.23 | _ | 0.0078 |
| deCODE_20_2737175_G_A | 7.71e-02 | -0.101 | 0.011 | -0.213 | _ | 0.0079 |

#### AP. SPINT1_CTP_Cis-pQTL

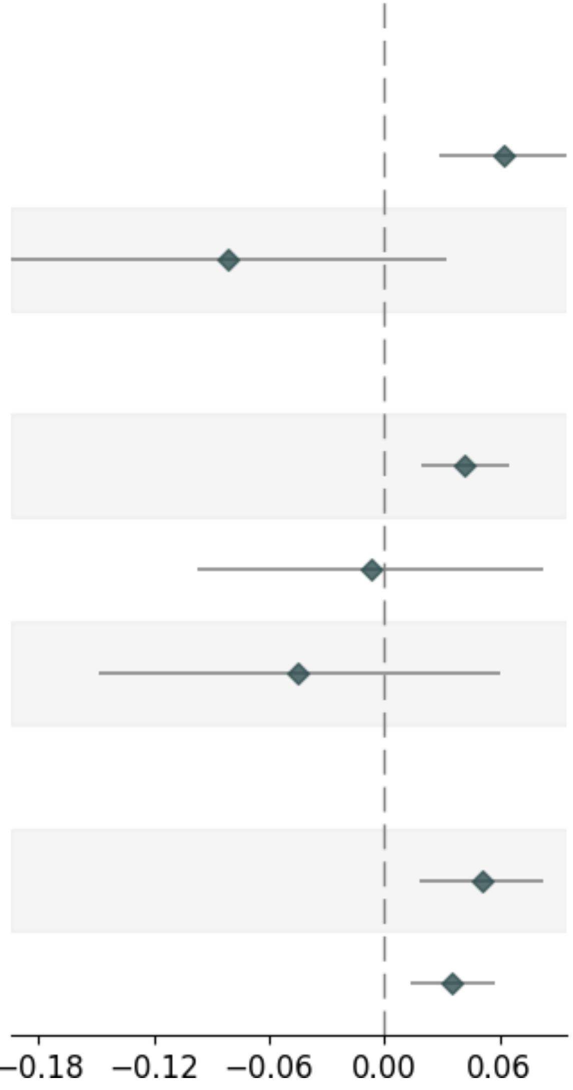


| Variable | Pvalue | Beta | Up_CI | Low_CI | nSNP | MAF |
|---|---|---|---|---|---|---|
| **deCODE_SingleSNP** | | | | | | |
| deCODE_15_40853721_C_T | 2.51e-04 | 0.062 | 0.095 | 0.029 | _ | 0.0475 |
| deCODE_15_40297623_G_A | 1.62e-01 | -0.081 | 0.032 | -0.194 | _ | 0.0403 |
| **UKB-PPP_SingleSNP** | | | | | | |
| UKB-PPP_15_40755579_T_C | 3.26e-04 | 0.042 | 0.065 | 0.019 | _ | 0.0631 |
| UKB-PPP_15_40935116_C_T | 8.75e-01 | -0.007 | 0.083 | -0.097 | _ | 0.0194 |
| UKB-PPP_15_40836877_C_T | 4.01e-01 | -0.045 | 0.06 | -0.149 | _ | 0.0078 |
| **combined** | | | | | | |
| deCODE | 2.10e-03 | 0.051 | 0.083 | 0.018 | 2 | _ |
| UKB-PPP | 1.47e-03 | 0.035 | 0.057 | 0.014 | 3 | _ |

### AQ. SULT1A1_CTP_Cis-pQTL

| Variable | Pvalue | Beta | Up_CI | Low_CI | nSNP | MAF |
|---|---|---|---|---|---|---|
| **UKB-PPP_SingleSNP** | | | | | | |
| UKB-PPP_16_28605807_T_C | 4.62e-13 | 0.244 | 0.31 | 0.178 | _ | 0.3545 |
| UKB-PPP_16_29425849_T_G | 1.06e-01 | 0.077 | 0.17 | -0.016 | _ | 0.0773 |
| **combined** | | | | | | |
| UKB-PPP | 5.90e-09 | 0.178 | 0.237 | 0.118 | 2 | _ |

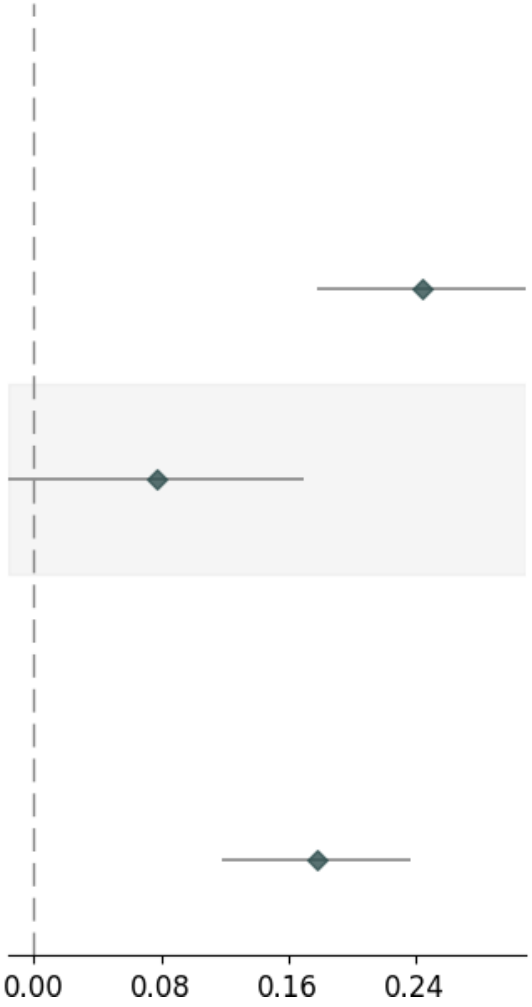

### AR. TIMP4_CTP_Cis-pQTL

| Variable | Pvalue | Beta | Up_CI | Low_CI | nSNP | MAF |
|---|---|---|---|---|---|---|
| **deCODE_SingleSNP** | | | | | | |
| deCODE_3_11939998_T_G | 4.84e-02 | 0.07 | 0.14 | 0.0 | _ | 0.1331 |
| deCODE_3_13079294_T_A | 4.68e-01 | 0.039 | 0.146 | -0.067 | _ | 0.2193 |
| deCODE_3_12005775_A_G | 5.69e-03 | 0.033 | 0.056 | 0.01 | _ | 0.262 |
| deCODE_3_12820758_G_A | 5.60e-01 | 0.027 | 0.116 | -0.063 | _ | 0.3594 |
| deCODE_3_11750830_T_C | 9.74e-01 | -0.001 | 0.085 | -0.088 | _ | 0.0827 |
| deCODE_3_11574093_C_T | 2.39e-01 | -0.117 | 0.078 | -0.312 | _ | 0.022 |
| **UKB-PPP_SingleSNP** | | | | | | |
| UKB-PPP_3_11985235_G_T | 2.86e-05 | 0.059 | 0.087 | 0.032 | _ | 0.1132 |
| UKB-PPP_3_11876274_A_G | 9.50e-01 | -0.004 | 0.134 | -0.143 | _ | 0.335 |
| UKB-PPP_3_12292833_T_C | 2.51e-01 | -0.024 | 0.017 | -0.064 | _ | 0.2708 |
| **combined** | | | | | | |
| deCODE | 1.90e-03 | 0.033 | 0.053 | 0.012 | 6 | _ |
| UKB-PPP | 6.96e-03 | 0.031 | 0.054 | 0.009 | 3 | _ |

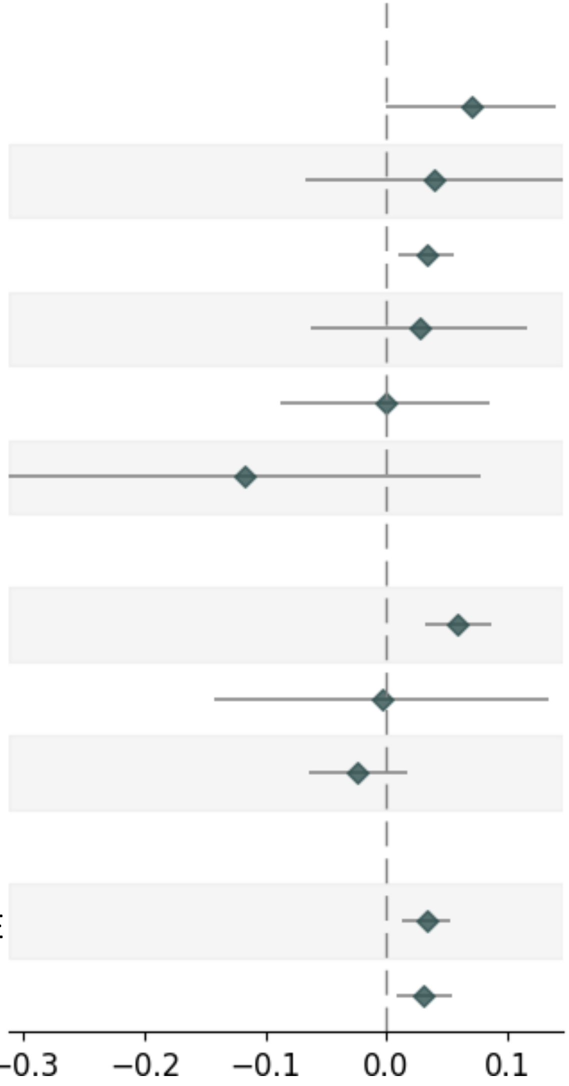

### AS. C15ORF48_SCZ_Trans-pQTL

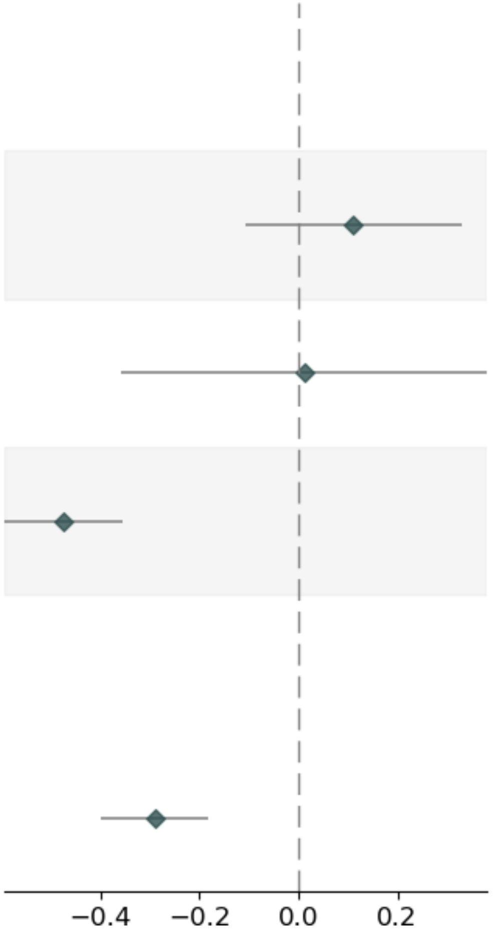


| Variable | Pvalue | Beta | Up_CI | Low_CI | nSNP | MAF |
|---|---|---|---|---|---|---|
| **deCODE_SingleSNP** | | | | | | |
| deCODE_2_31254972_G_A | 3.23e-01 | 0.11 | 0.329 | -0.108 | _ | 0.2847 |
| deCODE_2_226426699_A_C | 9.49e-01 | 0.012 | 0.381 | -0.357 | _ | 0.2142 |
| deCODE_3_52786995_A_G | 7.51e-15 | -0.474 | -0.354 | -0.593 | _ | 0.3891 |
| **combined** | | | | | | |
| deCODE | 1.74e-07 | -0.289 | -0.181 | -0.398 | 3 | _ |

### AT. IL1RL2_SCZ_Trans-pQTL_UKB-PPP

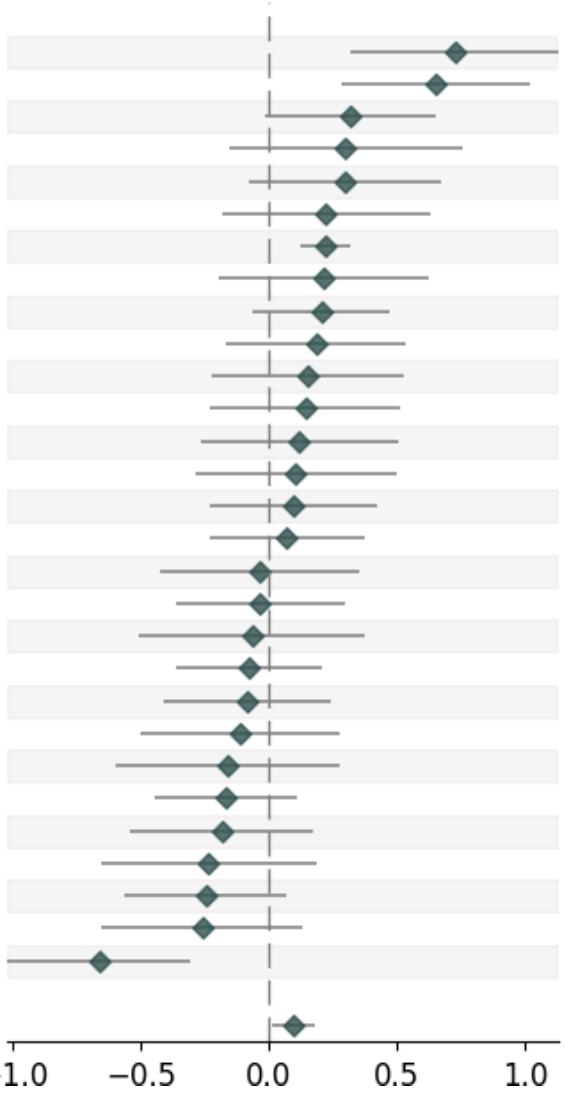


| Variable | Pvalue | Beta | Up_CI | Low_CI | nSNP | MAF |
|---|---|---|---|---|---|---|
| **UKB-PPP_SingleSNP** | | | | | | |
| UKB-PPP_10_63431885_G_T | 4.89e-04 | 0.726 | 1.134 | 0.318 | _ | 0.4987 |
| UKB-PPP_2_103271655_T_C | 5.54e-04 | 0.649 | 1.017 | 0.281 | _ | 0.0523 |
| UKB-PPP_9_5630344_C_T | 5.96e-02 | 0.318 | 0.649 | -0.013 | _ | 0.2806 |
| UKB-PPP_10_80524756_A_T | 1.96e-01 | 0.301 | 0.758 | -0.155 | _ | 0.2079 |
| UKB-PPP_7_73612048_C_T | 1.17e-01 | 0.3 | 0.676 | -0.075 | _ | 0.16 |
| UKB-PPP_6_119467368_T_A | 2.78e-01 | 0.224 | 0.629 | -0.181 | _ | 0.2953 |
| UKB-PPP_12_120985583_A_G | 1.27e-05 | 0.222 | 0.321 | 0.122 | _ | 0.3071 |
| UKB-PPP_16_30968809_A_T | 3.01e-01 | 0.216 | 0.627 | -0.194 | _ | 0.3751 |
| UKB-PPP_6_95559822_C_T | 1.30e-01 | 0.206 | 0.472 | -0.061 | _ | 0.0838 |
| UKB-PPP_16_88039588_C_T | 2.99e-01 | 0.186 | 0.536 | -0.165 | _ | 0.3586 |
| UKB-PPP_3_33416001_C_A | 4.22e-01 | 0.153 | 0.526 | -0.22 | _ | 0.116 |
| UKB-PPP_11_75743976_C_A | 4.56e-01 | 0.142 | 0.515 | -0.231 | _ | 0.1539 |
| UKB-PPP_7_6376984_T_C | 5.36e-01 | 0.121 | 0.505 | -0.263 | _ | 0.2524 |
| UKB-PPP_10_93079885_G_A | 6.00e-01 | 0.105 | 0.498 | -0.288 | _ | 0.4506 |
| UKB-PPP_19_5827754_G_A | 5.72e-01 | 0.094 | 0.421 | -0.233 | _ | 0.0416 |
| UKB-PPP_14_94378610_C_T | 6.46e-01 | 0.071 | 0.373 | -0.231 | _ | 0.0206 |
| UKB-PPP_7_129099519_T_C | 8.68e-01 | -0.033 | 0.357 | -0.423 | _ | 0.4273 |
| UKB-PPP_15_63499029_A_G | 8.41e-01 | -0.034 | 0.295 | -0.362 | _ | 0.3419 |
| UKB-PPP_4_98998475_G_A | 7.71e-01 | -0.066 | 0.378 | -0.51 | _ | 0.2784 |
| UKB-PPP_19_35547488_A_G | 6.00e-01 | -0.077 | 0.21 | -0.364 | _ | 0.3656 |
| UKB-PPP_1_230164203_T_C | 6.04e-01 | -0.086 | 0.24 | -0.412 | _ | 0.3875 |
| UKB-PPP_1_25682168_T_C | 5.71e-01 | -0.112 | 0.276 | -0.5 | _ | 0.4846 |
| UKB-PPP_20_5600143_C_T | 4.73e-01 | -0.161 | 0.279 | -0.601 | _ | 0.0279 |
| UKB-PPP_14_49639327_A_G | 2.37e-01 | -0.167 | 0.109 | -0.443 | _ | 0.4606 |
| UKB-PPP_1_172556819_C_T | 3.17e-01 | -0.182 | 0.175 | -0.539 | _ | 0.4371 |
| UKB-PPP_4_105174348_A_C | 2.74e-01 | -0.234 | 0.185 | -0.654 | _ | 0.162 |
| UKB-PPP_12_21178691_C_T | 1.24e-01 | -0.246 | 0.068 | -0.56 | _ | 0.3863 |
| UKB-PPP_10_17833858_T_G | 1.93e-01 | -0.259 | 0.132 | -0.65 | _ | 0.339 |
| UKB-PPP_9_34592701_C_T | 2.52e-04 | -0.663 | -0.308 | -1.019 | _ | 0.1695 |
| **combined** | | | | | | |
| UKB-PPP | 2.29e-02 | 0.096 | 0.179 | 0.013 | 29 | _ |

### AU. IL1RL2_SCZ_Trans-pQTL_DECODE

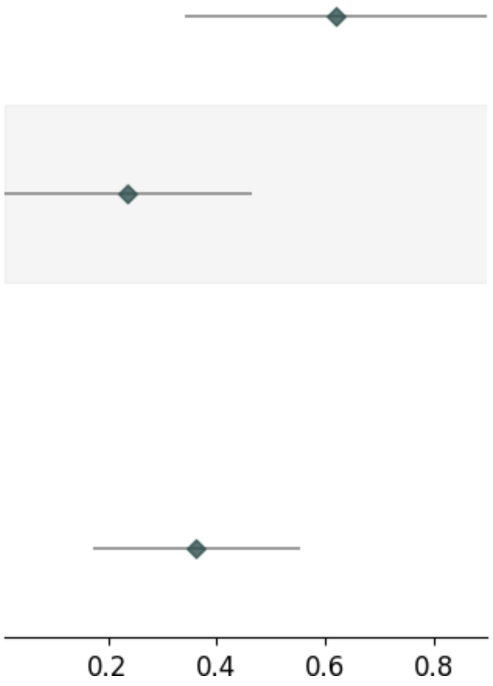


| Variable | Pvalue | Beta | Up_CI | Low_CI | nSNP | MAF |
|---|---|---|---|---|---|---|
| **Decode_SingleSNP** | | | | | | |
| Decode_12_120979185_A_G | 1.26e-05 | 0.621 | 0.899 | 0.342 | _ | 0.3158 |
| Decode_1_196695521_C_T | 4.07e-02 | 0.237 | 0.464 | 0.01 | _ | 0.4001 |
| **combined** | | | | | | |
| Decode | 1.97e-04 | 0.362 | 0.553 | 0.172 | 2 | _ |

### AV. ISL1_SCZ_Trans-pQTL

| Variable | Pvalue | Beta | Up_CI | Low_CI | nSNP | MAF |
|---|---|---|---|---|---|---|
| **deCODE_SingleSNP** | | | | | | |
| deCODE_3_52788905_C_T | 4.56e-14 | 1.015 | 1.279 | 0.752 | _ | 0.3908 |
| deCODE_1_196695521_C_T | 4.07e-02 | 0.237 | 0.464 | 0.01 | _ | 0.4001 |
| deCODE_10_7700504_A_G | 7.62e-01 | 0.049 | 0.362 | -0.265 | _ | 0.0659 |
| **combined** | | | | | | |
| deCODE | 8.58e-05 | 0.336 | 0.503 | 0.168 | 3 | _ |

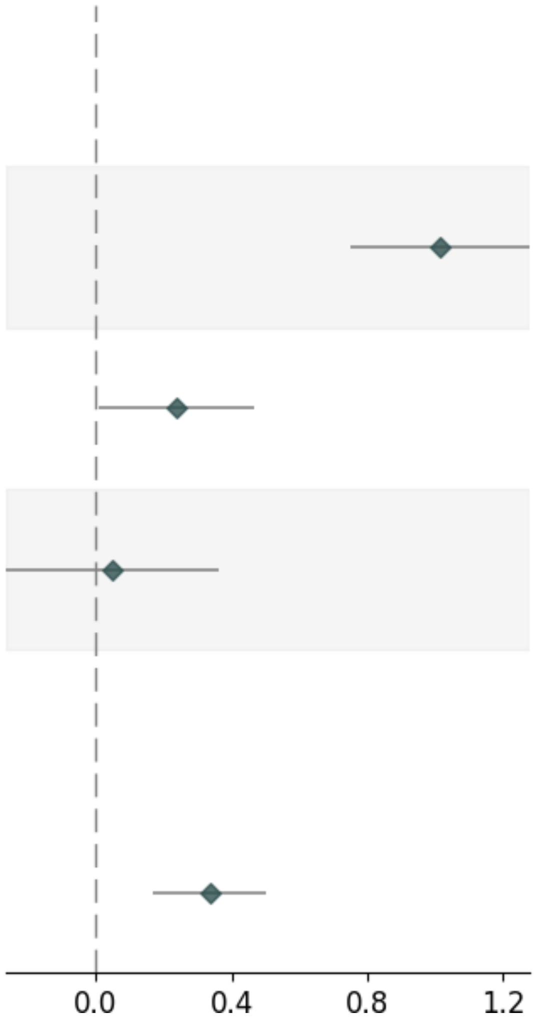

### AW. PHYKPL_SCZ_Trans-pQTL

| Variable | Pvalue | Beta | Up_CI | Low_CI | nSNP | MAF |
|---|---|---|---|---|---|---|
| **UKB-PPP_SingleSNP** | | | | | | |
| UKB-PPP_3_56844919_T_C | 1.54e-03 | -0.592 | -0.226 | -0.959 | _ | 0.3721 |
| UKB-PPP_3_52533601_A_G | 1.43e-17 | -1.353 | -1.042 | -1.663 | _ | 0.4611 |
| **combined** | | | | | | |
| UKB-PPP | 7.22e-08 | -0.894 | -0.569 | -1.22 | 2 | _ |

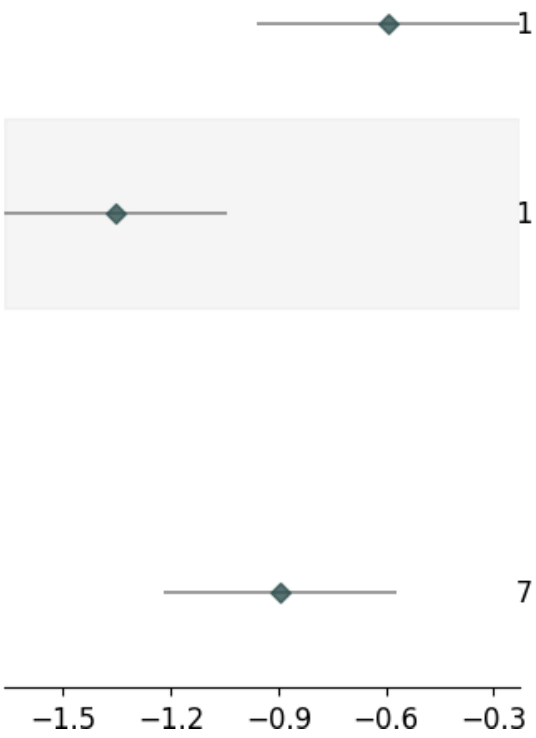

### AX. PI16_SCZ_Trans-pQTL

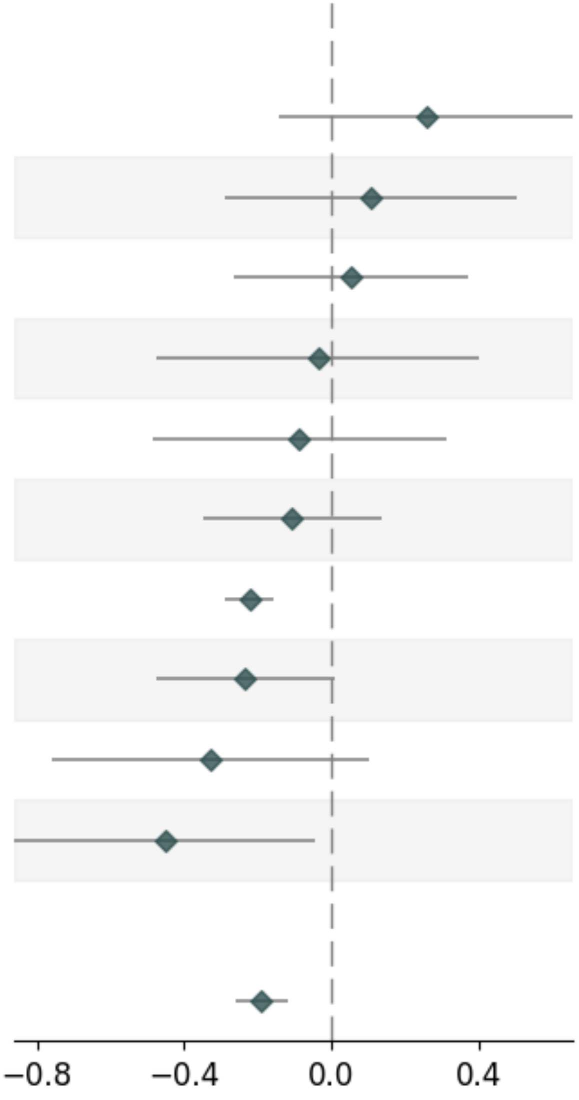


| Variable | Pvalue | Beta | Up_CI | Low_CI | nSNP | MAF |
|---|---|---|---|---|---|---|
| **UKB-PPP_SingleSNP** | | | | | | |
| UKB-PPP_3_132727060_C_A | 2.08e-01 | 0.257 | 0.657 | -0.143 | _ | 0.1576 |
| UKB-PPP_20_41117763_A_G | 5.97e-01 | 0.107 | 0.504 | -0.29 | _ | 0.3203 |
| UKB-PPP_2_27508073_T_C | 7.39e-01 | 0.054 | 0.373 | -0.265 | _ | 0.3914 |
| UKB-PPP_3_50407420_C_T | 8.72e-01 | -0.036 | 0.402 | -0.475 | _ | 0.0662 |
| UKB-PPP_1_219102578_A_G | 6.70e-01 | -0.087 | 0.312 | -0.485 | _ | 0.0272 |
| UKB-PPP_17_68883786_C_T | 3.85e-01 | -0.108 | 0.135 | -0.35 | _ | 0.0606 |
| UKB-PPP_3_52803839_T_C | 6.02e-11 | -0.222 | -0.156 | -0.289 | _ | 0.3422 |
| UKB-PPP_19_54255498_T_C | 5.88e-02 | -0.233 | 0.009 | -0.475 | _ | 0.3956 |
| UKB-PPP_2_162107791_G_A | 1.36e-01 | -0.327 | 0.103 | -0.758 | _ | 0.0602 |
| UKB-PPP_1_179557079_C_T | 3.07e-02 | -0.451 | -0.042 | -0.86 | _ | 0.038 |
| **combined** | | | | | | |
| UKB-PPP | 9.24e-08 | -0.19 | -0.12 | -0.26 | 10 | _ |

### AY. PSMD5_SCZ_Trans-pQTL

| Variable | Pvalue | Beta | Up_CI | Low_CI | nSNP | MAF |
|---|---|---|---|---|---|---|
| **deCODE_SingleSNP** | | | | | | |
| deCODE_3_52788840_T_C | 4.56e-14 | 1.144 | 1.441 | 0.847 | _ | 0.3908 |
| deCODE_1_196695521_C_T | 4.07e-02 | 0.304 | 0.594 | 0.013 | _ | 0.4001 |
| deCODE_7_95305261_G_A | 3.33e-01 | 0.174 | 0.527 | -0.179 | _ | 0.1555 |
| **combined** | | | | | | |
| deCODE | 3.73e-05 | 0.433 | 0.638 | 0.227 | 3 | _ |

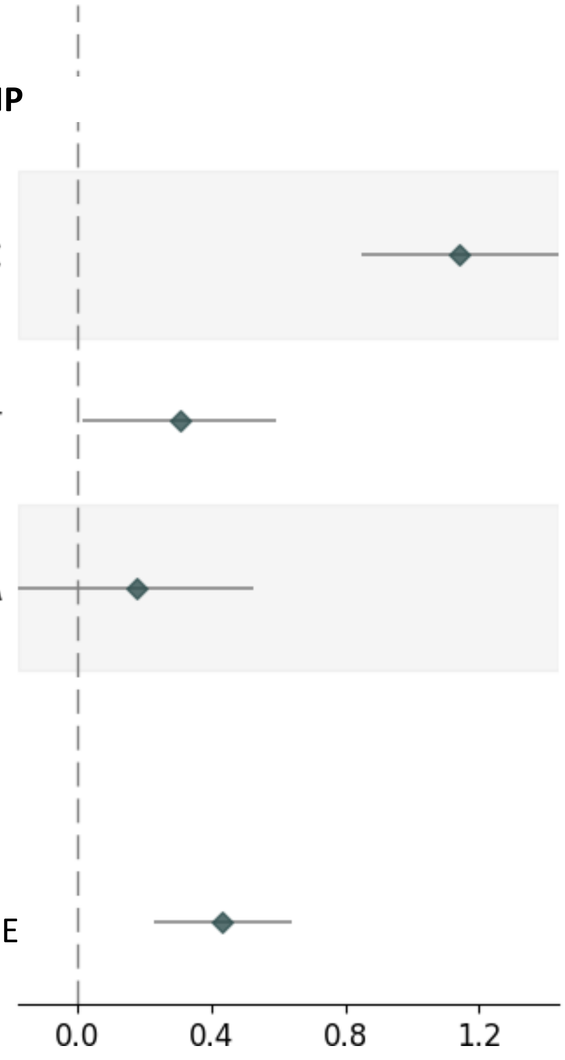

#### AZ. S100A16_SCZ_Trans-pQTL

| Variable | Pvalue | Beta | Up_CI | Low_CI | nSNP | MAF |
|---|---|---|---|---|---|---|
| **deCODE_SingleSNP** | | | | | | |
| deCODE_1_196695521_C_T | 4.07e-02 | 0.309 | 0.605 | 0.013 | _ | 0.4001 |
| deCODE_1_151836866_C_T | 4.24e-01 | -0.139 | 0.202 | -0.481 | _ | 0.4392 |
| deCODE_1_150812144_C_T | 4.00e-01 | -0.181 | 0.24 | -0.601 | _ | 0.0296 |
| deCODE_20_3719094_C_T | 4.68e-02 | -0.322 | -0.005 | -0.639 | _ | 0.0118 |
| **UKB-PPP_SingleSNP** | | | | | | |
| UKB-PPP_13_103049340_G_A | 5.61e-01 | 0.094 | 0.411 | -0.223 | _ | 0.011 |
| UKB-PPP_17_30318092_G_A | 6.86e-04 | -0.538 | -0.227 | -0.848 | _ | 0.3296 |
| UKB-PPP_19_48703417_G_A | 1.47e-06 | -0.558 | -0.331 | -0.785 | _ | 0.4907 |
| **combined** | | | | | | |
| deCODE | 7.02e-01 | -0.056 | 0.232 | -0.345 | 4 | _ |
| UKB-PPP | 3.96e-05 | -0.362 | -0.189 | -0.534 | 3 | _ |

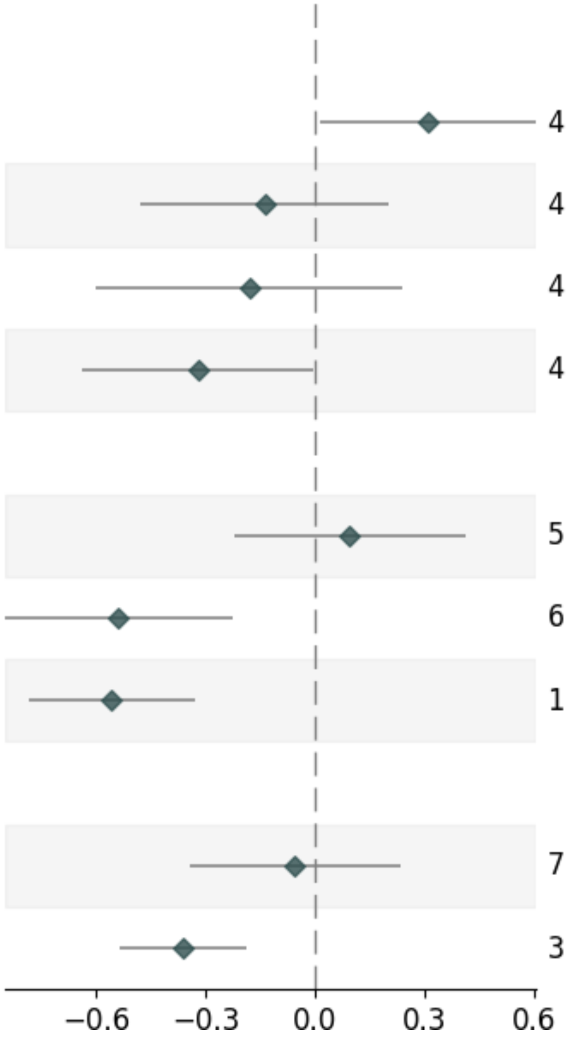

### BA. ANXA4_CTP_Trans-pQTL

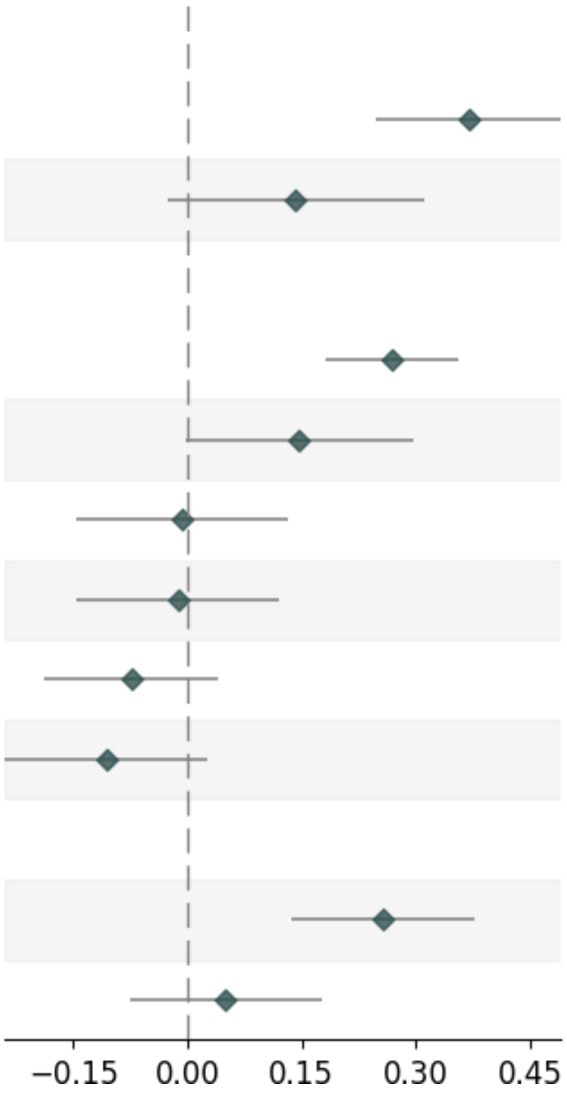


| Variable | Pvalue | Beta | Up_CI | Low_CI | nSNP | MAF |
|---|---|---|---|---|---|---|
| **deCODE_SingleSNP** | | | | | | |
| deCODE_10_63311455_C_A | 3.28e-09 | 0.369 | 0.491 | 0.247 | _ | 0.4348 |
| deCODE_9_132989126_C_T | 1.00e-01 | 0.141 | 0.31 | -0.027 | _ | 0.0163 |
| **UKB-PPP_SingleSNP** | | | | | | |
| UKB-PPP_10_63344740_A_G | 1.90e-09 | 0.269 | 0.356 | 0.181 | _ | 0.4235 |
| UKB-PPP_17_29518756_C_T | 5.45e-02 | 0.147 | 0.297 | -0.003 | _ | 0.4631 |
| UKB-PPP_11_180258_T_C | 9.25e-01 | -0.007 | 0.133 | -0.147 | _ | 0.2593 |
| UKB-PPP_5_132485305_G_A | 8.50e-01 | -0.013 | 0.121 | -0.147 | _ | 0.3881 |
| UKB-PPP_3_56815721_T_C | 2.07e-01 | -0.074 | 0.041 | -0.189 | _ | 0.3963 |
| UKB-PPP_19_55025227_G_A | 1.14e-01 | -0.107 | 0.026 | -0.24 | _ | 0.1663 |
| **combined** | | | | | | |
| deCODE | 2.75e-05 | 0.258 | 0.378 | 0.137 | 2 | _ |
| UKB-PPP | 4.41e-01 | 0.05 | 0.176 | -0.076 | 6 | _ |

### BB. CCL21_CTP_Trans-pQTL_UKB-PPP

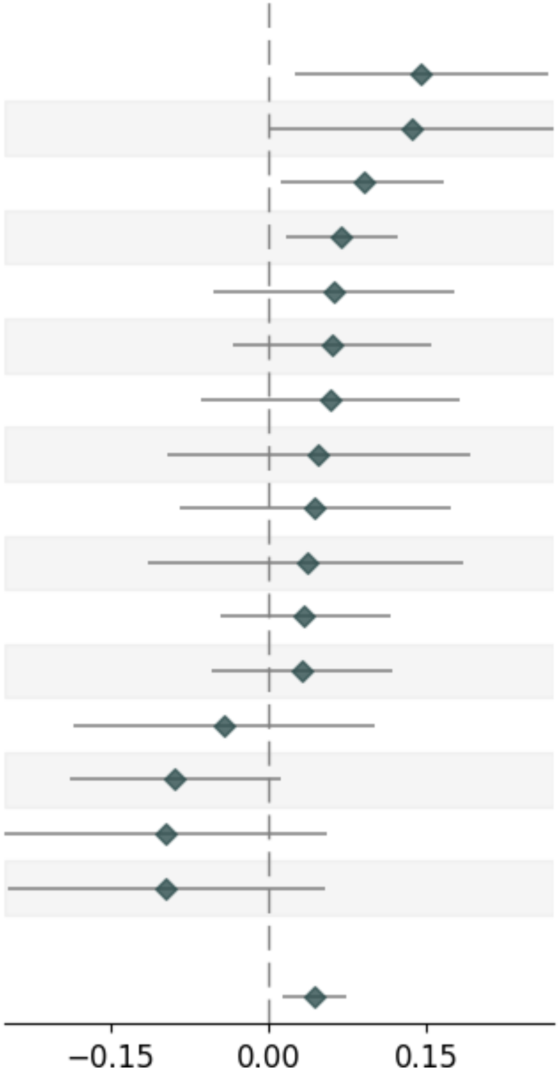


| Variable | Pvalue | Beta | Up_CI | Low_CI | nSNP | MAF |
|---|---|---|---|---|---|---|
| **UKB-PPP_SingleSNP** | | | | | | |
| UKB-PPP_16_54402053_T_G | 1.79e-02 | 0.145 | 0.265 | 0.025 | _ | 0.3435 |
| UKB-PPP_12_6384185_A_G | 4.71e-02 | 0.136 | 0.271 | 0.002 | _ | 0.2461 |
| UKB-PPP_12_111446804_T_C | 2.31e-02 | 0.09 | 0.167 | 0.012 | _ | 0.4827 |
| UKB-PPP_3_132521256_A_C | 1.09e-02 | 0.069 | 0.123 | 0.016 | _ | 0.1123 |
| UKB-PPP_16_90059414_C_T | 2.85e-01 | 0.062 | 0.177 | -0.052 | _ | 0.071 |
| UKB-PPP_3_56815721_T_C | 2.07e-01 | 0.061 | 0.155 | -0.034 | _ | 0.3959 |
| UKB-PPP_18_59429782_T_C | 3.46e-01 | 0.059 | 0.182 | -0.064 | _ | 0.3663 |
| UKB-PPP_2_65344150_A_C | 5.18e-01 | 0.047 | 0.191 | -0.096 | _ | 0.2744 |
| UKB-PPP_5_35883632_T_C | 5.06e-01 | 0.044 | 0.173 | -0.085 | _ | 0.2731 |
| UKB-PPP_19_33260733_C_T | 6.40e-01 | 0.036 | 0.185 | -0.114 | _ | 0.3362 |
| UKB-PPP_5_128021834_C_T | 4.00e-01 | 0.034 | 0.115 | -0.046 | _ | 0.2486 |
| UKB-PPP_22_30133642_C_A | 4.74e-01 | 0.031 | 0.117 | -0.054 | _ | 0.4432 |
| UKB-PPP_20_10669113_C_T | 5.66e-01 | -0.042 | 0.101 | -0.185 | _ | 0.2058 |
| UKB-PPP_5_115995638_A_G | 8.19e-02 | -0.089 | 0.011 | -0.189 | _ | 0.3949 |
| UKB-PPP_13_28029870_T_C | 2.13e-01 | -0.097 | 0.056 | -0.25 | _ | 0.0129 |
| UKB-PPP_16_85979737_G_A | 2.08e-01 | -0.097 | 0.054 | -0.247 | _ | 0.1317 |
| **combined** | | | | | | |
| UKB-PPP | 4.66e-03 | 0.043 | 0.074 | 0.013 | 16 | _ |

### BC. CCL21_CTP_Trans-pQTL_DECODE

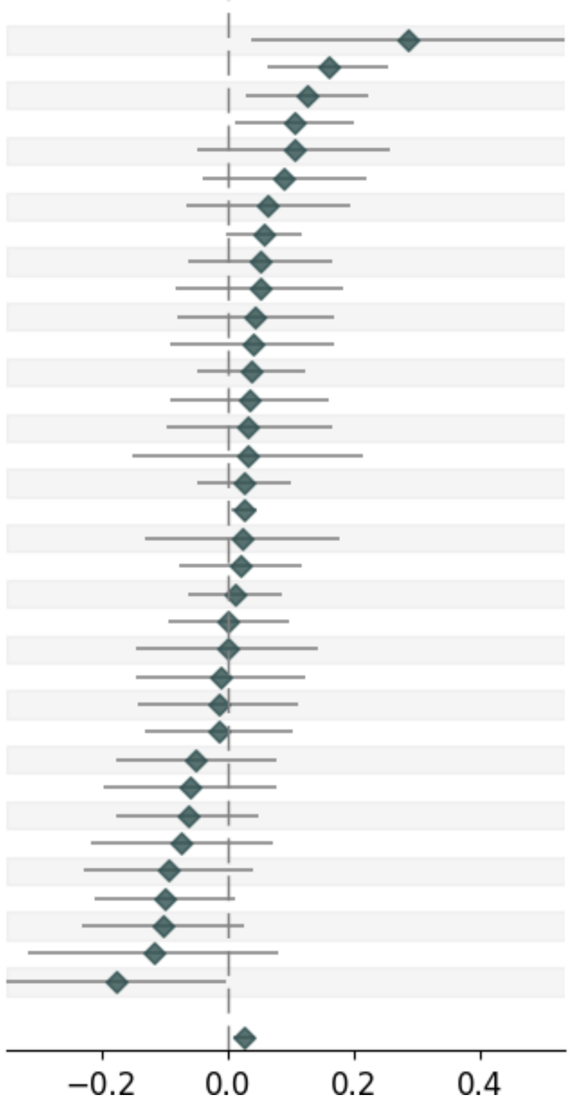


| Variable | Pvalue | Beta | Up_CI | Low_CI | nSNP | MAF |
|---|---|---|---|---|---|---|
| deCODE_SingleSNP | | | | | | |
| deCODE_3_131404591_C_T | 2.37e-02 | 0.287 | 0.535 | 0.038 | _ | 0.0151 |
| deCODE_9_33684438_C_T | 1.15e-03 | 0.159 | 0.254 | 0.063 | _ | 0.1032 |
| deCODE_12_111472415_A_G | 9.97e-03 | 0.127 | 0.223 | 0.03 | _ | 0.3866 |
| deCODE_3_130359542_C_T | 2.65e-02 | 0.106 | 0.199 | 0.012 | _ | 0.0926 |
| deCODE_3_134303269_A_G | 1.78e-01 | 0.105 | 0.258 | -0.048 | _ | 0.0303 |
| deCODE_3_133480468_A_G | 1.81e-01 | 0.089 | 0.219 | -0.041 | _ | 0.0133 |
| deCODE_20_44032864_T_C | 3.33e-01 | 0.064 | 0.194 | -0.066 | _ | 0.3415 |
| deCODE_3_133230589_A_G | 5.80e-02 | 0.058 | 0.118 | -0.002 | _ | 0.0821 |
| deCODE_3_133068151_C_A | 3.72e-01 | 0.052 | 0.167 | -0.062 | _ | 0.0064 |
| deCODE_18_59419702_G_A | 4.54e-01 | 0.051 | 0.183 | -0.082 | _ | 0.3554 |
| deCODE_5_35885880_T_C | 4.94e-01 | 0.044 | 0.168 | -0.081 | _ | 0.328 |
| deCODE_3_135069173_T_G | 5.47e-01 | 0.04 | 0.17 | -0.09 | _ | 0.0175 |
| deCODE_5_128021834_C_T | 4.00e-01 | 0.037 | 0.123 | -0.049 | _ | 0.1895 |
| deCODE_3_129955529_G_T | 5.79e-01 | 0.036 | 0.161 | -0.09 | _ | 0.1869 |
| deCODE_3_137718840_C_T | 6.18e-01 | 0.033 | 0.165 | -0.098 | _ | 0.0077 |
| deCODE_3_127974580_A_G | 7.35e-01 | 0.032 | 0.214 | -0.151 | _ | 0.0125 |
| deCODE_22_30107182_G_A | 4.69e-01 | 0.027 | 0.101 | -0.047 | _ | 0.423 |
| deCODE_3_132404638_T_C | 7.86e-03 | 0.027 | 0.047 | 0.007 | _ | 0.1254 |
| deCODE_3_129513411_G_A | 7.70e-01 | 0.023 | 0.176 | -0.13 | _ | 0.0383 |
| deCODE_3_134855811_G_A | 6.90e-01 | 0.02 | 0.118 | -0.078 | _ | 0.0068 |
| deCODE_3_132099751_G_A | 7.61e-01 | 0.012 | 0.086 | -0.063 | _ | 0.0467 |
| deCODE_2_65363202_G_A | 9.89e-01 | 0.001 | 0.097 | -0.095 | _ | 0.3658 |
| deCODE_3_132143092_A_G | 9.95e-01 | -0.0 | 0.144 | -0.145 | _ | 0.3199 |
| deCODE_11_75358827_G_T | 8.78e-01 | -0.01 | 0.124 | -0.144 | _ | 0.4955 |
| deCODE_13_28109027_C_T | 8.25e-01 | -0.014 | 0.112 | -0.141 | _ | 0.2206 |
| deCODE_3_133510799_G_A | 8.14e-01 | -0.014 | 0.103 | -0.131 | _ | 0.047 |
| deCODE_3_125192997_C_T | 4.30e-01 | -0.051 | 0.076 | -0.178 | _ | 0.2921 |
| deCODE_20_10672061_G_A | 3.85e-01 | -0.06 | 0.076 | -0.197 | _ | 0.2414 |
| deCODE_3_133699224_G_A | 2.75e-01 | -0.063 | 0.05 | -0.176 | _ | 0.0134 |
| deCODE_3_133854951_T_C | 3.22e-01 | -0.073 | 0.072 | -0.217 | _ | 0.0076 |
| deCODE_3_134476980_T_C | 1.68e-01 | -0.094 | 0.04 | -0.227 | _ | 0.0107 |
| deCODE_3_134416433_T_C | 8.20e-02 | -0.099 | 0.013 | -0.212 | _ | 0.0229 |
| deCODE_17_49376575_A_C | 1.14e-01 | -0.103 | 0.025 | -0.231 | _ | 0.2056 |
| deCODE_3_131509936_G_A | 2.45e-01 | -0.118 | 0.081 | -0.316 | _ | 0.0118 |
| deCODE_3_134305900_T_G | 4.74e-02 | -0.176 | -0.002 | -0.35 | _ | 0.0098 |
| **combined** | | | | | | |
| deCODE | 2.87e-03 | 0.025 | 0.042 | 0.009 | 35 | _ |

deCODE

#### BD. CD4_CTP_Trans-pQTL

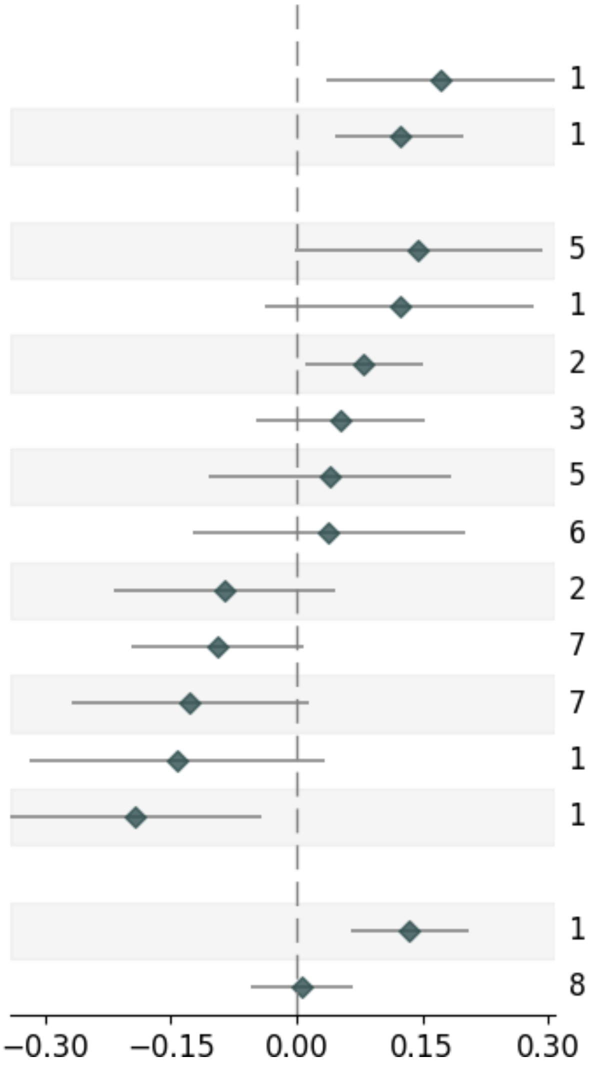


| Variable | Pvalue | Beta | Up_CI | Low_CI | nSNP | MAF |
|---|---|---|---|---|---|---|
| **Decode_SingleSNP** | | | | | | |
| Decode_12_111494996_C_T | 1.31e-02 | 0.173 | 0.309 | 0.036 | _ | 0.3805 |
| Decode_1_196695521_C_T | 1.52e-03 | 0.124 | 0.2 | 0.047 | _ | 0.4001 |
| **UKB-PPP_SingleSNP** | | | | | | |
| UKB-PPP_1_109247173_C_T | 5.46e-02 | 0.145 | 0.293 | -0.003 | _ | 0.1552 |
| UKB-PPP_17_43797377_C_A | 1.33e-01 | 0.123 | 0.283 | -0.037 | _ | 0.0288 |
| UKB-PPP_12_111446804_T_C | 2.31e-02 | 0.08 | 0.15 | 0.011 | _ | 0.4826 |
| UKB-PPP_11_116791110_T_C | 3.06e-01 | 0.052 | 0.153 | -0.048 | _ | 0.0581 |
| UKB-PPP_1_199005775_C_T | 5.89e-01 | 0.04 | 0.184 | -0.104 | _ | 0.2443 |
| UKB-PPP_14_94378610_C_T | 6.45e-01 | 0.038 | 0.201 | -0.124 | _ | 0.0207 |
| UKB-PPP_8_19966981_T_C | 2.04e-01 | -0.086 | 0.047 | -0.219 | _ | 0.2872 |
| UKB-PPP_19_44911142_C_A | 7.44e-02 | -0.094 | 0.009 | -0.198 | _ | 0.1095 |
| UKB-PPP_7_72841636_A_G | 7.89e-02 | -0.127 | 0.015 | -0.269 | _ | 0.078 |
| UKB-PPP_11_116652851_A_C | 1.14e-01 | -0.142 | 0.034 | -0.318 | _ | 0.009 |
| UKB-PPP_8_78696019_A_T | 1.23e-02 | -0.192 | -0.042 | -0.341 | _ | 0.3104 |
| **combined** | | | | | | |
| Decode | 1.89e-04 | 0.135 | 0.206 | 0.064 | 2 | _ |
| UKB-PPP | 8.31e-01 | 0.007 | 0.068 | -0.054 | 11 | _ |

#### BE. CMPK1_CTP_Trans-pQTL

| Variable | Pvalue | Beta | Up_CI | Low_CI | nSNP | MAF |
|---|---|---|---|---|---|---|
| **deCODE_SingleSNP** | | | | | | |
| deCODE_10_63311455_C_A | 3.28e-09 | 0.389 | 0.518 | 0.26 | _ | 0.4348 |
| deCODE_1_196705140_C_T | 3.30e-03 | 0.187 | 0.312 | 0.062 | _ | 0.4315 |
| **combined** | | | | | | |
| deCODE | 3.34e-06 | 0.262 | 0.372 | 0.151 | 2 | _ |

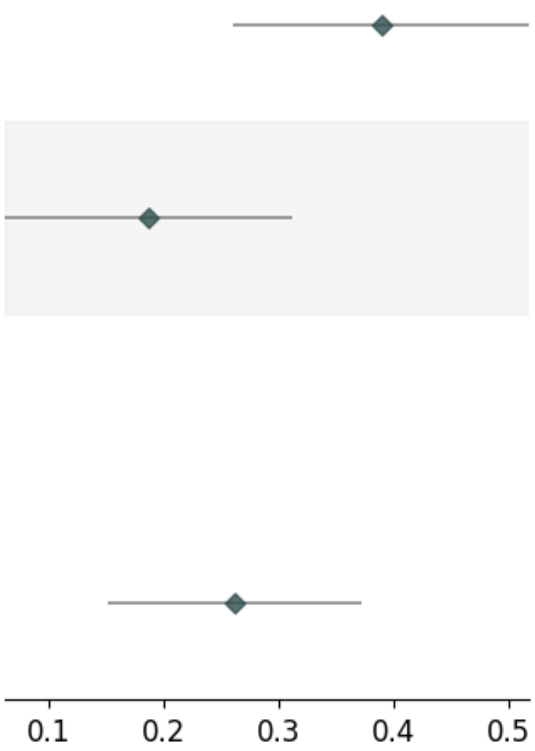

#### BF. EFNA4_CTP_Trans-pQTL

| Variable | Pvalue | Beta | Up_CI | Low_CI | nSNP | MAF |
|---|---|---|---|---|---|---|
| **deCODE_SingleSNP** | | | | | | |
| deCODE_16_370723_T_G | 1.35e-03 | 0.218 | 0.351 | 0.085 | _ | 0.4024 |
| deCODE_196695521_C_T | 1.52e-03 | 0.169 | 0.273 | 0.065 | _ | 0.4001 |
| **UKB-PPP_SingleSNP** | | | | | | |
| UKB-PPP_9_133274295_A_T | 2.79e-02 | 0.189 | 0.357 | 0.021 | _ | 0.1842 |
| UKB-PPP_16_363747_A_G | 5.52e-04 | 0.177 | 0.277 | 0.076 | _ | 0.459 |
| UKB-PPP_12_111446804_T_C | 2.31e-02 | 0.114 | 0.212 | 0.016 | _ | 0.4831 |
| UKB-PPP_16_20345959_C_T | 6.58e-01 | 0.024 | 0.13 | -0.082 | _ | 0.1665 |
| UKB-PPP_7_151717243_T_C | 9.24e-01 | 0.007 | 0.159 | -0.144 | _ | 0.2872 |
| UKB-PPP_2_221522212_T_C | 9.41e-01 | -0.004 | 0.098 | -0.106 | _ | 0.4869 |
| UKB-PPP_19_43648948_A_G | 2.06e-01 | -0.062 | 0.034 | -0.157 | _ | 0.156 |
| UKB-PPP_4_76480299_C_T | 3.54e-01 | -0.062 | 0.069 | -0.193 | _ | 0.4568 |
| **combined** | | | | | | |
| deCODE | 6.63e-05 | 0.186 | 0.278 | 0.095 | 2 | _ |
| UKB-PPP | 2.42e-01 | 0.039 | 0.105 | -0.026 | 8 | _ |

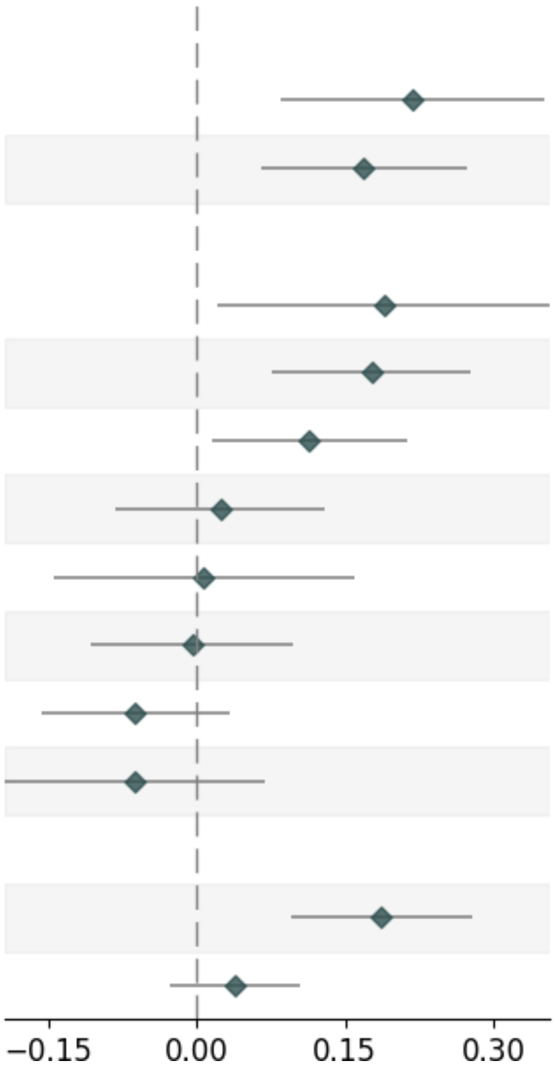

### BG. EREG_CTP_Trans-pQTL

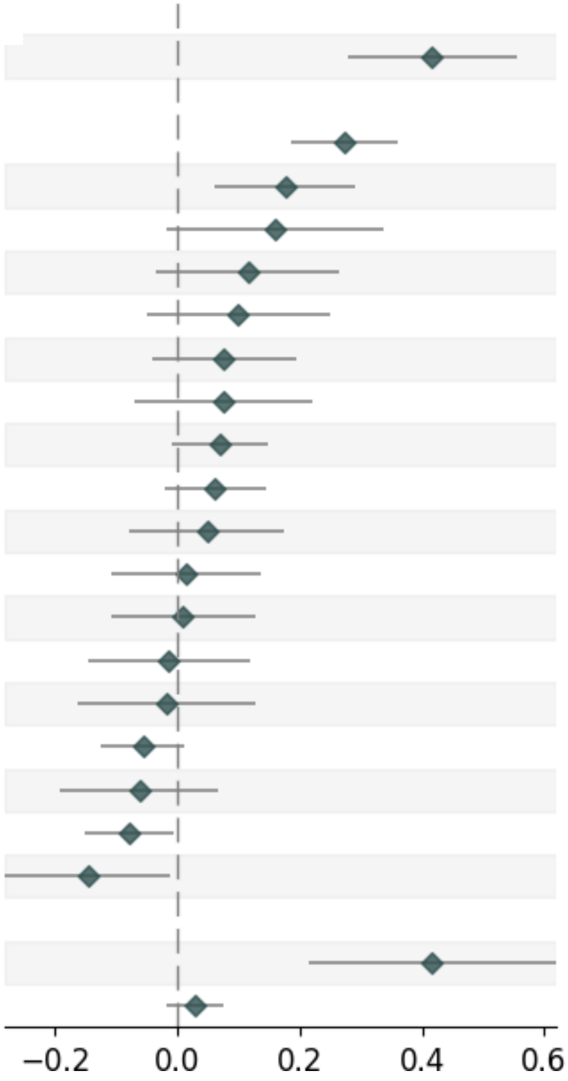

| Variable | Pvalue | Beta | Up_CI | Low_CI | nSNP | MAF |
|---|---|---|---|---|---|---|
| **deCODE_SingleSNP** | | | | | | |
| deCODE_10_63311455_C_A | 3.28e-09 | 0.417 | 0.555 | 0.279 | _ | 0.4348 |
| **UKB-PPP_SingleSNP** | | | | | | |
| UKB-PPP_10_63288546_C_A | 8.00e-10 | 0.273 | 0.36 | 0.186 | _ | 0.4119 |
| UKB-PPP_20_19300647_C_T | 2.62e-03 | 0.176 | 0.29 | 0.061 | _ | 0.3539 |
| UKB-PPP_15_64868190_T_A | 7.43e-02 | 0.161 | 0.337 | -0.016 | _ | 0.1745 |
| UKB-PPP_20_45845988_C_T | 1.30e-01 | 0.115 | 0.265 | -0.034 | _ | 0.009 |
| UKB-PPP_8_108643441_G_A | 1.87e-01 | 0.1 | 0.249 | -0.049 | _ | 0.0223 |
| UKB-PPP_4_101862194_C_T | 2.01e-01 | 0.077 | 0.196 | -0.041 | _ | 0.398 |
| UKB-PPP_9_4746539_C_T | 3.01e-01 | 0.077 | 0.222 | -0.069 | _ | 0.4944 |
| UKB-PPP_9_96430637_A_C | 8.73e-02 | 0.069 | 0.149 | -0.01 | _ | 0.2296 |
| UKB-PPP_5_151455788_A_G | 1.36e-01 | 0.062 | 0.144 | -0.02 | _ | 0.3841 |
| UKB-PPP_14_92750678_G_A | 4.44e-01 | 0.049 | 0.175 | -0.077 | _ | 0.0376 |
| UKB-PPP_3_123114156_C_T | 8.00e-01 | 0.016 | 0.138 | -0.106 | _ | 0.407 |
| UKB-PPP_16_81837364_C_T | 8.67e-01 | 0.01 | 0.129 | -0.108 | _ | 0.1381 |
| UKB-PPP_1_171980610_T_C | 8.39e-01 | -0.014 | 0.118 | -0.145 | _ | 0.1964 |
| UKB-PPP_10_49040879_G_A | 8.13e-01 | -0.018 | 0.128 | -0.163 | _ | 0.2389 |
| UKB-PPP_19_55027704_T_G | 1.14e-01 | -0.055 | 0.013 | -0.124 | _ | 0.166 |
| UKB-PPP_10_71847568_C_T | 3.44e-01 | -0.062 | 0.067 | -0.191 | _ | 0.3972 |
| UKB-PPP_8_105569300_A_T | 3.24e-02 | -0.078 | -0.007 | -0.15 | _ | 0.2865 |
| UKB-PPP_3_58294963_A_G | 3.32e-02 | -0.146 | -0.012 | -0.281 | _ | 0.3245 |
| **combined** | | | | | | |
| deCODE | 5.50e-05 | 0.417 | 0.62 | 0.214 | 1 | _ |
| UKB-PPP | 2.09e-01 | 0.03 | 0.077 | -0.017 | 18 | _ |

### BH. F12_CTP_Trans-pQTL

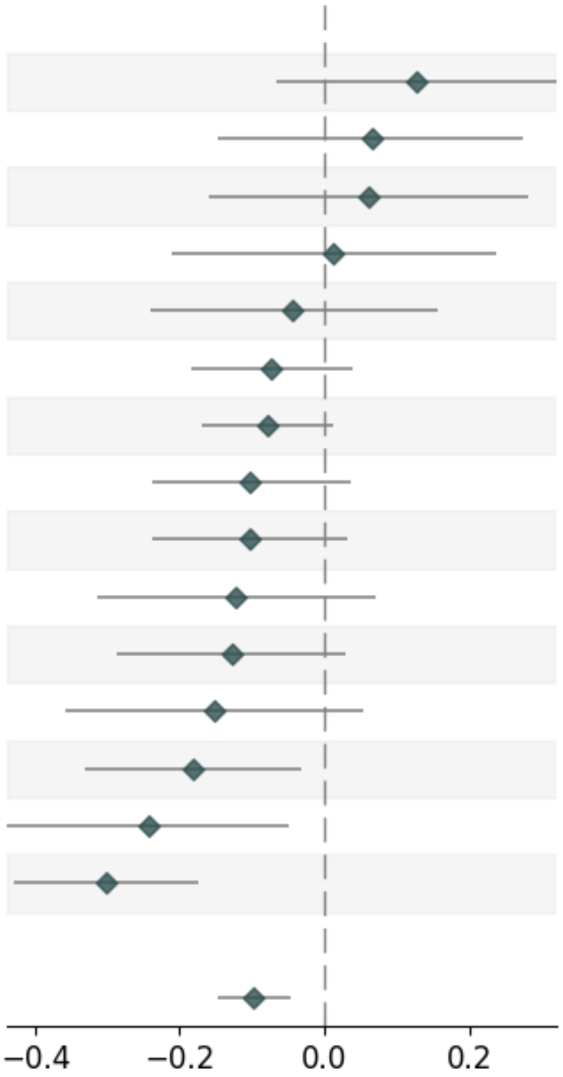


| Variable | Pvalue | Beta | Up_CI | Low_CI | nSNP | MAF |
|---|---|---|---|---|---|---|
| **UKB-PPP_SingleSNP** | | | | | | |
| UKB-PPP_12_123398859_G_A | 1.93e-01 | 0.128 | 0.322 | -0.065 | _ | 0.0937 |
| UKB-PPP_22_43928975_G_A | 5.47e-01 | 0.065 | 0.275 | -0.146 | _ | 0.2154 |
| UKB-PPP_22_49911450_T_A | 5.86e-01 | 0.061 | 0.281 | -0.159 | _ | 0.0599 |
| UKB-PPP_16_11677156_C_T | 9.13e-01 | 0.012 | 0.237 | -0.212 | _ | 0.0182 |
| UKB-PPP_6_130093090_A_G | 6.73e-01 | -0.043 | 0.156 | -0.241 | _ | 0.3126 |
| UKB-PPP_5_176219714_C_T | 2.00e-01 | -0.073 | 0.038 | -0.183 | _ | 0.0784 |
| UKB-PPP_2_27508073_T_C | 8.78e-02 | -0.079 | 0.012 | -0.169 | _ | 0.3916 |
| UKB-PPP_19_54255498_T_C | 1.46e-01 | -0.102 | 0.036 | -0.239 | _ | 0.3952 |
| UKB-PPP_5_175834047_G_A | 1.29e-01 | -0.104 | 0.031 | -0.239 | _ | 0.2136 |
| UKB-PPP_19_795457_A_G | 2.15e-01 | -0.122 | 0.071 | -0.314 | _ | 0.359 |
| UKB-PPP_5_175687038_G_A | 1.13e-01 | -0.128 | 0.03 | -0.286 | _ | 0.007 |
| UKB-PPP_8_125494452_G_A | 1.52e-01 | -0.151 | 0.055 | -0.357 | _ | 0.391 |
| UKB-PPP_7_73611821_A_T | 1.76e-02 | -0.182 | -0.032 | -0.332 | _ | 0.1271 |
| UKB-PPP_16_72187565_A_G | 1.45e-02 | -0.243 | -0.048 | -0.438 | _ | 0.1095 |
| UKB-PPP_10_63431885_G_T | 4.22e-06 | -0.301 | -0.173 | -0.429 | _ | 0.499 |
| **combined** | | | | | | |
| UKB-PPP | 1.72e-04 | -0.097 | -0.046 | -0.147 | 15 | _ |

### BI. MYC_CTP_Trans-pQTL

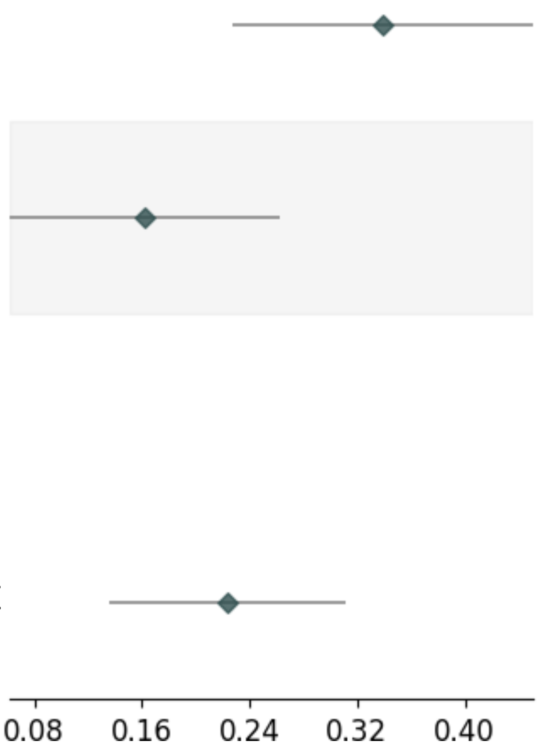


| Variable | Pvalue | Beta | Up_CI | Low_CI | nSNP | MAF |
|---|---|---|---|---|---|---|
| **deCODE_SingleSNP** | | | | | | |
| deCODE_10_63311455_C_A | 3.28e-09 | 0.339 | 0.451 | 0.227 | _ | 0.4348 |
| deCODE_1_196677131_A_G | 1.55e-03 | 0.162 | 0.263 | 0.062 | _ | 0.3973 |
| **combined** | | | | | | |
| deCODE | 5.67e-07 | 0.224 | 0.312 | 0.136 | 2 | _ |

### BJ. SYT17_CTP_Trans-pQTL

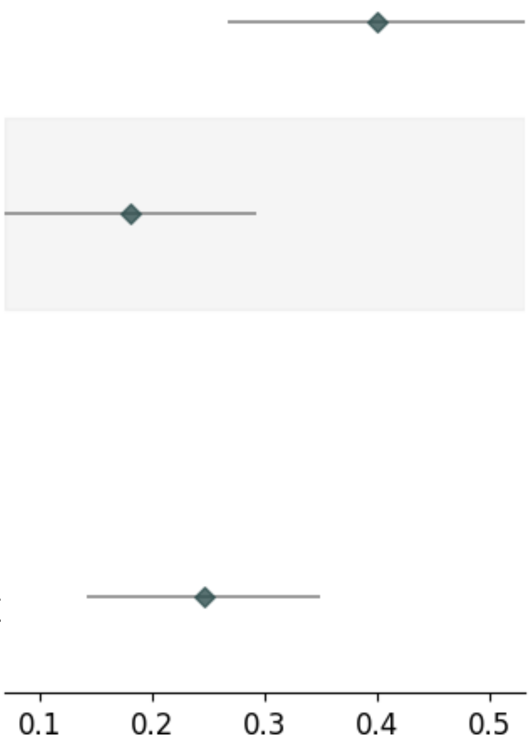


| Variable | Pvalue | Beta | Up_CI | Low_CI | nSNP | MAF |
|---|---|---|---|---|---|---|
| **deCODE_SingleSNP** | | | | | | |
| deCODE_10_63311455_C_A | 3.28e-09 | 0.4 | 0.532 | 0.267 | _ | 0.4348 |
| deCODE_1_196677131_A_G | 1.55e-03 | 0.181 | 0.292 | 0.069 | _ | 0.3973 |
| **combined** | | | | | | |
| deCODE | 3.36e-06 | 0.246 | 0.349 | 0.142 | 2 | _ |

### BK. TNS2_CTP_Trans-pQTL

| Variable | Pvalue | Beta | Up_CI | Low_CI | nSNP | MAF |
|---|---|---|---|---|---|---|
| **deCODE_SingleSNP** | | | | | | |
| deCODE_1_196695521_C_T | 1.52e-03 | 0.196 | 0.317 | 0.075 | _ | 0.4001 |
| deCODE_3_49684923_G_A | 7.12e-19 | -0.307 | -0.239 | -0.374 | _ | 0.3072 |
| **combined** | | | | | | |
| deCODE | 5.90e-06 | -0.166 | -0.094 | -0.237 | 2 | _ |

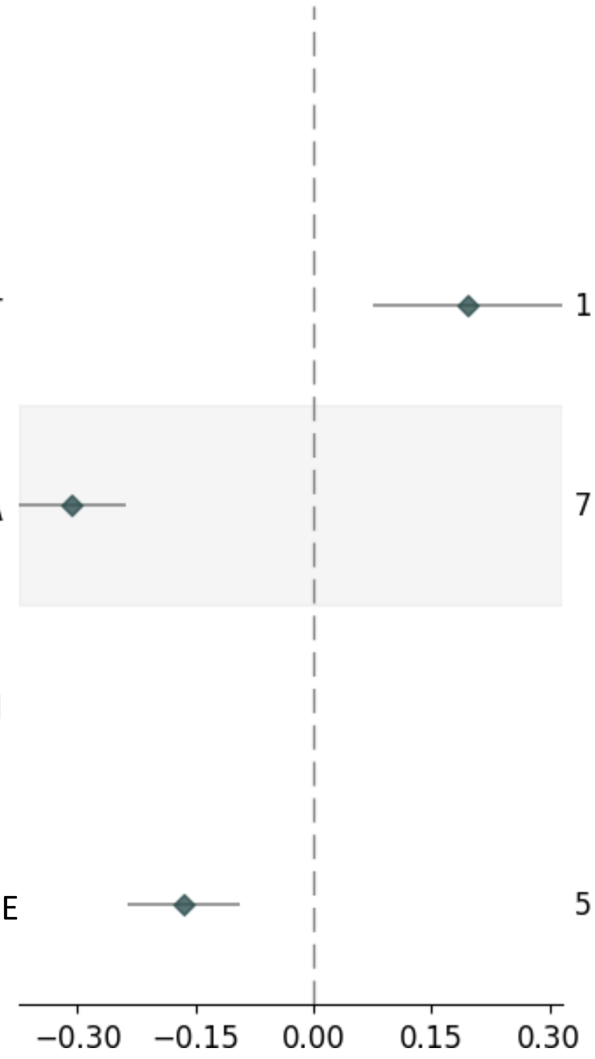
