## Supplementary figures and images for "Large-Scale Mendelian Randomization Study Reveals Circulating Blood-based Proteomic Biomarkers for Psychopathology and Cognitive Task Performance"

### Fig1_MR.tiff

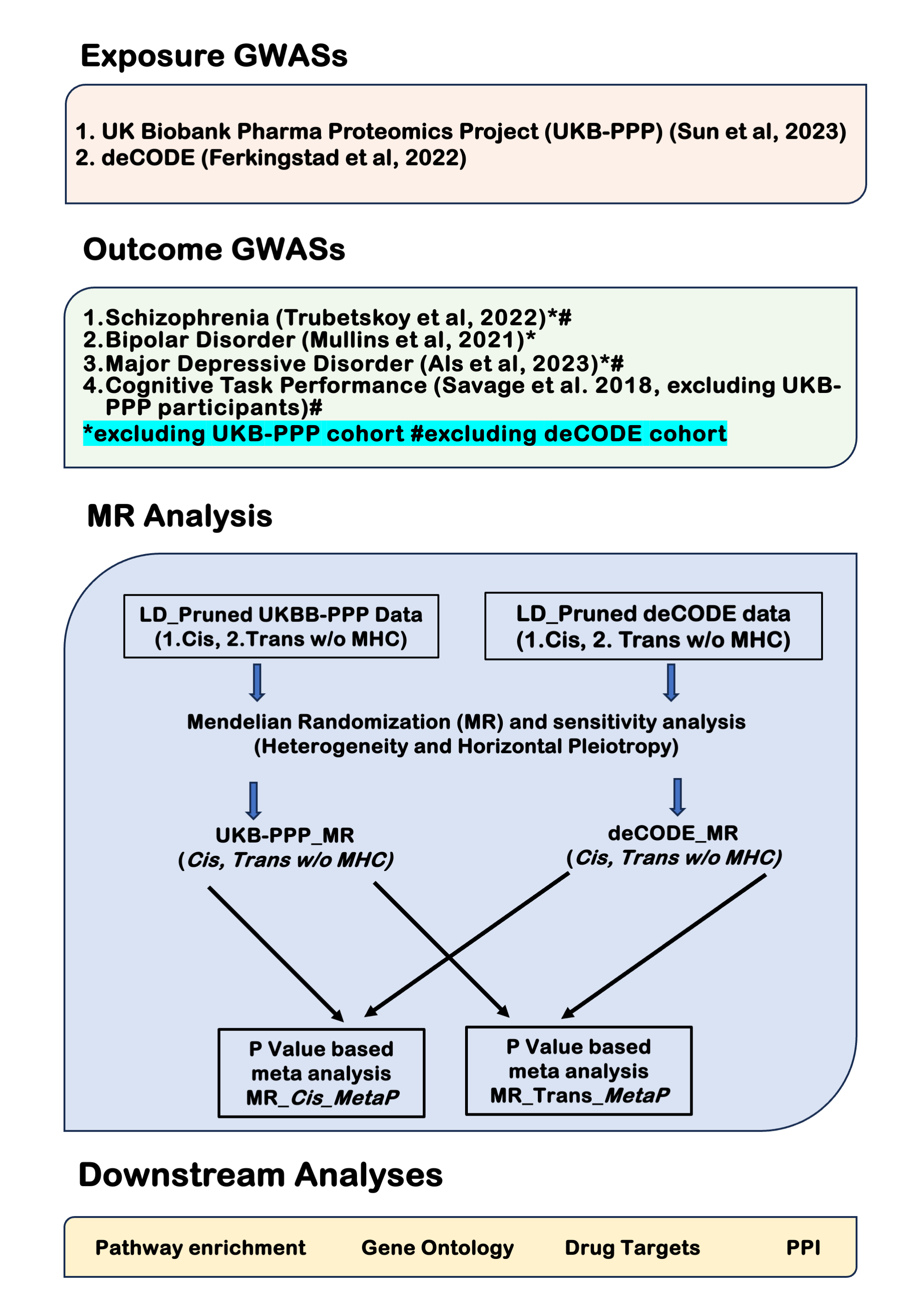

### Figure_2_MR.tiff

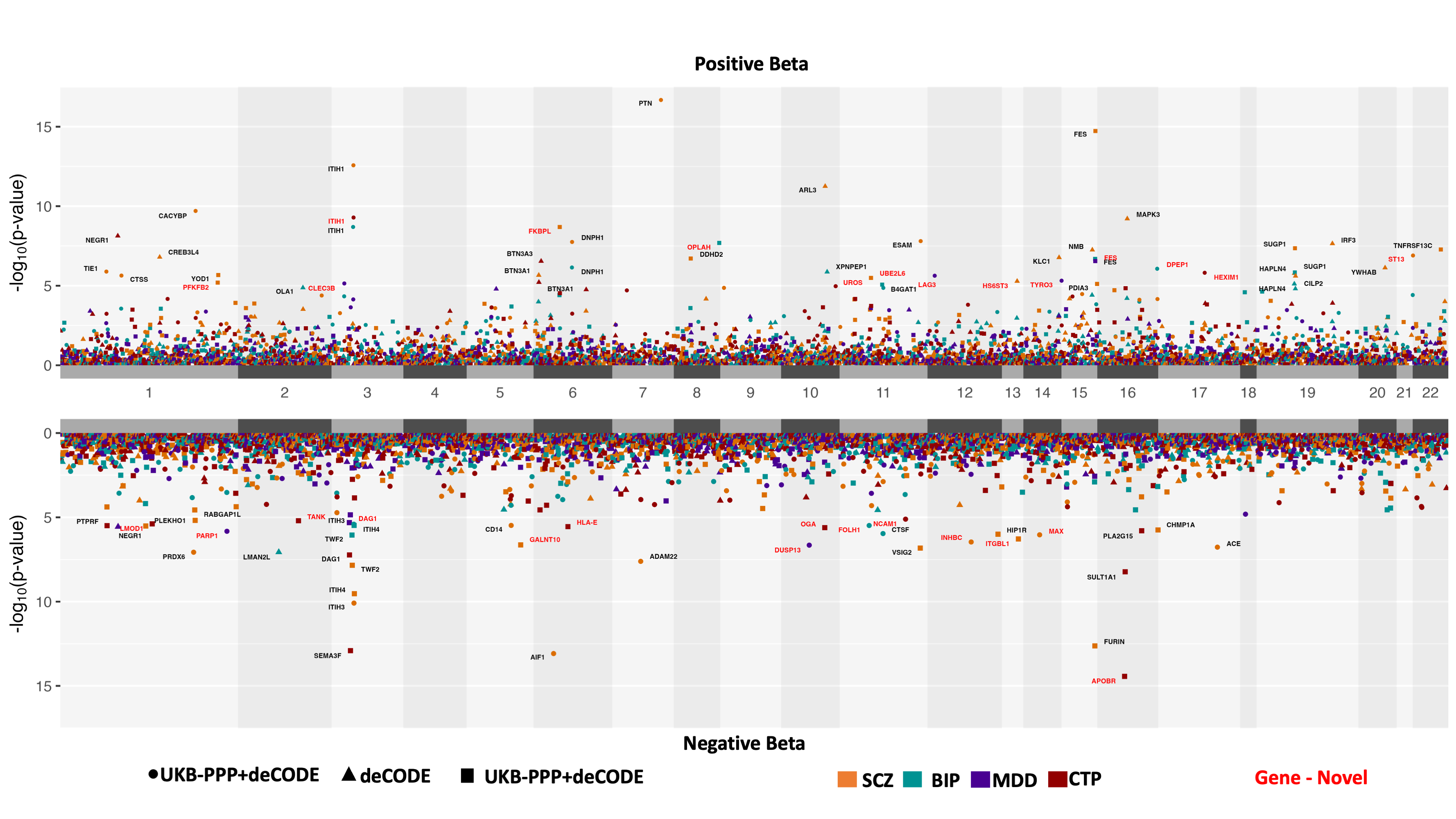

### Figure_3_MR.tiff

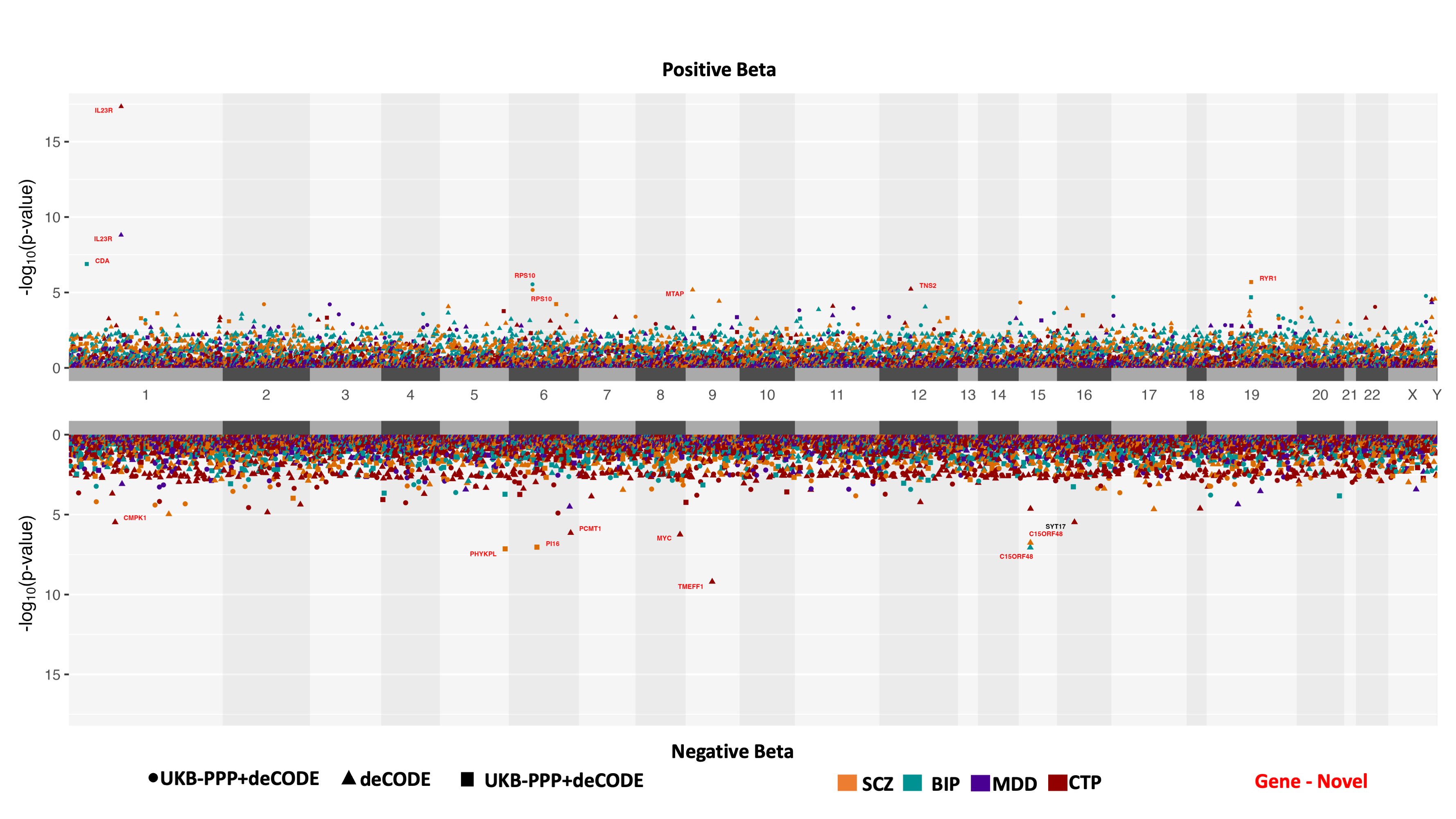

### Figure_4A_MR.tiff

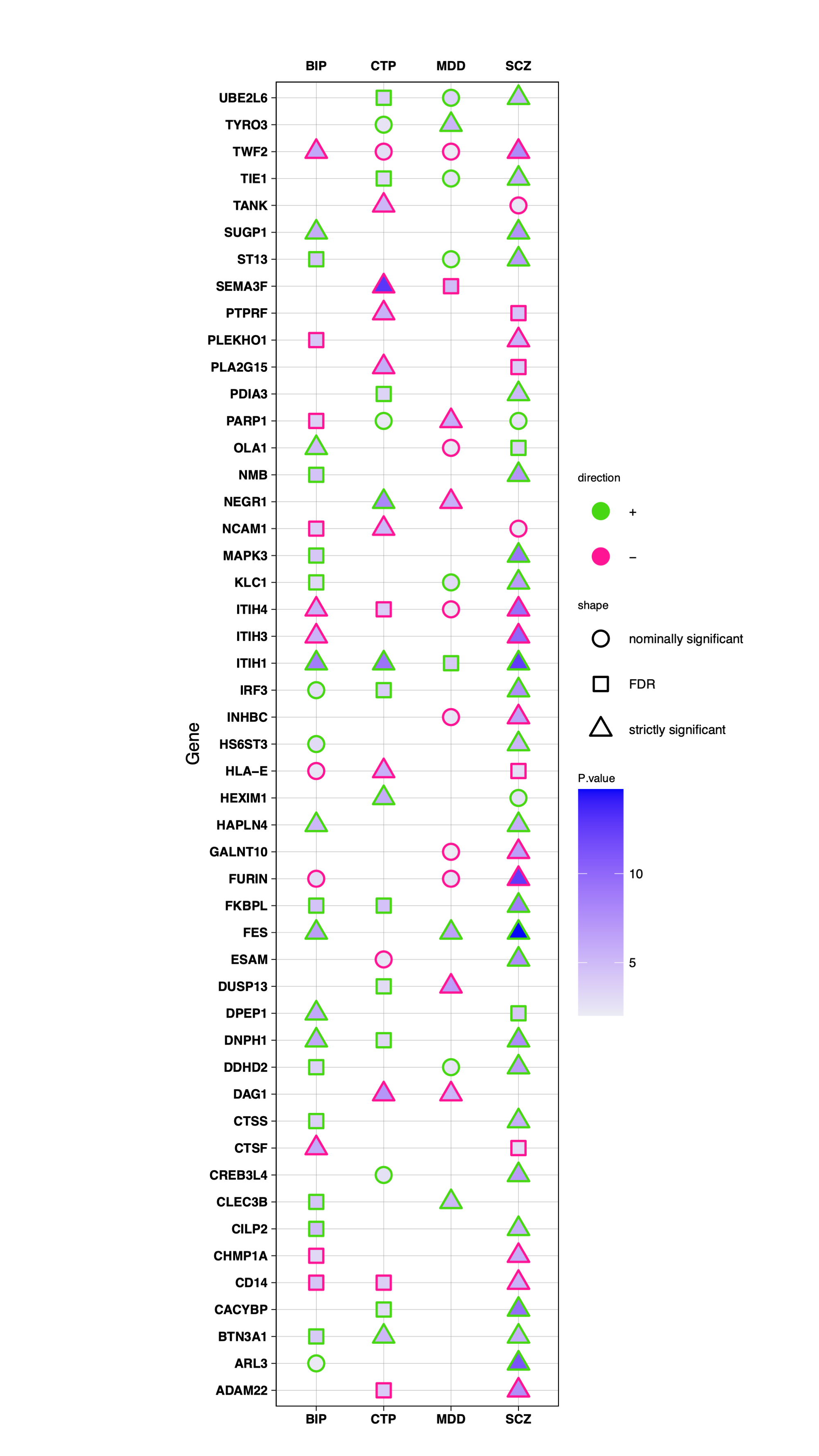

### Figure_4B_MR.tiff

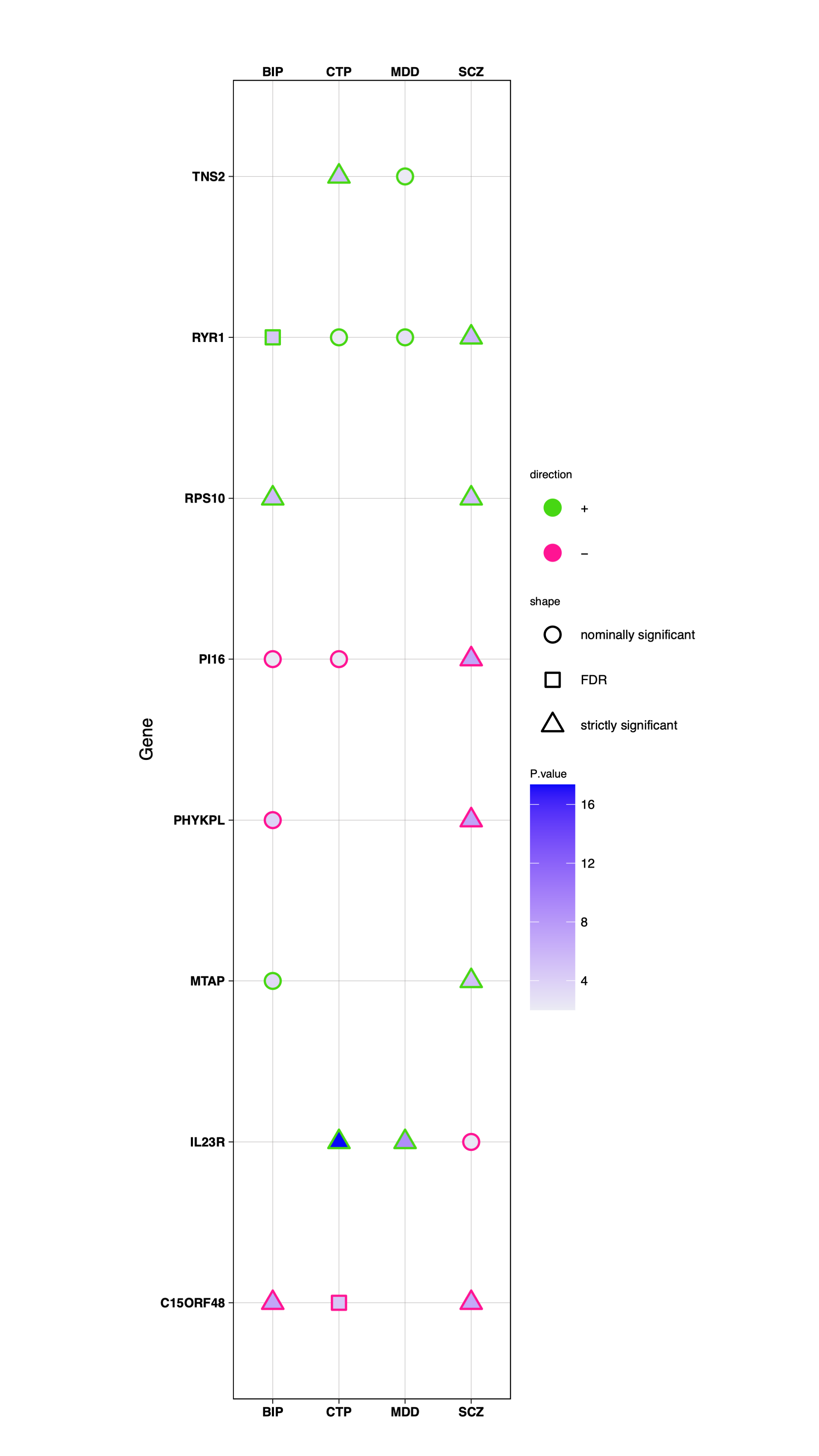

### Supplementary_Fig6A.png

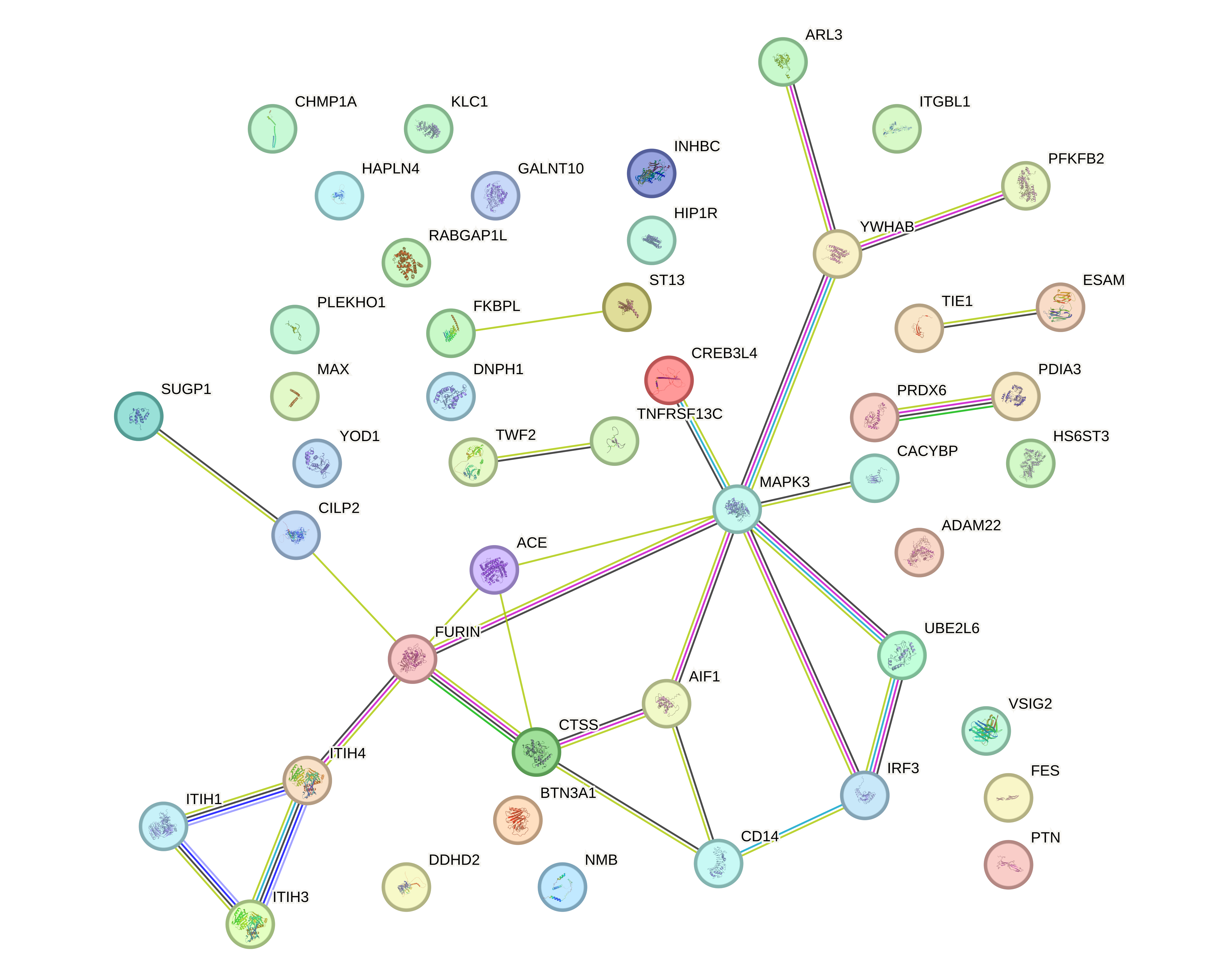

### Supplementary_Fig6B.png

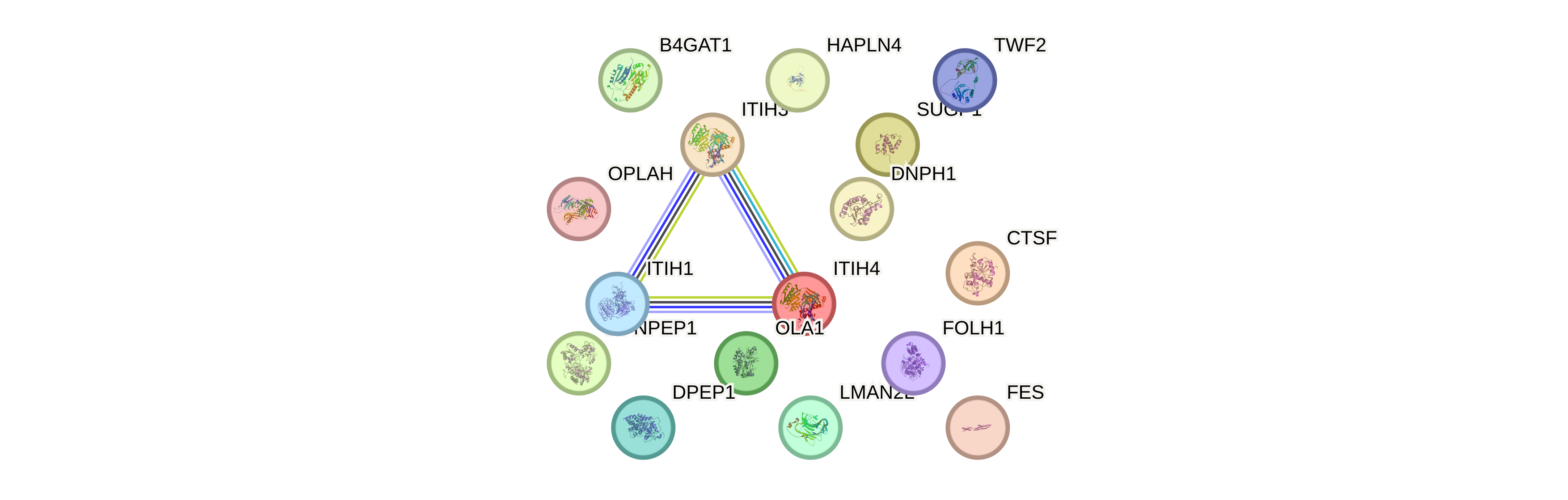

### Supplementary_Fig6C.png

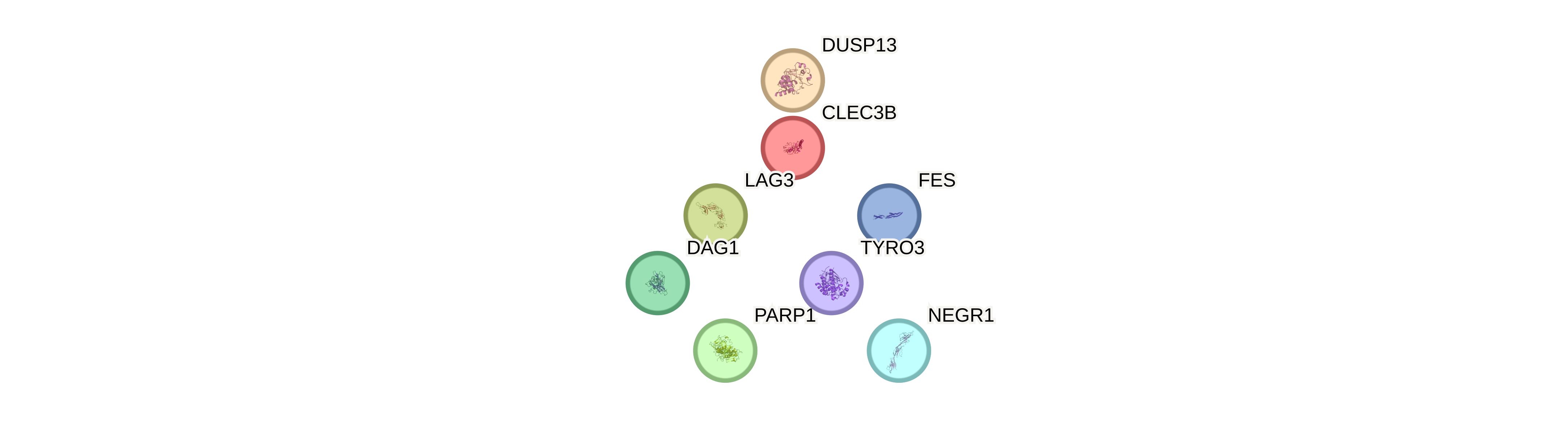

### Supplementary_Fig6D.png

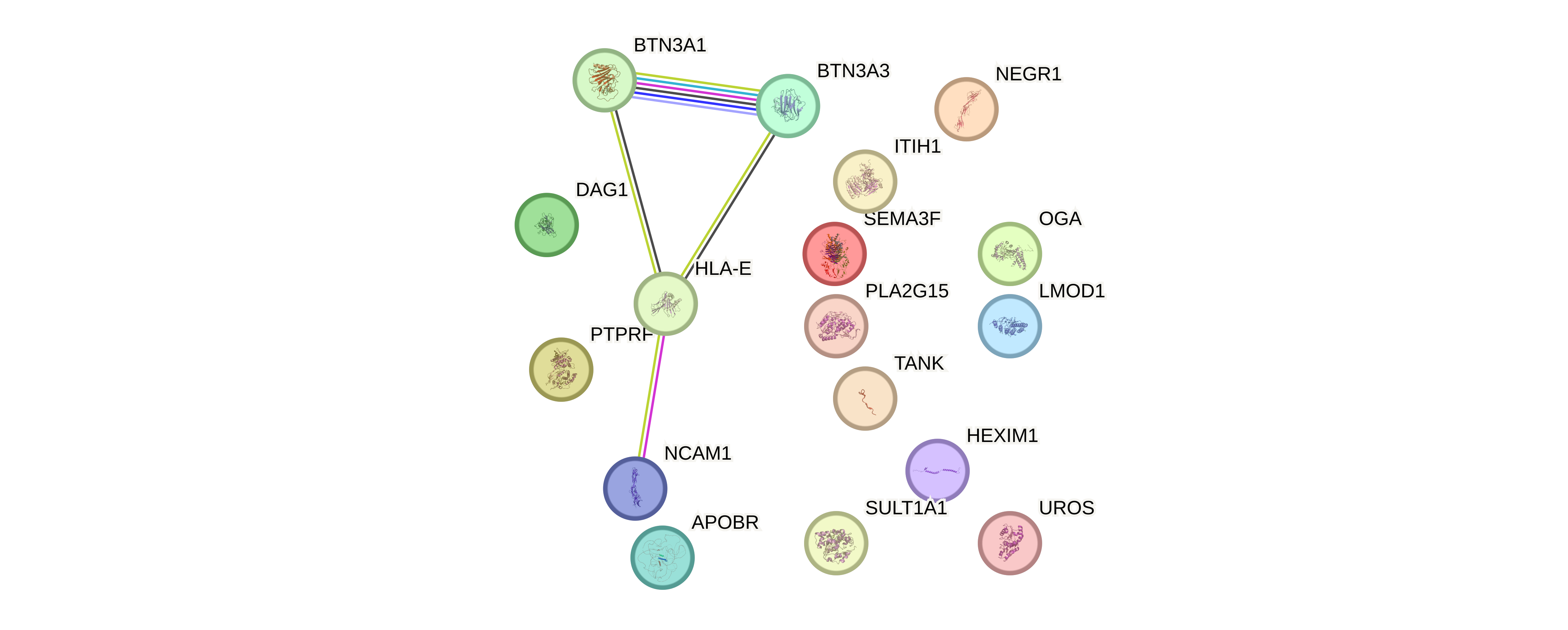

### Supplementary_Fig_4A_MR.tiff

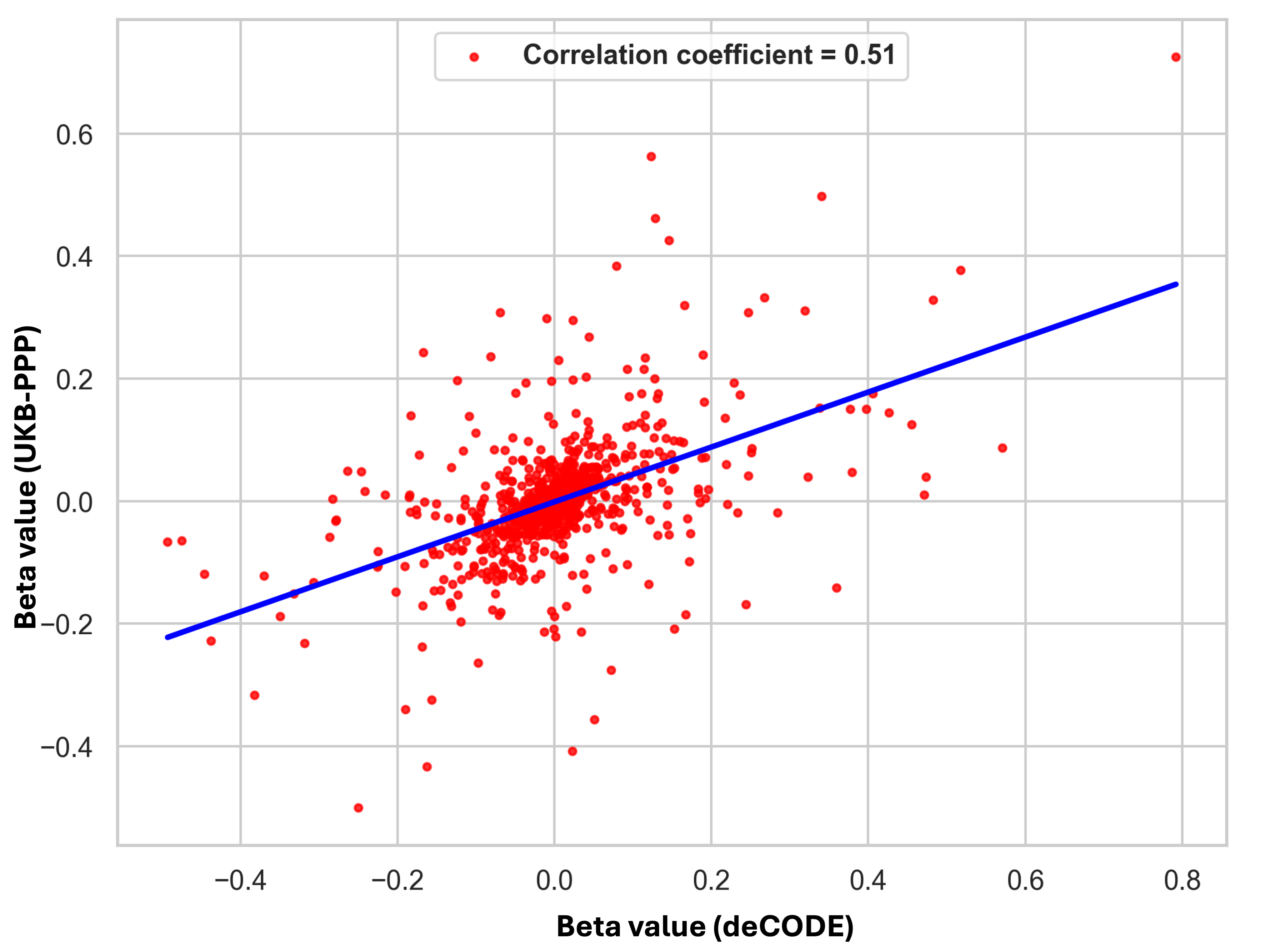

### Supplementary_Fig_4B_MR.tiff

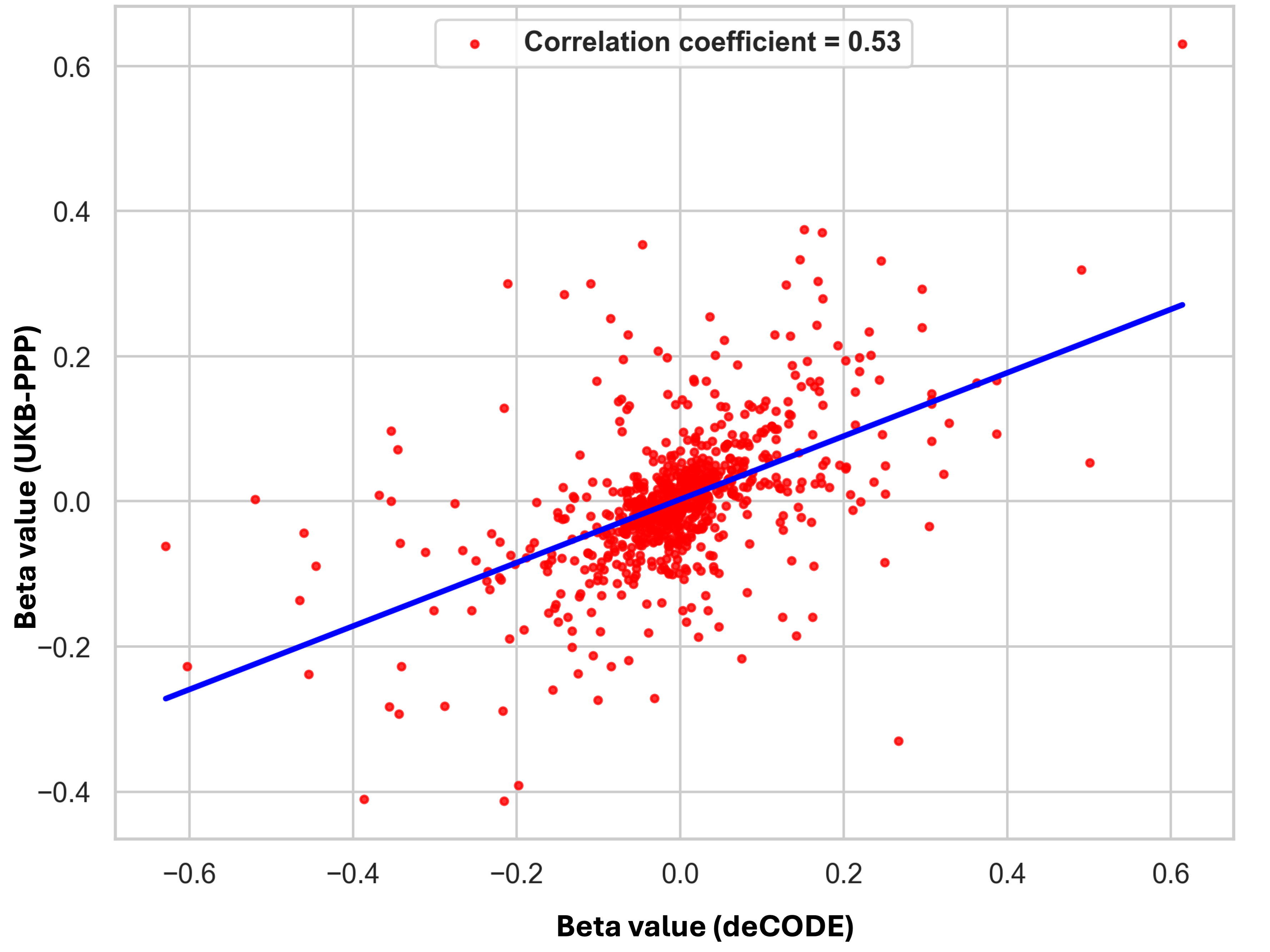

### Supplementary_Fig_4C_MR.tiff

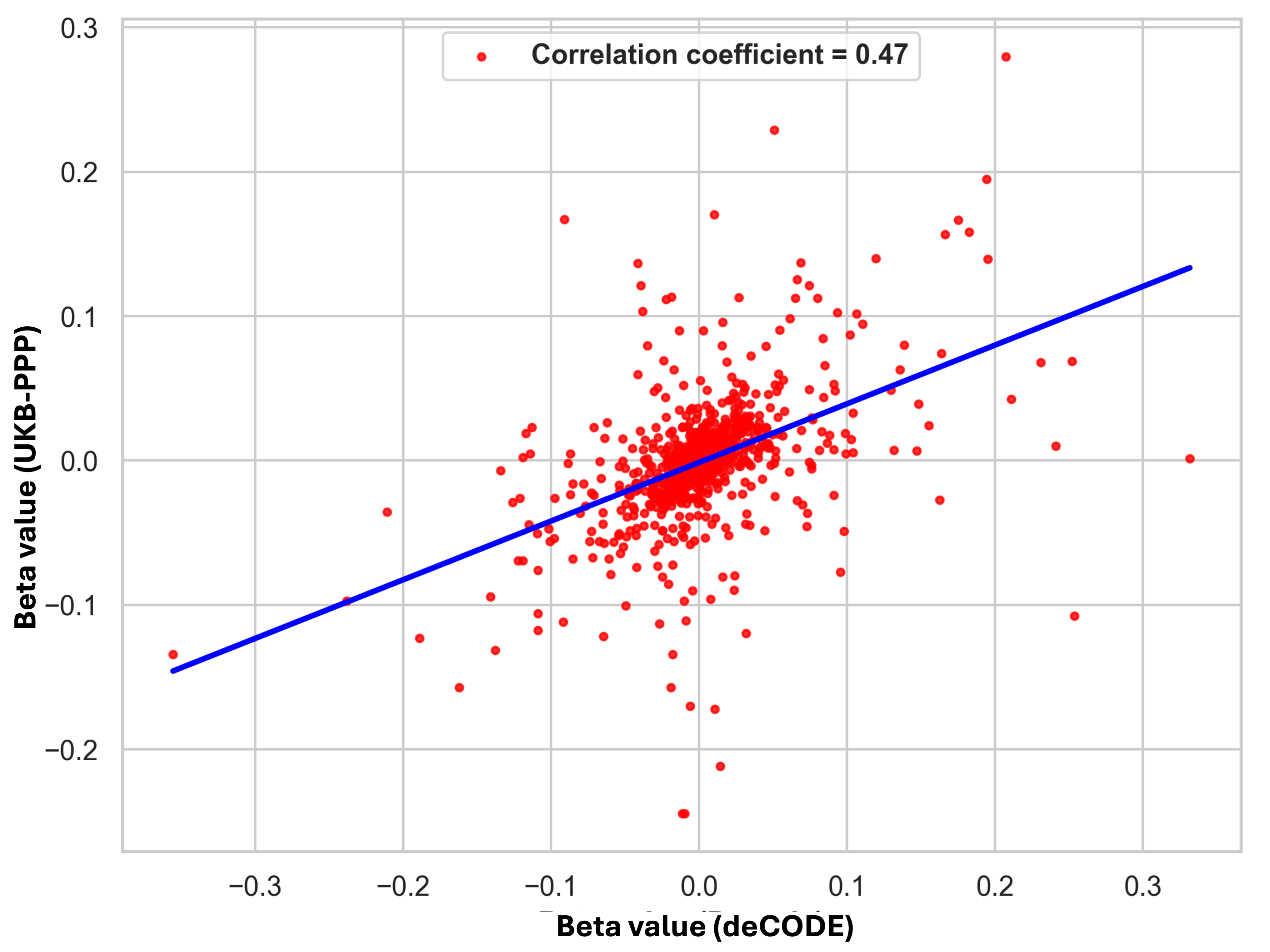

### Supplementary_Fig_4D_MR.tiff

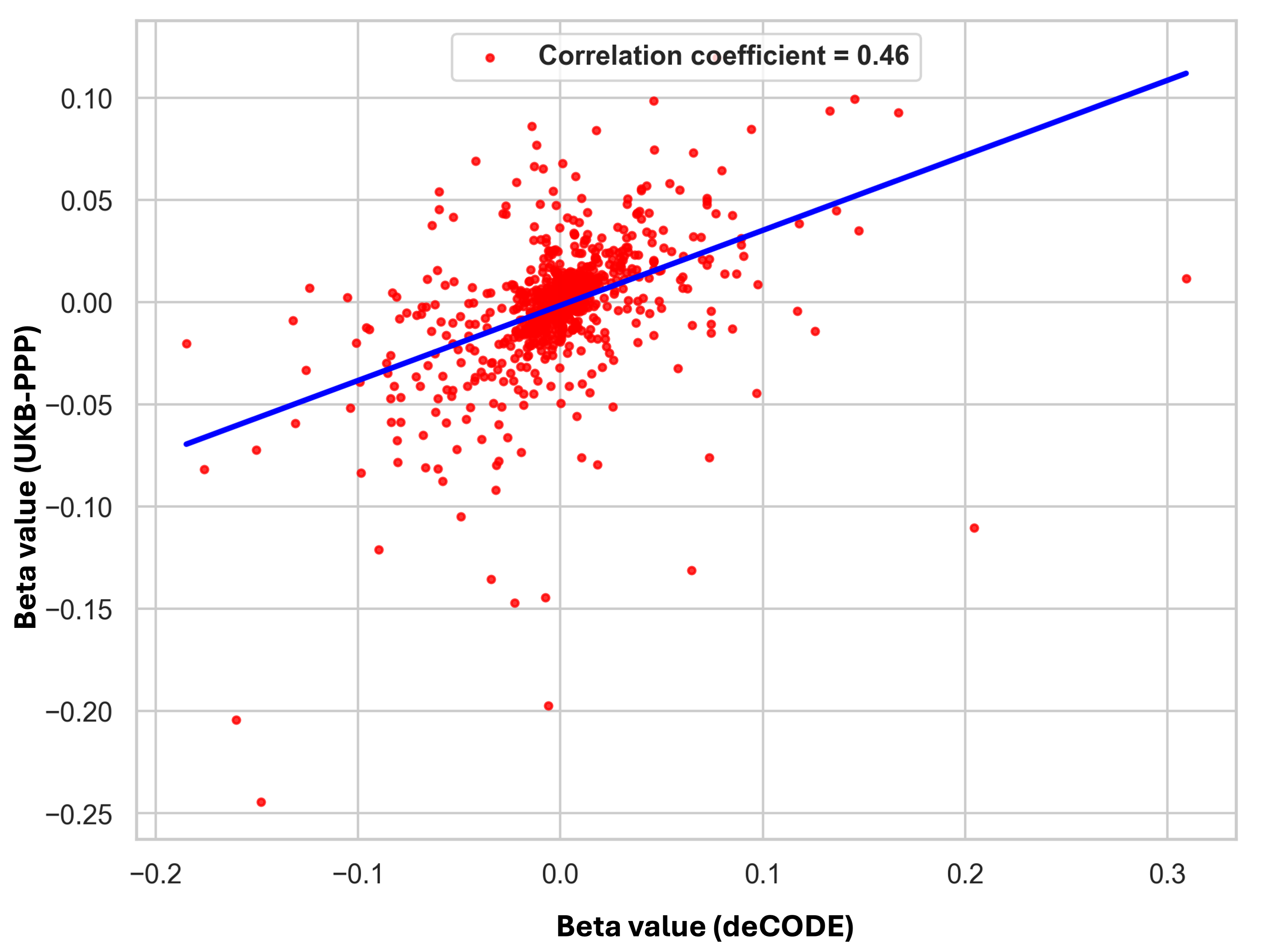

### Supplementary_Fig_5A_MR.tiff

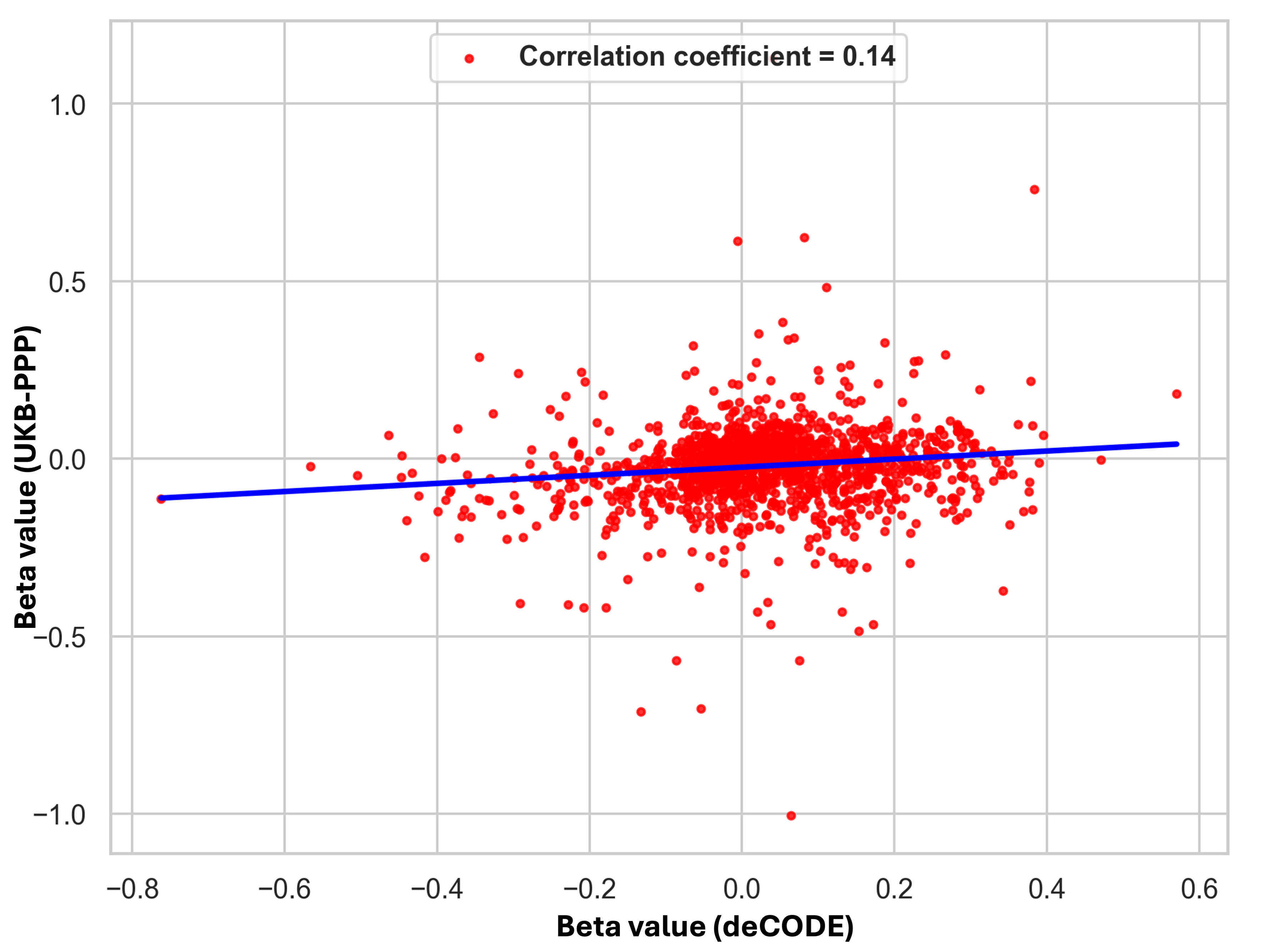

### Supplementary_Fig_5B_MR.tiff

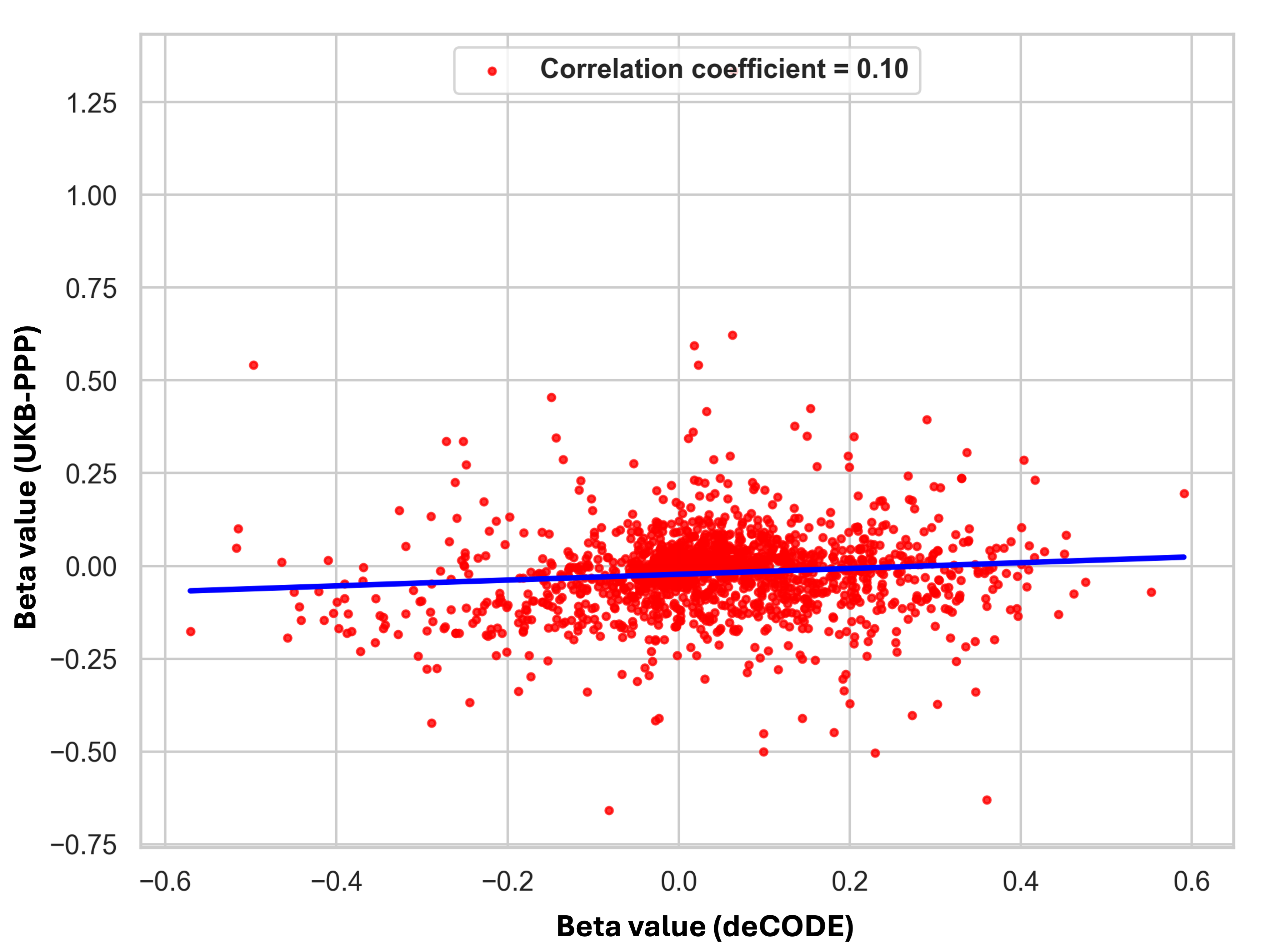

### Supplementary_Fig_5C_MR.tiff

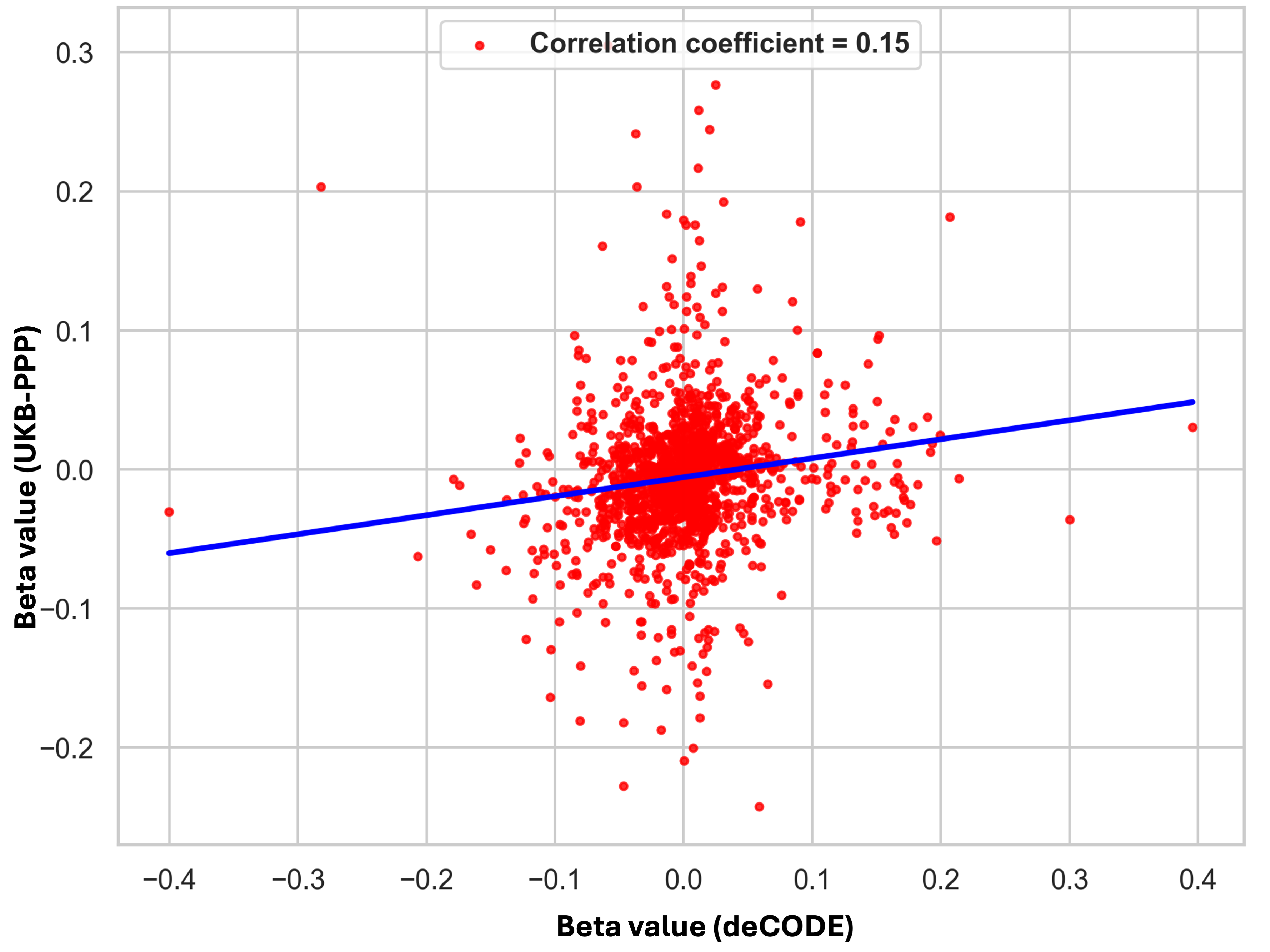

### Supplementary_Fig_5D_MR.tiff

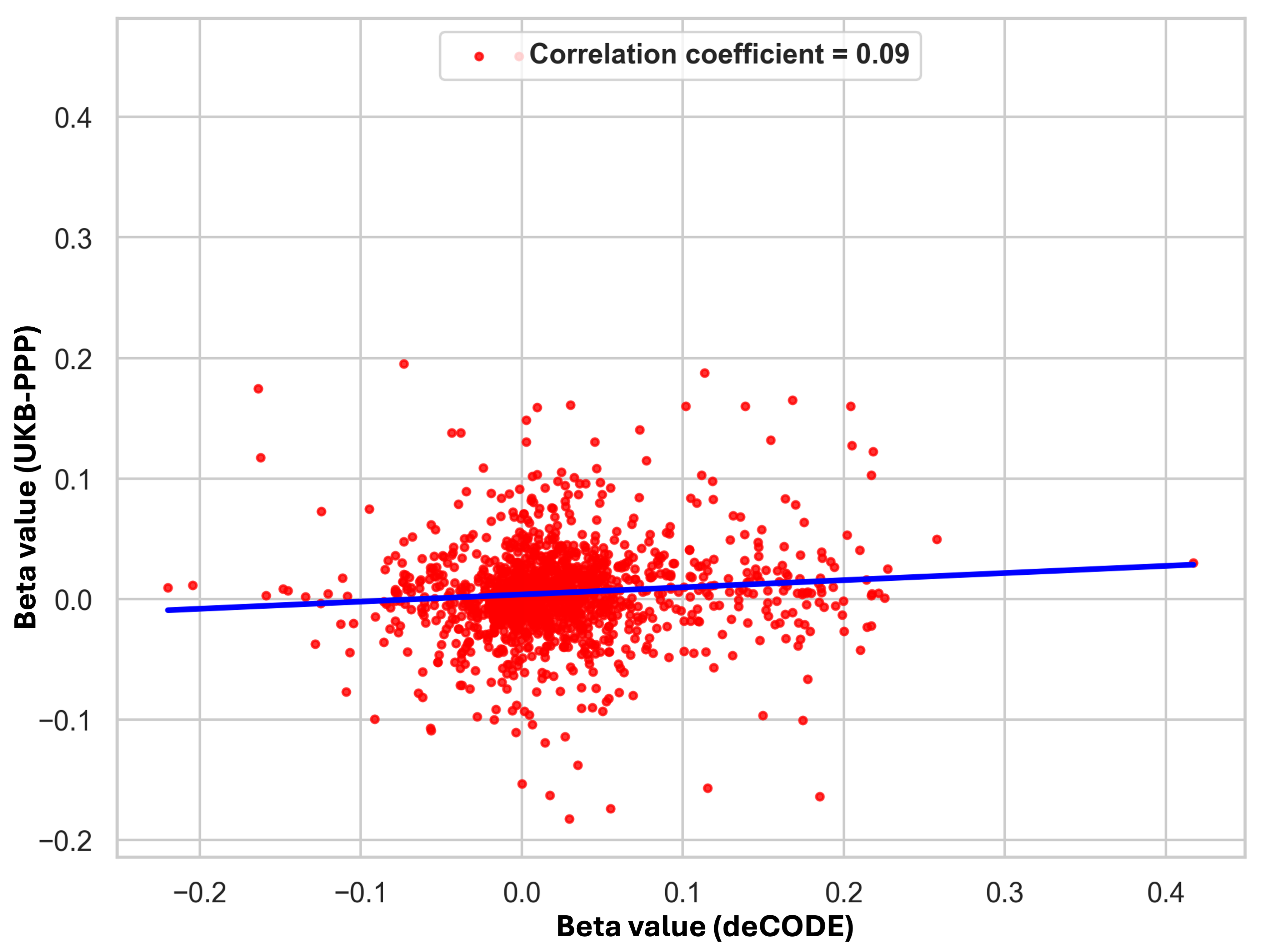

### Supplementary_Figure_2A_MR.tiff

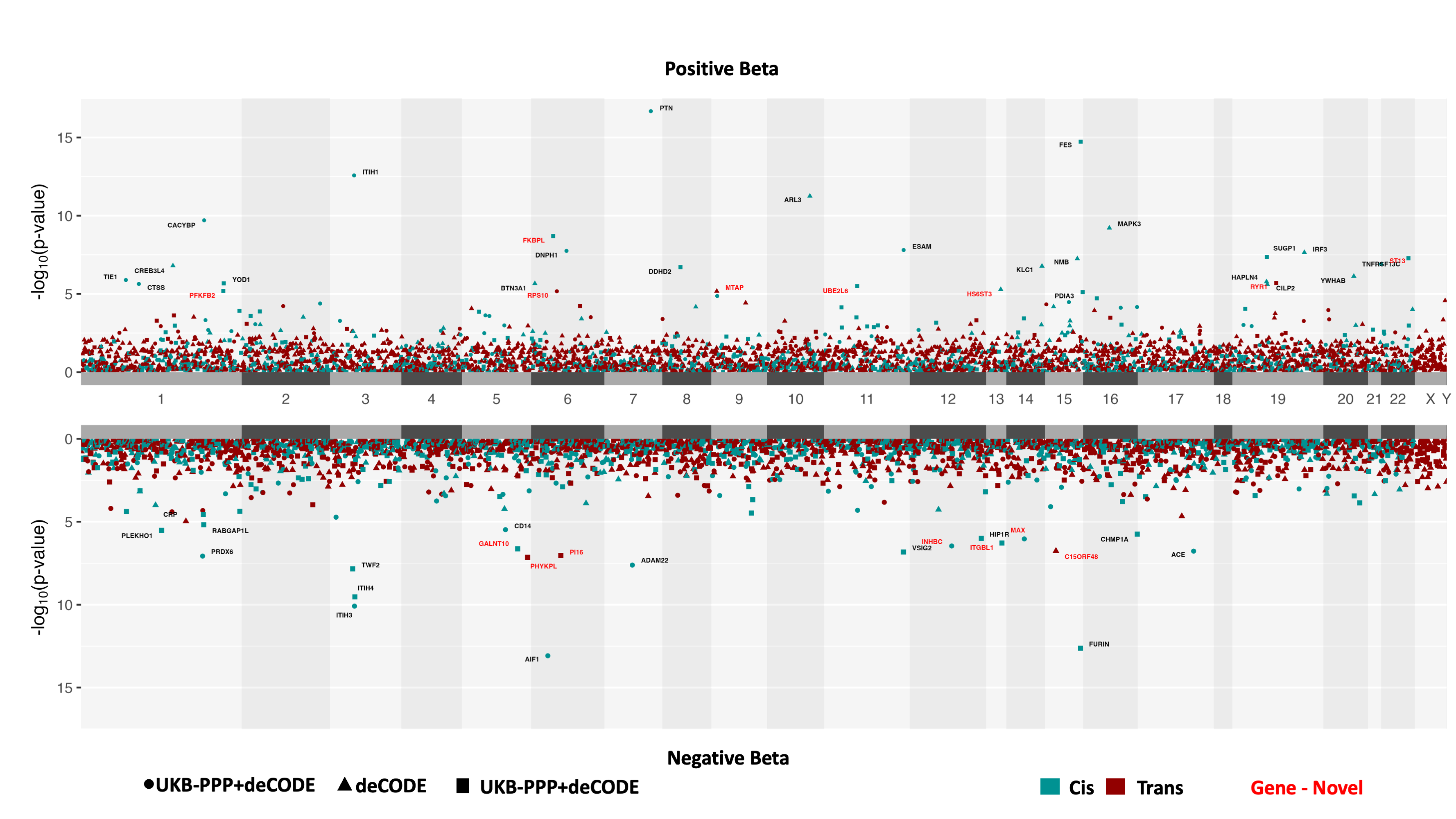

### Supplementary_Figure_2B_MR.tiff

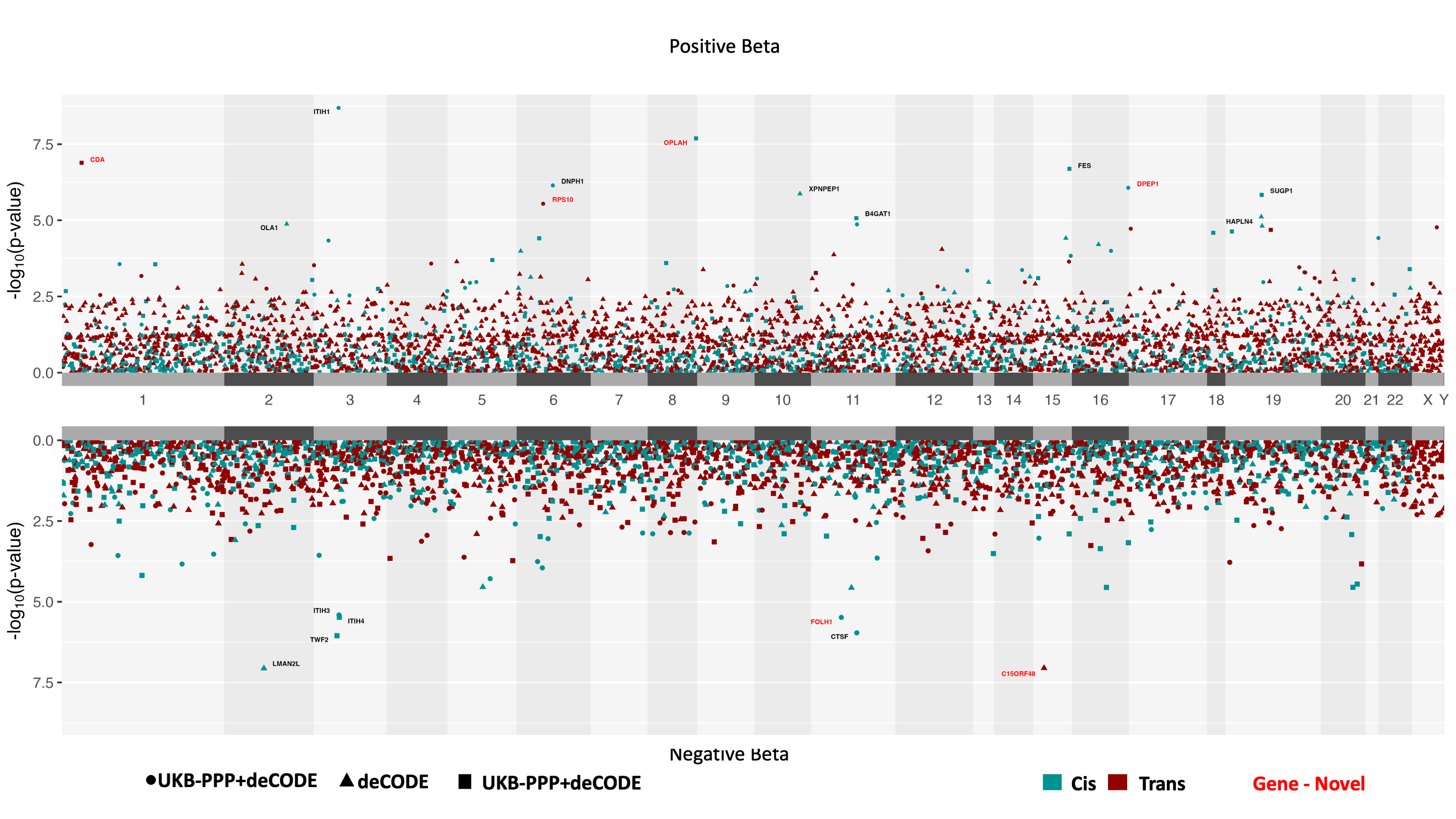

### Supplementary_Figure_2C_MR.tiff

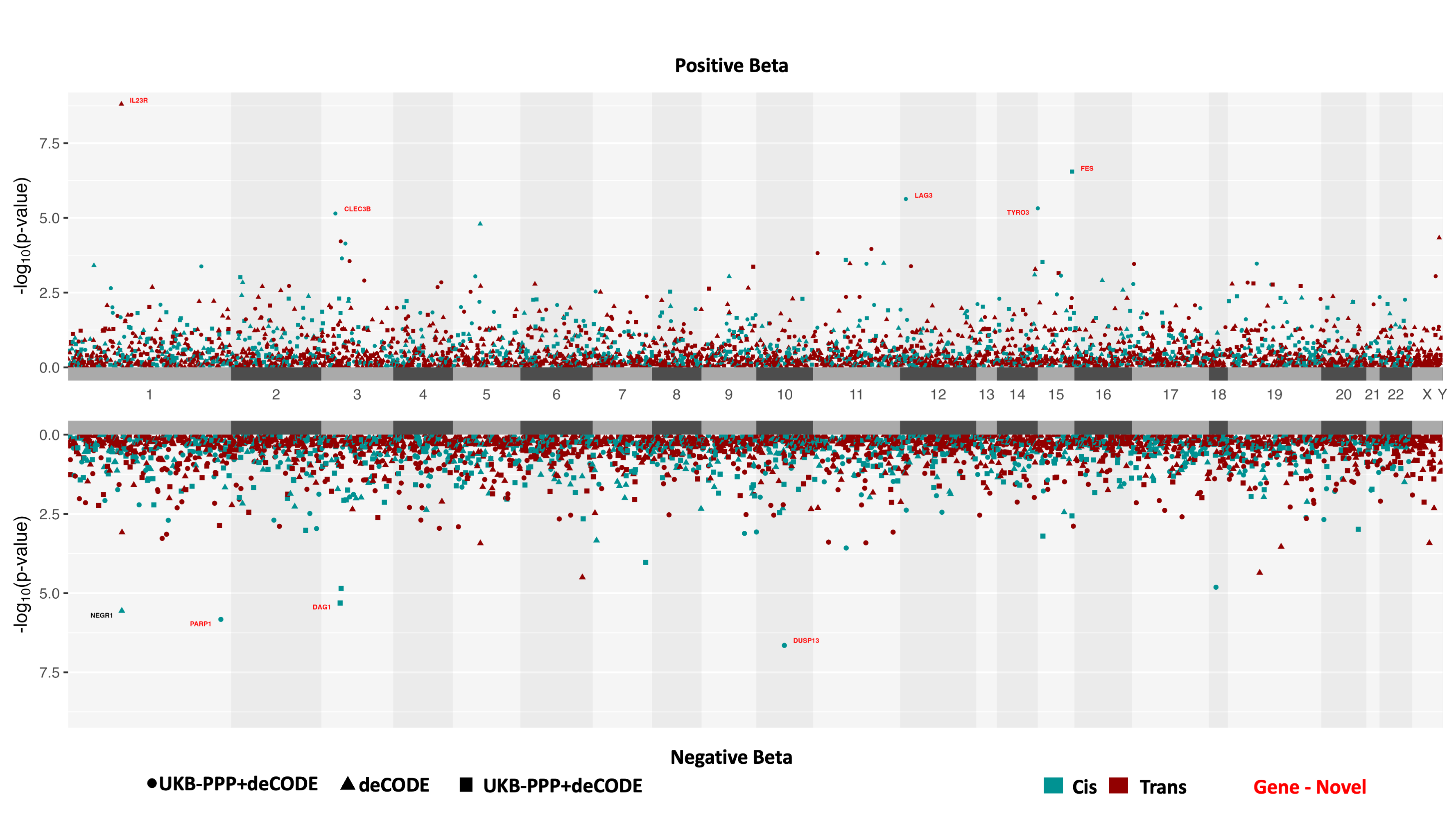

### Supplementary_Figure_2D_MR.tiff

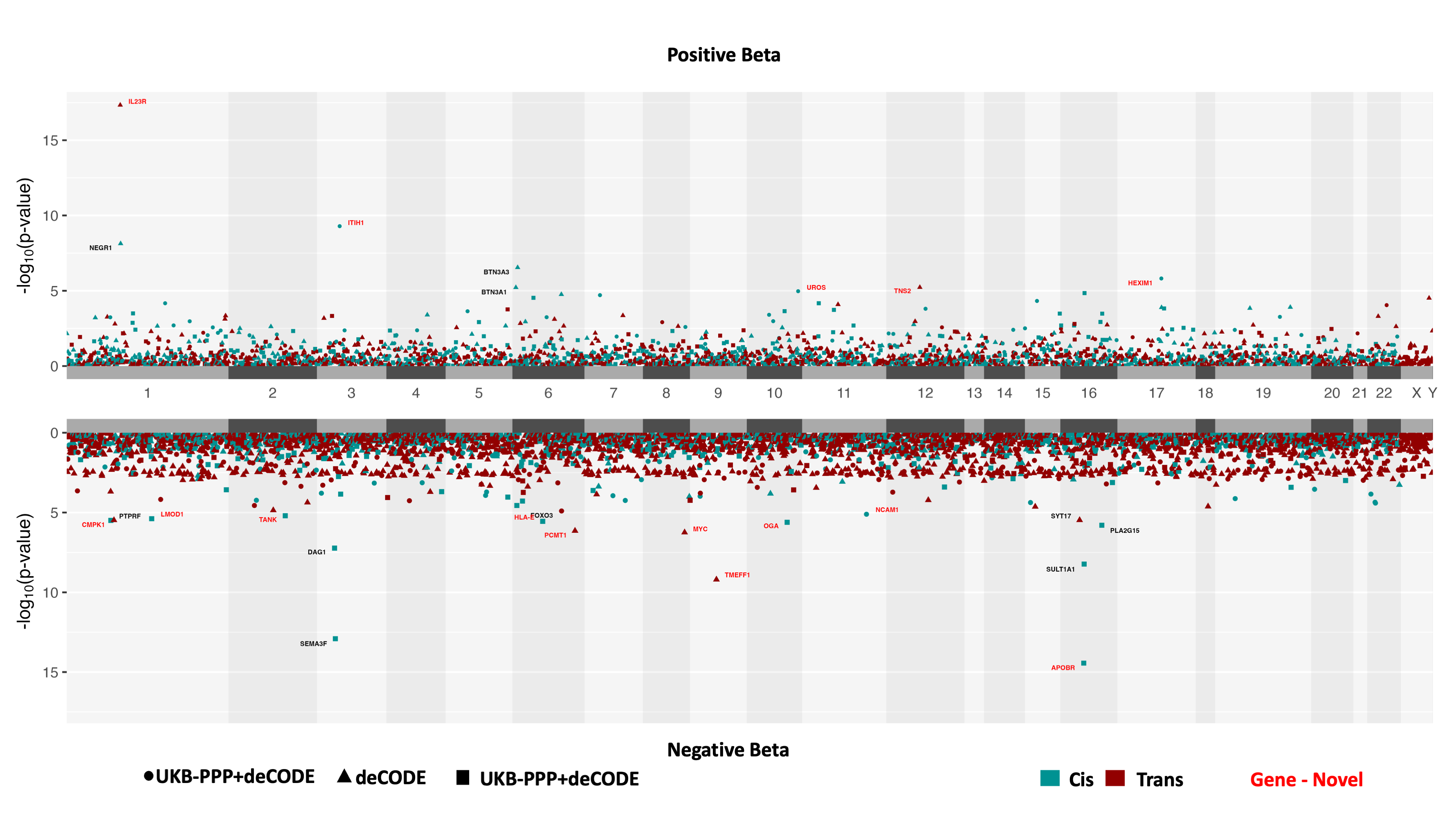
